# A study protocol for a multi-arm, parallel group randomized-controlled trial evaluating the individual and combined effects of behavioural promotion and hardware provision on handwashing with soap in peri-urban Lusaka, Zambia

**DOI:** 10.1101/2025.11.25.25340819

**Authors:** Katherine Davies, Jenala Chipungu, Katayi Mwila-Kazimbaya, Sarah Bick, Herbert Nyirenda, Anjali Sharma, Robert Dreibelbis

## Abstract

**Introduction:** Diarrhoea and lower respiratory infections remain leading causes of under-five mortality in sub-Saharan Africa. Handwashing with soap (HWWS) is a cost-effective intervention, yet adherence remains low. Effective interventions must address multiple behavioural influences, as knowledge alone is insufficient without supportive environments. Multi-component handwashing interventions target diverse behavioural determinants, yet few studies isolate the individual and combined effects of different components. This multi-arm randomised-controlled trial in Lusaka, Zambia, aims to assess the individual and combined effects of hand hygiene behavioural promotion and handwashing hardware and supply provision on HWWS at handwashing opportunities, strengthening the evidence base for designing sustainable handwashing interventions.

**Methods and analysis:** This superiority, multi-arm, parallel group randomised-controlled trial will be conducted in five peri-urban communities in Lusaka, Zambia, with 1800 households randomly assigned to four arms (450 households per arm) (1:1:1:1): (A) Handwashing hardware and supply provision only, (B) Hand hygiene behavioural promotion only, (AB) Hand hygiene behavioural promotion + hardware and supply provision, (C) control (no intervention). Households receiving the hardware and supply intervention (A, AB) will receive a handwashing facility and supplies to make liquid soap solution with a maintenance check after three-months. Households receiving the hand hygiene behavioural promotion intervention (B, AB) will receive five bi-weekly visits with a sixth follow-up visit four weeks later. HWWS behaviour of one household member will be assessed through structured observations at baseline and endline (6-months after intervention delivery begins).

**Ethics and dissemination:** Ethical approval was obtained from London School of Hygiene and Tropical Medicine, Research Ethics Committee, London, UK (Ref: 31387), the University of Zambia Biomedical Research Ethics Committee (Ref: UNZABREC 5834-2024) and the Zambia National Health Research Authority (NHRA-1823/23/12/2024). Results of the main trial will be submitted for publication in a peer-reviewed journal.

**Trial Registration Number:** NCT06865495; ClinicalTrials.gov; Pre-results; Initial release date 04-Mar-2025

**STRENGTHS AND LIMITATIONS:**

- The study provides an opportunity to directly assess the individual and combined impact of handwashing hardware and supply provision and behavioural promotion on handwashing with soap, which can inform future hand hygiene promotion programmes.
- The large study sample and comprehensive outcome measures will provide unique information about the individual and combined effects of intervention components on behaviour and health.
- While structured observations have inherent limitations (participants may alter their behaviour when they are aware they are being observed), it remains the gold standard for the measurement of routine behaviours in domestic contexts.
- The lack of blinding of study participants and limited blinding of assessors collecting data is a limitation to the study design. This is an acknowledged challenge in the behavioural and environmental health research sector.
- Since the intervention is implemented at the household level, there is some potential for contamination of the intervention message between households within communities. However, this risk is minimal given the large sampling interval. Contamination will be assessed during endline data collection.

## INTRODUCTION

Diarrhoea and lower respiratory infections remain leading causes of mortality in children under-5, with the highest burden observed in sub-Saharan Africa. Cause-specific mortality estimates from 2019 attributed 9.1% of global under-five deaths to diarrhoea and 13.9% to lower respiratory infections [1]. Interventions promoting handwashing with soap (HWWS) have been shown to reduce the risk of acute respiratory infection (ARI) by 17% and diarrhoea by 30% in low-income and middle-income settings [2, 3]. While global and regional disease outbreaks, such as COVID-19 and cholera, have driven renewed global attention to hand hygiene interventions, HWWS practice remains low, particularly in low-resource settings [4, 5]. Households with a dedicated handwashing facility are approximately twice as likely to practice HWWS after faecal contact compared to those without [5, 6]. However, these estimates are largely based on cross-sectional data, limiting insights into causal mechanisms. Global monitoring suggests that 73% of households in sub-Saharan Africa lack access to a handwashing facility with soap and water at home [7]. Cost and affordability of handwashing facilities and materials are frequently cited as barriers to HWWS globally [8].

While there is robust evidence linking changes in key hygiene behaviours to health outcomes, the mechanics of supporting and promoting lasting changes in individual behaviour remain uncertain. Traditional handwashing interventions often focus on cognitive determinants of HWWS, with limited attention to a broad range of behavioural and infrastructural factors. Several systematic reviews of handwashing determinants in community settings found that knowledge of disease transmission alone is insufficient to drive HWWS, particularly in the face of unconducive behavioural settings or competing priorities [8–10]. The reviews highlight the critical role of environmental enablers – such as the presence of a dedicated handwashing facility – in overcoming psychological trade-offs that hinder HWWS. Successful handwashing interventions therefore must integrate multiple elements to address these diverse influences [11, 12].

A recent scoping review of hand hygiene guidelines highlighted a lack of clear, evidence-based guidance on how to develop, implement and sustain hand hygiene behaviour interventions [13]. Multi-component interventions targeting cognitive, behavioural and infrastructural determinants of hand washing are needed. However, typical evaluation approaches comparing a multi-component intervention as a single package against a control group limit the ability to isolate the effects of individual components and understand their specific contributions to HWWS behaviour change [14, 15]. A recent systematic review reported substantial heterogeneity in the behaviour change techniques used across hand hygiene interventions and noted that the complexity of intervention packages often hinders the ability to disentangle the individual and combined effects of specific components [16]. The authors emphasised the need for research that is intentionally designed to evaluate these effects.

This multi-arm randomised-controlled trial aims to address this evidence gap by evaluating the individual and combined effects of handwashing hardware and supply provision and hand hygiene behavioural promotion on HWWS at handwashing opportunities (i.e. when hands could have been washed or not) in peri-urban communities in Lusaka, Zambia. By examining the impact and interactions of these intervention components, the study aims to enhance the evidence base for designing more effective and sustainable handwashing programmes.

### Objectives

1. To evaluate the individual and combined effects of intervention components on HWWS behaviour at handwashing opportunities (Table 1).
2. To evaluate the individual and combined effects of intervention components on HWWS at specific handwashing opportunities (e.g., before eating) (Table 1).
3. To evaluate the individual and combined effects of intervention components on hand-rinsing at handwashing opportunities (Table 1).
4. To assess if water insecurity is an effect modifier on the relationship between the intervention components and hand hygiene behaviour.
5. To explore the impact of the intervention components on handwashing knowledge and self-reported handwashing.
6. To explore the durability of handwashing facilities over a 6-month period.

## METHODS AND ANALYSIS

### Study Design

The study is a superiority, multi-arm, parallel group randomised-controlled trial assessing both the individual and combined effects of two interventions - hand hygiene behavioural promotion and handwashing hardware and supply provision - on observed HWWS. Given the potential for interaction between intervention components, a multi-arm trial using an “inside-the-table” analysis will be used, rather than a factorial trial (using an “at-the-margins” analysis) [18, 19].

**Table 1.** Handwashing Opportunities.

| Handwashing Opportunity |
| --- |
| Before cooking |
| Before eating |
| Before feeding a child (spoon feeding, manual feeding or serving) |
| Before breastfeeding |
| After toilet use |
| After handling child/adult faeces or cleaning a child's bottom |
| After contact with animals |
| After coughing or sneezing |

**Table 2.** Research Question (PICOT format) [17].

|  |  |
| --- | --- |
| **Research Question** | What are the individual and combined effects of handwashing hardware and supply provision and behavioural promotion on handwashing with soap (HWWS) behaviour at handwashing opportunities? |
| **Population** | Households in peri-urban communities of Lusaka, Zambia with at least one child under the age of 5, no fixed handwashing facility, and at least one adult (18+) who can consent to the study. Participants must be permanent residents of the selected communities (i.e., expect to remain in the community for the next 3-6 months). |
| **Intervention** | Households randomised to one of four arms (1:1:1:1): (A) handwashing hardware and supply provision only, (B) hand hygiene behavioural promotion only, (AB) hand hygiene behavioural promotion + handwashing hardware and supply provision, (C) no intervention. At the end of the study period, the most effective combination of interventions will be delivered in the control group. |
| **Comparison** | Our analysis will concern five pairwise comparisons:<br>1) Behavioural promotion against control (B vs. C)<br>2) Hardware and supply provision against control (A vs. C)<br>3) Behavioural promotion + hardware and supply provision against control (AB vs C)<br>4) Behavioural promotion + hardware and supply provision against hygiene promotion alone (AB vs B)<br>5) Behavioural promotion + hardware and supply provision against hardware and supply provision alone (AB vs A). |
| **Outcome** | The primary outcome is the probability of HWWS within three minutes of any handwashing opportunity (Table 1) in the correct sequence (e.g., before or after the relevant opportunity). |
| **Time** | Outcomes measured at baseline and endline (6-months after intervention delivery begins). |

A one-stage approach will be used to randomise 1800 households to one of four trial arms (450 households per arm) (1:1:1:1): (A) handwashing hardware and supply provision only, (B) hand hygiene behavioural promotion only, (AB) hand hygiene behavioural promotion + handwashing hardware and supply provision, (C) control (no intervention) (Fig 1). A block randomisation approach will be used during allocation of households to study arms, ensuring proportionate representation of participants across study areas. This protocol was drafted using the SPIRIT reporting guidelines [20]. The SPIRIT checklist can be found in supplementary material (S2 Table).

**Fig 1.**
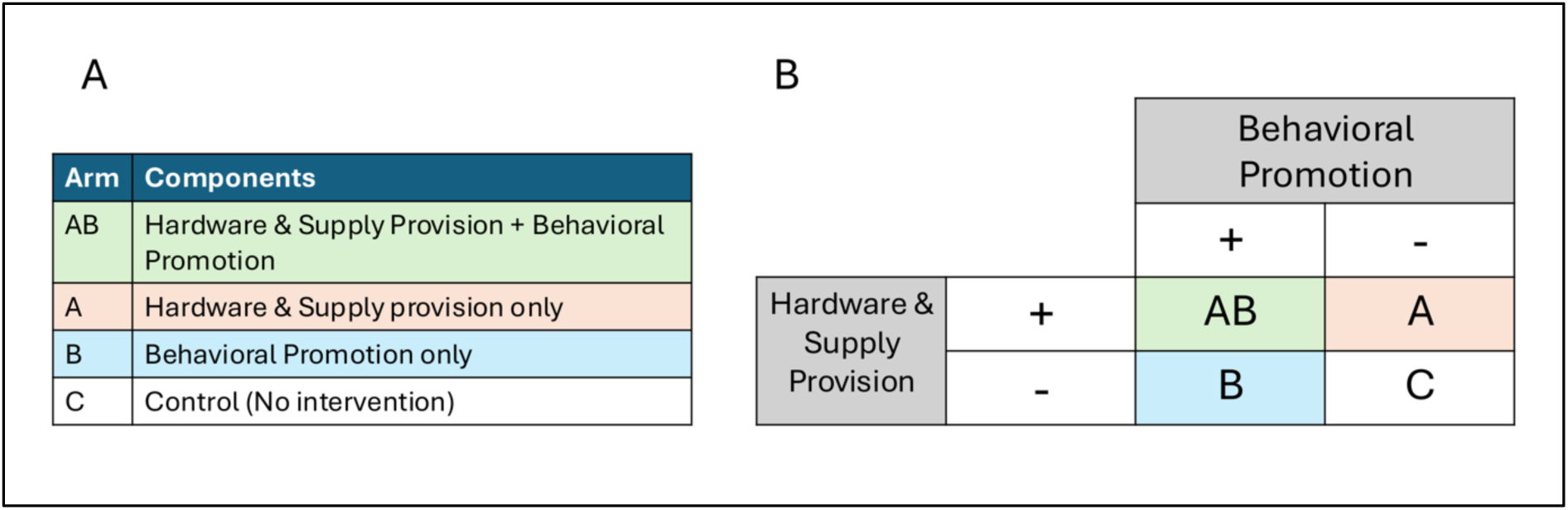
(A) Description of the intervention components within each arm and (B) Table of the 2×2 multi-arm study design.

### Study Setting

This community-based study is based in Lusaka, the capital and largest city in Zambia. Nearly 62% of Lusaka’s population live in peri-urban areas [21], characterised by dense, informal settlements, with unplanned housing and persistent challenges in accessing water, sanitation and hygiene (WASH) services. These communities experience seasonal flooding, leading to cyclical cholera outbreaks and other public health risks. Several WASH-related indicators - including limited soap use for handwashing, reliance on unimproved sanitation facilities, and uncovered household water storage - were identified as risk factors for waterborne and diarrhoeal illness in these communities [22]. An estimated 71% of the urban population in Zambia lack access to a household handwashing facility with soap and water [23].

Lusaka city is in Lusaka district, which is divided into 33 wards (small administrative units). Wards where we recently conducted feasibility research for this study were excluded (n=2). From the remaining wards, five were selected in collaboration with the study team, drawing on prior engagement with the District Health Office’s Environmental Health Technician (EHT). The selection was based key factors including population characteristics, geographical boundaries, access to WASH services and the burden of hygiene-related diseases. Although ward selection was not random, the findings will remain fully generalizable to low-income populations in the region. The five selected wards are Chainda, Mpulungu, Chawama, Mtendere, and Kanyama (Fig 2).

**Fig 2.**
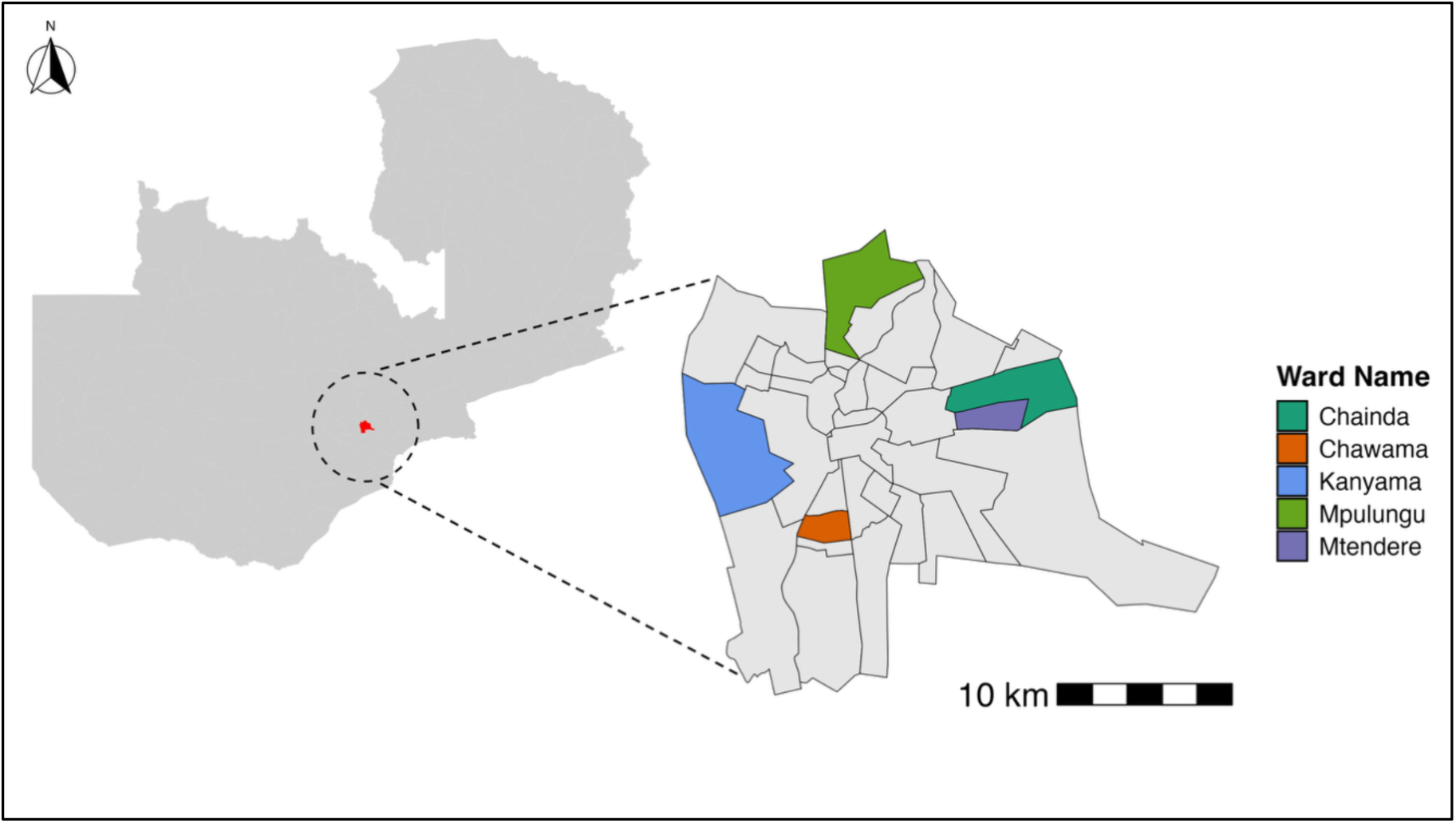
Map of Zambia with Lusaka District highlighted in red (left) and map of Lusaka District with the five selected study wards highlighted (right). Administrative boundaries of Zambia were sourced from the Humanitarian Data Exchange (HDX), Zambia – Subnational Edge-matched Administrative Boundaries, licenced under CC BY-IGO (available from: https://data.humdata.org/dataset/cod-em-zmb) (left graph) and Stanford Digital Repository – Zambian 2006 to 2010 Constituency and Ward Boundaries, not subject to copyright (available from: http://purl.stanford.edu/yc436vm9005) (right graph) [24].

### Eligibility Criteria

Households eligible for the trial must meet all the following criteria at enrolment:

- At least one adult (aged 18 or older) who can consent to the study on behalf of all members of the household.
- At least one child under the age of 5.

Exclusion criteria:

- Non-permanent resident of the selected community and/or plans to leave the community within the next 3-6 months
- Household already own a fixed handwashing facility (e.g., sink, handwashing station).

The head of household or a nominated representative will provide written, informed consent for household participation in the study. Data collection activities will be completed with the adult (over 18 years or older) most responsible for caring for the child under the age of 5 at the time of data collection. If a child under 5 is not present, the adult (18 years or older) most responsible for childcare will be selected for observation. If such an individual is not present, another randomly selected household member (18+) will be observed instead. Separate consent will be obtained from the household member participating in data collection activities.

### Interventions

This household-level, multi-component intervention includes the delivery of hand hygiene behavioural promotion and handwashing hardware and supplies to households in pre-selected peri-urban communities in Lusaka, Zambia. The intervention is outlined according to the TIDieR-WASH guidelines [25], for which the checklist can be found in supplementary material (S3 Table).

A Theory of Change (ToC) was developed to illustrate how the intervention components are expected to increase HWWS, and how this may be influenced by other factors (Fig 3). We hypothesize the behavioural messaging component (Arms AB + B) will improve the capability, opportunity, and motivation of household members to practice HWWS. For households receiving hardware and supplies (Arms AB + A), we hypothesize the presence of a HWF with soap and water in the household will increase the opportunity for HWWS at key opportunities. Assumptions that need to hold true for change to occur are outline in the ToC.

**Fig 3.**
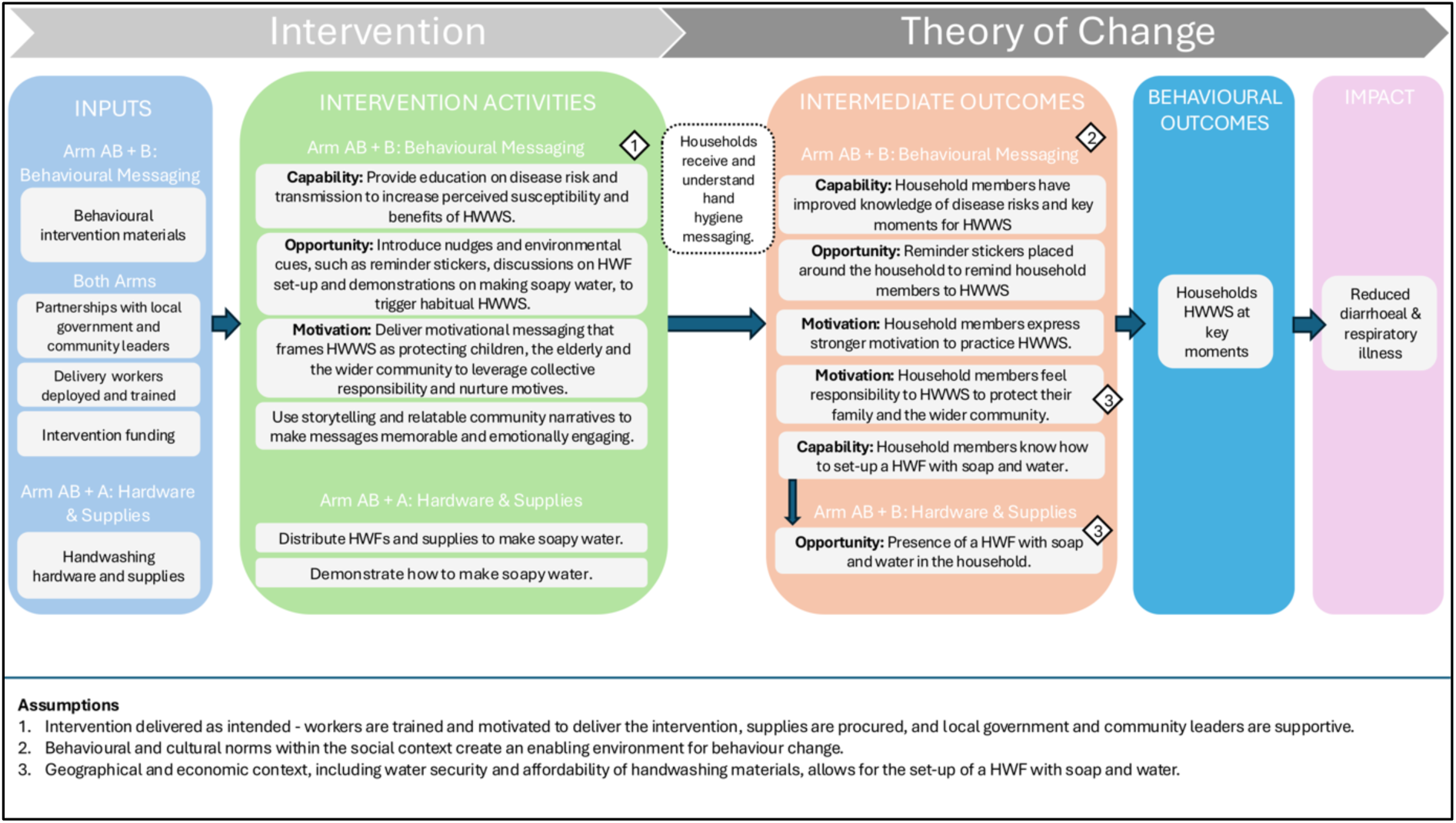
A Theory of Change (ToC) for the behavioural messaging (Arms AB + B) and hardware & supplies provision (Arms AB + A) components of the intervention. Elements specific to each intervention component are grouped under sub-headings to highlight their unique pathways. Assumptions that need to hold true for change to occur are detailed at the bottom of the figure.

This multi-component intervention will be delivered by a team of community volunteers known as neighbourhood health committee (NHC) members all of whom are residents of the selected communities. These volunteers typically oversee health promotion efforts, vaccination campaigns and disease surveillance within the communities they serve. They will be recruited on a short-term basis by the local research partner (CIDRZ) and overseen by the research team there. Intervention delivery teams will be given custom t-shirts with the intervention logo to create a distinct visual identity for the team and avoid direct associations with staff employed for data collection. The interventions will be delivered to all members of the household present at the time of delivery.

### Handwashing Hardware Provision (Arms AB + A)

Households will be provided with a locally available handwashing facility called the Kalingalinga bucket (Fig 4, A), which was the favoured handwashing facility out of several designs in a previous feasibility study [26]. Facilities are manufactured locally, typically by metal workers who operate from a community called Kalingalinga, after which the bucket and stand are named. Handwashing facilities will be tested and quality checked prior to distribution. As cost and affordability of handwashing materials are commonly reported barriers to HWWS [8], and this study will be conducted in low-income communities, handwashing facilities will be provided to households free of charge. The cost of the facility is a maximum of $10 USD. Of note, $17 USD per household is the mean costs included in economic models of the cost of achieving universal hand hygiene in domestic settings [27], thus representing a viable intervention option for scale.

**Fig 4.**
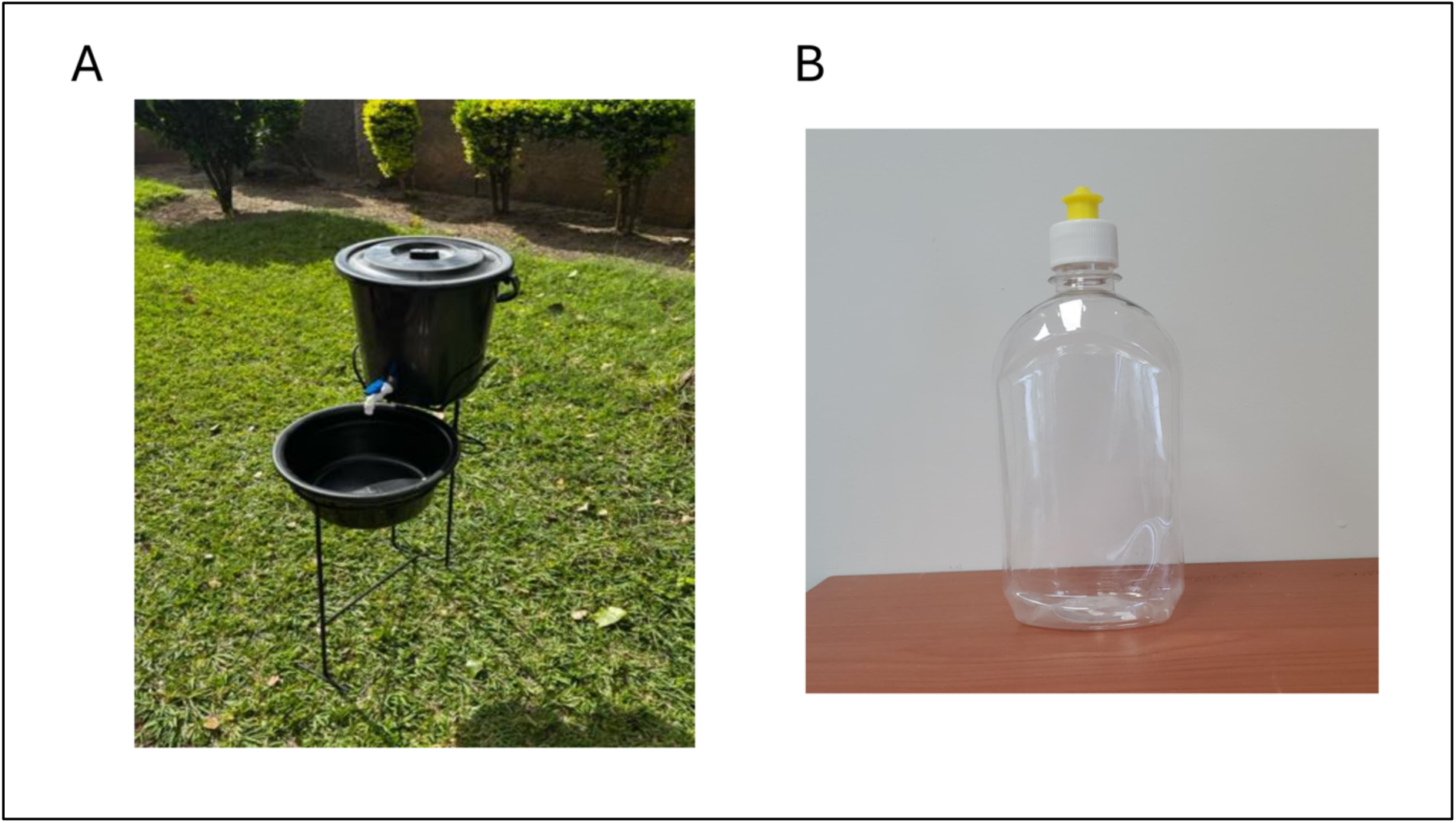
(A) Kalingalinga Bucket and (B) Plastic detergent container for soapy water. Both photographs are original and were taken by the author.

Households will also be given instructions on how to make liquid soap (soapy water) from locally available soap products. Households will be provided with powdered soap and a hard plastic detergent container (Fig 4, B). Intervention delivery workers (NHC members) will set-up the handwashing facility, and demonstrate how it works, including where to place soap near the stand. Discussions will also be held on maintenance and positioning of the handwashing facility when it is provided. Participants will be advised to report if their handwashing facility is damaged or stolen between follow-up visits. Any problems will be immediately addressed, or the handwashing station replaced. Intervention delivery workers will return to households after three months to ensure there are no issues with the handwashing station (e.g., broken stand or tap, or stolen), to bring more powdered soap and to remind households how to make soapy water. Both visits should take no longer than 30 minutes.

### Hand Hygiene Behavioural Promotion (Arms AB + B)

Household-based hand hygiene behavioural promotion will be based on an innovative intervention informed by multiple behavioural theories and models [28–30]. Households will receive a behavioural intervention called *Ulupwa* (The Family Intervention), which employs interactive storytelling and visual aids to promote HWWS. Designed as a participatory experience, *Ulupwa* actively involves families through storytelling, games and discussion.

Intervention delivery workers will act as storytellers, narrating stories about three characters with different handwashing habits. Interactive elements are embedded throughout to encourage reflection on personal handwashing behaviours, explore common barriers to sustained practice, and support families to develop their own practical solutions. A key prop for the intervention is a set of visual ‘playing’ cards that illustrate the characters, handwashing barriers and other related concepts. The intervention supports the household to realise all family members should take responsibility for maintaining safe handwashing practices and is designed to involve the whole family. The intervention progresses through five 30-minute visits conducted on a bi-weekly basis, with a final sixth visit 4-weeks after the fifth visit (3-months after the first visit) (Table 3). More details are outlined in supplementary material (S1 Text).

**Table 3.** Intervention Visit Content.

| **Visit Topic** | **Content** |
| --- | --- |
| **Visit 1: Identifying Behaviours** | This visit aims to build a connection and trust with the family and create an environment where resources can lead to transformative conversations. During the visit, storytellers will understand handwashing behaviours of each family member and discuss their perceived handwashing barriers. |
| **Visit 2: Understanding disease risks** | This visit tells a story about Nevers, a boy who does not understand the importance of handwashing behaviour. The visit aims to build awareness around the disease risks associated with inadequate handwashing with soap (HWWS) and provide information on when to wash hands. |
| **Visit 3: Encouraging practical solutions** | This visit tells the story of Sami, Nevers' sister, who knows handwashing is important but often forgets or does not have time to wash hands. This visit aims to introduce nudges or other environmental cues to prompt HWWS. During the visit, households not receiving hardware will receive guidance on how to set-up and position a handwashing facility in their home and will be shown how to make soapy water. All households will be given handwashing reminder cards to position around the household. |
| **Visit 4: Promoting practice** | This visit tells the story of Always, Sami and Nevers' cousin, who always washes hands with soap and water and wants to ensure all family members take responsibility for maintaining safe handwashing practices. This visit aims to motivate all household members to take responsibility for handwashing, focussing on protecting household members and the community from disease. Wider benefits such as improved school attendance will also be discussed. |
| **Visit 5: Reflection and Commitment** | This visit reinforces key messages across the intervention period and aims to encourage participants to re-assess their behaviours and pledge to adopt improved hygiene practices. |
| **Visit 6: Final Visit** | This visit asks participants to reflect on the intervention as a whole and covers any information participants would like to revisit. This includes making more soapy water if relevant. |

Households in the comparison group will receive no intervention during the study period. However, at the end of the study period, the most effective combination of interventions will be delivered to these households.

### Criteria for discontinuation

Households that miss more than three consecutive visits will not receive further intervention visits. However, the trial will follow the intention-to-treat principle, so no households will be excluded from the analysis regardless of discontinuation.

### Monitoring Adherence

Intervention delivery workers will complete electronic data capture forms at each household visit. Data captured includes, but is not limited to, the number of household members present (all intervention arms), content discussed (all intervention arms) and condition/use of the Kalingalinga buckets (Arms AB + A).

### Concomitant Interventions

There are no prohibited concomitant interventions during the trial. However, households will be asked if they have received any other health-related interventions before (at baseline) and during the study period (at endline).

### Patient and Public Involvement Statement

Households from similar peri-urban communities in Lusaka participated in the feasibility and pilot testing of intervention components. Eight households received the first five hygiene promotion visits, and were invited to evaluate content, comprehensibility and willingness to engage in the full programme. The programme was revised based on participant feedback.

Households identified the Kalingalinga bucket as the favoured handwashing facility out of several designs in a previous ranking exercises [26] and Trial of Improved Practices. A small pilot on the acceptability and use of *soapy water* with ten households was also conducted. Sixteen households participated in pilot baseline outcome assessments, including the three-hour structured observation.

Before study commencement, the research team will share the study protocol and ethical board approvals with the provincial and district health offices and request for permission to conduct the study in the pre-defined communities. Upon written approval, the research team will hold introductory meetings with the sister-in-charge (health facility manager) at community health facilities and sensitization meetings with community representatives (neighbourhood health committee members) who are the gate keepers responsible for health promotion.

Sensitization meetings will be conducted in the local language and include information on the study objective and aims, targeted study participants, and the procedures outlined in the protocol. Input will be requested on the best way to recruit participants. Following these meetings, attendees will disseminate information through established communication channels to ensure awareness of research activities.

### Outcomes

Primary Outcome:

*1. Handwashing with soap (HWWS) at all handwashing opportunities*

The primary outcome is the probability of HWWS within three minutes of any handwashing opportunity (Table 1) in the correct sequence (e.g., before or after the relevant opportunity).

Secondary outcomes:

*2. Handwashing with soap (HWWS) at each specific handwashing opportunity.*

a. The probability of HWWS before cooking (within 3-minutes before).
b. The probability of HWWS before eating (within 3-minutes before).
c. The probability of HWWS before feeding a child (within 3-minutes before).
d. The probability of HWWS before breastfeeding (within 3-minutes before).
e. The probability of HWWS after toilet use (within 3-minutes after).
f. The probability of HWWS after handling child/adult faeces or cleaning a child’s bottom (within 3-minutes after).
g. The probability of HWWS after contact with animals (within 3-minutes after).
h. The probability of HWWS after coughing/sneezing (within 3-minutes after).

*3. Hand-rinsing measured as a three-level outcome: no action, hand-rinsing or handwashing with soap (HWWS) at all handwashing opportunities.*

The probability of either HWWS, hand rinsing with water only, or taking no action within three minutes of any handwashing opportunity (Table 1) in the correct sequence (e.g., before or after the relevant opportunity).

Outcomes 1-3 will be measured through direct observation of individual handwashing behaviour over a three-hour observation period at baseline and endline (6-months after intervention delivery begins).

*4. Handwashing Facility (HWF) presence*

The probability that a household has a fully functional handwashing facility with water and soap present. This outcome will be measured at baseline and endline (6-months after intervention delivery begins).

*5. Handwashing knowledge and reported practice.*

**Knowledge:** The number of opportunities for handwashing with soap (Table 1) correctly specified in response to the question “When do you think it is important to wash hands with soap?”. **Reported practice:** The number of opportunities for handwashing with soap (Table 1) reported in response to the question “When did you wash your hands with soap yesterday?”

Both questions will be assessed through a survey of open-ended questions with pre-coded responses and multiple rounds of probing. This outcome will be measured at baseline and endline (6-months after intervention delivery begins) with one household member.

### Tertiary outcomes

*6. Diarrhoeal Illness*

The risk of self-reported diarrhoea at least once in the past 7 days (experiencing 3 or more loose stools in 24 hours).

*7. Respiratory Illness*

The risk of self-reported respiratory illness at least once in the past 7 days (experiencing coughing or difficulty breathing).

Outcomes 6 and 7 will be measured through a survey with primary respondents at baseline and endline (6-months after intervention delivery begins) and will be collected for all household members. Results will be stratified by age group.

### Participant Timeline

Data will be collected from each household over a period of six months. Households will be enrolled, randomised and have a baseline visit at Week 0. Due to the length of visit required for baseline measurements, and to ensure observations start on-time, households will be enrolled and consented a few days ahead of baseline data collection. Intervention delivery will begin two weeks later. The research coordinator will provide the intervention allocation list to intervention delivery teams on Friday the week before intervention delivery begins. Households that do not receive a baseline visit before this list is provided, will still receive the intervention. Endline data will be collected 6-months after intervention delivery begins. Measurement time points are detailed in Table 4 and Fig 5.

**Fig 5.**
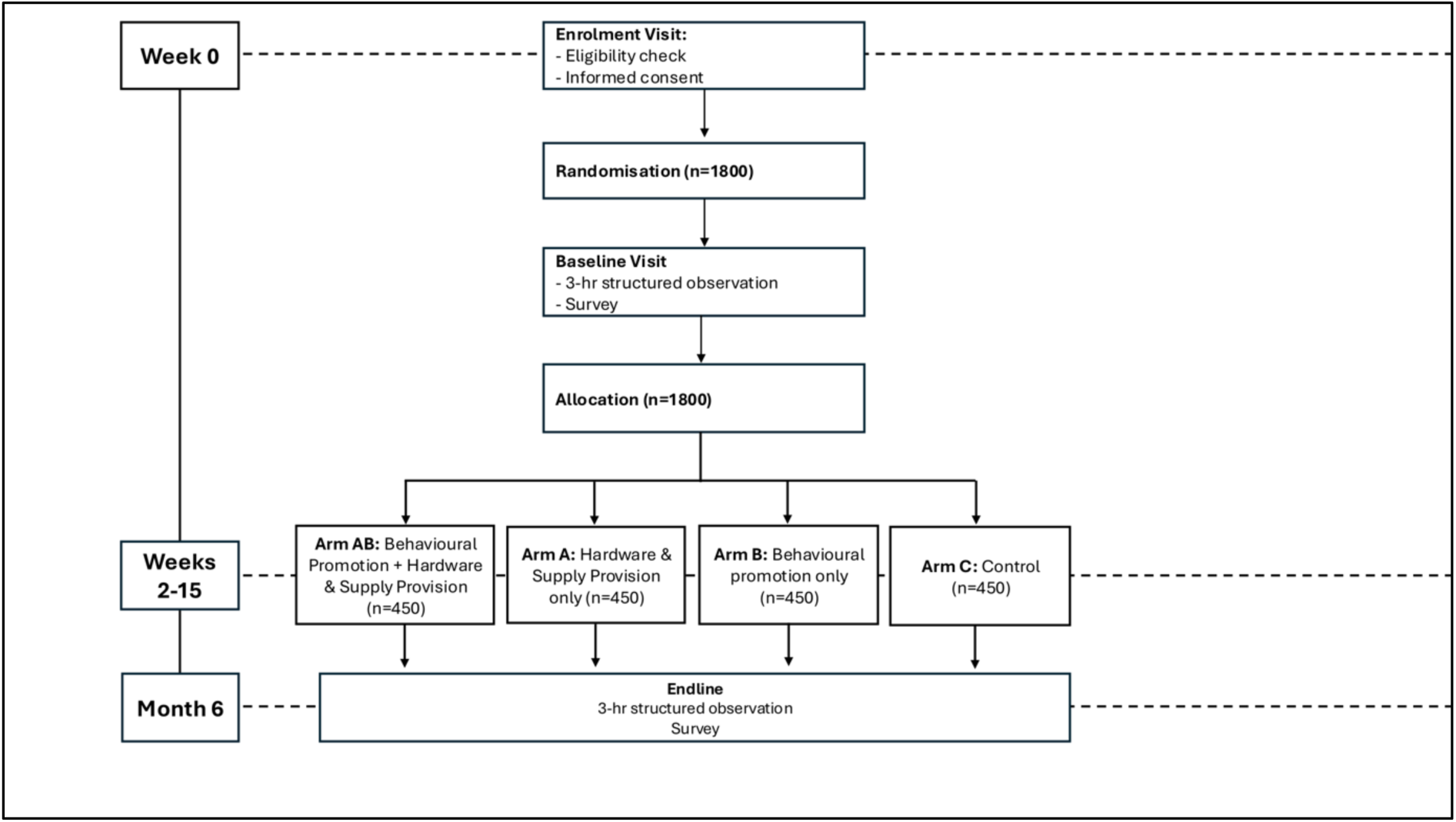
Study Design Flow Diagram.

**Table 4.** SPIRIT figure showing schedule of enrolment, intervention and assessments.

|  | STUDY PERIOD |  |  |  |  |  |  |  |
| --- | --- | --- | --- | --- | --- | --- | --- | --- |
|  | Enrolment | Intervention Visits |  |  |  |  |  | Endline |
|  |  | V1 | V2 | V3 | V4 | V5 | V6 / V2* |  |
| TIMEPOINT (WEEK) | 0 | 2 | 4 | 6 | 8 | 10 | 15 | 27 |
| **ENROLMENT:** |  |  |  |  |  |  |  |  |
| Eligibility screen | X |  |  |  |  |  |  |  |
| Informed consent | X |  |  |  |  |  |  |  |
| Randomization | X |  |  |  |  |  |  |  |
| **INTERVENTIONS:** |  |  |  |  |  |  |  |  |
| Hardware and supplies (AB + A) |  | X |  |  |  |  | X |  |
| Behavioural Promotion (AB + B) |  | X | X | X | X | X | X |  |
| **ASSESSMENTS** |  |  |  |  |  |  |  |  |
| Observation: handwashing behaviour (outcomes 1-3) | X |  |  |  |  |  |  | X |
| Survey: observed HWF presence (outcome 4) | X |  |  |  |  |  |  | X |
| Survey: Handwashing knowledge + reported practice (outcome 5) | X |  |  |  |  |  |  | X |
| Survey: Diarrhoea + respiratory illness (outcomes 6 + 7) | X |  |  |  |  |  |  | X |
| Intervention adherence Indicators |  | X | X | X | X | X | X |  |

375 \*Visit 6 for households receiving behavioural promotion (Arms AB + B) / Visit 2 for households receiving the hardware intervention (Arm A).

Household enrolment and baseline data collection started on 14^th^ March 2025 and was completed on 30^th^ May 2025. Endline is due to begin on 22^nd^ September 2025 and will be completed by 5^th^ December 2025.

### Sample Size

For the sample size calculation, we modelled the primary outcome as a set of repeated binary outcomes (multiple handwashing opportunities over the observation window for one person) assuming intra-individual correlation. We assumed that the prevalence of HWWS at a handwashing opportunity would be 25% in the control group [5]. We also assumed an average of six observed handwashing opportunities per household per 3hr observation period based on prior research in Malawi [31], and unpublished data in Zambia, and Uganda; and an intra-cluster correlation coefficient (ICC) of 0.3 based on unpublished data from Uganda. To account for five multiple comparisons (A vs C, B vs C, AB vs C, AB vs B, AB vs A), sample size calculations were adjusted by the Bonferroni method (family-wise α / 5) as a conservative assumption [32]. We calculated that a sample of 450 clusters (households) per arm would allow us to detect an absolute minimum difference in HWWS of 6.6% (proportion of opportunities in which hands were washed with soap) between any two groups 6-months after intervention delivery begins, with 80% power (α=0·01).

### Recruitment

A sampling interval for household enrolment was calculated using ward-level population estimates from the central statistics office (CSO) in Zambia, assuming a household size of five (based on urban estimates from the Zambia Demographic and Health Survey) [33]. We assumed that any inaccuracies would be consistent across wards. The total sample was distributed proportionally across wards, and the interval (*k)* was calculated as *k* = N / *n*, where N is the estimated number of households in the ward and *n* is the target sample size for that ward. This gave a preliminary sampling interval of 49 households (Table 5). Assuming an average of four households per plot [34], data collectors will visit every 12^th^ plot.

**Table 5.** Sampling Interval Calculation.

| Ward | Population (p) | Estimated number of households per ward ($N = p / 5$) | Proportion ($p = N / \text{total } N$) | Sample Size per ward ($n = p \times 1800$) | Sampling Interval ($k = N / n$) |
| --- | --- | --- | --- | --- | --- |
| Chainda | 38,393 | 7679 | 0.09 | 157 | 49 |
| Mpulungu | 57,596 | 11519 | 0.13 | 235 | 49 |
| Chawama | 70,181 | 14036 | 0.16 | 286 | 49 |
| Mtendere | 106,128 | 21226 | 0.24 | 433 | 49 |
| Kanyama | 169,253 | 33851 | 0.38 | 690 | 49 |

Sampling and enrolment will be concurrent and conducted by members of the study data collection team. Data collectors will be distributed evenly across the zones within wards. Teams will proceed to the first plot or free-standing house along their sampling interval. In a plot, the team will select one household at random using a random number generator available on data collection tablets. If a household is empty at the time of enrolment, two additional attempts will be made to contact that households – once on the same day and once the next day. If households are unreachable, or not eligible, the next household in the plot or along the street will be approached.

### Allocation

#### Sequence generation

Randomisation will be stratified by ward and use a permuted block design to reduce predictability and ensure approximate balance among the four trial arms over the 11-week enrolment period. Prior to enrolment, the trial statistician (SB), with no prior knowledge of the participants, households or communities, will use randomly permuted blocks (sizes 4, 8 and 12) to generate a randomisation sequence for each ward with its length corresponding to the expected enrolment for the respective ward.

#### Implementation and concealment mechanism

The random sequences will be embedded into a password-protected online dashboard (Rshiny app) integrated with the electronic data capture system (ODK), which will be used to assign households to treatments automatically and sequentially as they are enrolled, according to fixed characteristics of ward, and precise (millisecond) timestamp of when enrolment data were submitted to the ODK server. The original randomisation sequence including details of blocking will be kept securely, and the resulting allocations will be visible only to study coordinators and made available only to those responsible for providing or preparing the interventions and not to those enrolling households or collecting outcome data at any point.

#### Blinding

Due to the nature of the intervention, it will not be possible to blind study participants to intervention allocation. This is an inherent challenge in behaviour change and environmental health research and is an acknowledged limitation. Data collection teams – who are separate from the team who will deliver the intervention - will be unaware of study arm allocation.

However, it is possible they will notice the presence of hardware whilst conducting structured observations at endline. Investigators aware of study objectives and hypotheses will not participate in data collection or intervention delivery. Investigators conducting data analysis will be blinded to intervention allocation.

### Data Collection Methods

#### Structured Observations

Outcomes will be measured using three-hour structured observations at baseline and endline (6-months after intervention delivery begins). One eligible adult from each household will be observed at each timepoint. If the child under 5 is present, the adult (18 years or older) most responsible for caring for the child at the time of the visit will be selected for observation. If the child under 5 is not present, the adult (18 years or older) most responsible for childcare will be selected for observation. If such an individual is not present, another randomly selected household member (18+) will be observed instead.

A list of key handwashing behaviours we will observe with definitions is provided in Table 6. Other behaviours we will observe include handwashing opportunities (Table 1) and hygiene management activities (refilling handwashing facility, moving soap near to handwashing facility, moving handwashing facility, preparing soap and disposing of grey water from handwashing facility).

**Table 6.** Handwashing behaviour definitions.

| Outcome Category | Definition |
| --- | --- |
| **Hands washed with soap and water (HWWS)** | Both hands washed under running water (e.g., from a tap or jug/cup) with soap. |
| **Hands rinsed (water only)** | Both hands washed under running water (e.g., from tap or jug/cup) with no soap.<br>Dipping hands in a basin with water to clean hands (with or without soap).<br>Both hands have contact with water (with or without soap), but other activities are being performed other than handwashing (e.g., doing laundry or washing dishes). |
| **No action** | Only one hand washed.<br>No action. |

Observation data will be captured using electronic data capture forms on Open Data Kit (ODK). Forms will be configured to automatically record a timestamp when a data collector documents a behaviour. To determine whether hands were washed at a handwashing opportunity, we will allow a three-minute timeframe between the opportunity and the handwashing behaviour. This is to account for the time taken to fetch soap from the household when handwashing should occur after a handwashing opportunity [15]. Past three minutes, or if the household member engages in another activity between the handwashing opportunity and handwashing, the event will be classified as “no action”. If the household member does not wash their hands at a handwashing opportunity but performs an activity involving the use of soap (e.g., laundry or washing dishes), within three minutes before/after the key occasion, this will be recorded as “contact with soap and water but not HWWS” but categorised as hand rinsing.

Structured observations will be conducted for three hours, between 7am-4pm. Our prior work in both this context and other settings has identified two critical windows where we anticipate higher numbers of hand hygiene opportunities – 7 am to 10 am (specifically around morning mealtimes and domestic chores) and 1 pm to 4 pm (specifically around mid/day evening meal preparation and the return of individuals to the home after going to the market or employment). We anticipate that due to data collection logistics (one observer completing two observations per day), approximately half the sample will be observed at each timepoint (early morning vs afternoon observation).

Data collection staff will position themselves in a place where most of the household – specifically food preparation areas and toilet infrastructure - are in view. Care will be taken to ensure the observer’s position and presence does not disturb the daily organisation and activities of the household. Indoor observations will occur only in shared or common spaces within the home. Based on previous research in the area, the behaviours of interest (food preparation, childcare, etc.) occur in the areas immediately outside of the house.

While there are known limitations to structured observations, it remains the gold standard for the measurement of routine behaviours in the domestic context [35, 36]. Other measures of behaviour using self-reported practice have very low reliability and poor association with actual behaviours [37, 38].

### Household Surveys

A household survey will be completed after the observation at baseline and endline. At baseline, the survey will collect information on household demographics – including assets, income and household size – as well as access to water, sanitation, and hygiene (WASH) services, and experiences of water insecurity. The survey will also collect demographic information on each household member as well as self-reported cases of diarrhoea or respiratory illness in the past week. At endline, the survey will reassess household WASH access and update selected demographic information, including diarrhoea and respiratory illness among household members. Demographic data for the specific household member observed at endline will also be re-collected. An additional module at endline will assess exposure to the intervention as a measure of contamination.

The household survey completed at baseline and endline will include a health behaviour module. Participants will be asked to self-report when they washed hands yesterday, then will be asked when they believe HWWS is important. To reduce potential bias associated with focusing solely on handwashing, the survey will also incorporate additional modules on child immunization, child feeding, and child engagement / play. These modules are adapted from UNICEF’s MICS6 questionnaire for children under five. All surveys will be completed after observations to avoid biasing behaviour. Surveys will be completed using electronic data capture forms on Open Data Kit (ODK).

### Data Quality

Data collection teams will undergo a week-long training before study implementation, focusing on data collection methods. The training will include practical sessions where team members role-play household visits and practice translating and delivering questions in different languages (English, Nyanja and Bemba). Additionally, they will conduct practice observations using videos of domestic settings. To reinforce learning, team members will complete a test observation using a video, with feedback provided on submitted data.

### Process Data Collection

Process data will be collected by intervention delivery workers during their intervention visits using electronic data capture forms on ODK. Data captured includes, but is not limited to, the number of household members present (all arms), reported content discussed (all arms), and condition/use of the Kalingalinga buckets (Arms AB + A). Households will be asked if they have received any other health-related interventions before (at baseline) and during the study period (at endline).

### Retention

Participants will receive study booklets where the dates of their upcoming visits will be recorded. Separate booklets will be maintained for intervention delivery workers and data collection teams (who conduct baseline and endline assessments) to prevent unblinding. Before endline data collection, intervention delivery workers will remove intervention booklets to ensure data collectors remain blinded to household allocation.

### Data Management

All households will be given a QR code corresponding to a unique alphanumeric code which will be used to link household data through the various stages of the study. QR codes will be kept with the household and attached to signed consent forms. Electronic data entry forms will be built using ODK. All data will be anonymized prior to sharing with LSHTM. Data will be transferred between LSHTM and CIDRZ using online secure file sharing systems. De-identified data will be sent weekly for monitoring purposes.

Participants will have the right to gain access to, change, or oppose the collection of any data during the interviews or observations. Participants have the right to withdraw altogether from the study, without providing any justification, by contacting any of the investigators listed on their participation information sheet by mail or phone. Data and all appropriate documentation will be stored for a minimum of 5 years after the completion of the study, including the follow-up period.

### Statistical analysis

The analysis and reporting of results will be in line with the Consolidated Standards of Reporting Trials (CONSORT) Statement for randomised controlled trials [39] and its extension for multi-arm trials [40]. Statistical analysis will be conducted using R (V4.5.0). Full details of all models and analytic procedures, including covariates to be included in the adjusted models, additional subgroup analyses, and sensitivity analyses, will be set out *a priori* in a Statistical Analysis Plan that will be prepared nearer the data analysis stage and published prior to any analysis of endline data.

Our analysis will concern five pairwise comparisons:

1. Behavioural promotion against control (B vs. C)
2. Hardware and supplies provision against control (A vs. C)
3. Behavioural promotion + hardware and supplies provision against control (AB vs C)
4. Behavioural promotion + hardware and supplies provision against behavioural promotion alone (AB vs B)
5. Behavioural promotion + hardware and supplies provision against hardware and supplies provision alone (AB vs A).

The remaining comparison (A vs. B) may be analysed as an exploratory analysis if there is evidence of effectiveness of either intervention.

A two-stage approach to analysis of each outcome across the multiple trial arms will be used, as described in Korn et al. [41], involving first comparing individual intervention arms A, B, and AB to arm C (pairwise comparisons 1-3 above), followed by comparing AB (the combination of A and B) to the best of A, B, and C (comparison 3, 4, or 5) if AB is found to be superior to C in the first step (the null hypothesis for comparison 3 is rejected). This approach allows us to retain a larger statistical power than testing the five comparisons simultaneously as a different nominal significance threshold is used for each step, while maintaining the same overall family-wise type I error rate.

Descriptive statistics of demographic and outcome measures at baseline will be reported by trial arm to determine any imbalances. The primary outcome is HWWS behaviour (binary variable: yes or no) at any key handwashing opportunity at the 6-month follow-up, with potentially multiple opportunities per household during the observation period. Random-effects logistic regression accounting for clustering by household will be used to compare the probability of HWWS at a key handwashing opportunity between arms for each pairwise comparison. Margins will be used post-estimation to convert odds ratios to predicted probabilities (risk) for interpretation. Sensitivity to clustering at other levels (such as the data collector conducting observations) may be explored. Unadjusted and adjusted results will be presented for all analyses. Covariates in adjusted analyses will be specified a priori and will include the strata used in randomisation (ward location).

Bias due to missing data will be investigated, and participants who were missing at baseline or endline, or who withdrew from the trial between baseline and endline will be compared to those with complete data. We will also explore the extent of missingness due to zero handwashing opportunities during the observation period. If required, sensitivity of the results to missing data will be investigated using different approaches for handling missing data, such as adjusting for predictors of missingness that are related to the outcome. Further models allowing for missing not at random mechanism may also be considered.

Secondary outcomes 2a-g (HWWS at each specific handwashing opportunity) will be compared in the same way as the primary outcome, but as there will likely be fewer events than the primary outcome per observation, analysis will proceed given data availability. Other secondary outcomes will be compared between the intervention arms using appropriate regression models taking account of clustering where necessary. Distributional assumptions will be verified as appropriate.

To assess possible heterogeneity of the intervention effects, subgroup analyses will be undertaken as secondary analyses, including the planned subgroup of water insecurity experienced by the household measured using the Household Water Insecurity Experiences Scale [42].

As the primary aim of the trial is to evaluate the effectiveness of offering the intervention/s, the intention-to-treat principle will be used in the primary analysis of all outcomes. Endline outcome data will still be collected for households that did not receive the intervention as per protocol, and these households will be included in the primary analysis according to their original group, even if they received no intervention, partial intervention or the wrong intervention. Per-protocol analyses may be conducted as exploratory analyses, but these will be interpreted with caution. As the primary analysis will be based on differences between arms at endline, households who were missing at endline, or who withdrew from the trial between baseline and endline, will not be included in the primary analysis.

### Data Monitoring & Auditing

As data are only collected at two time points, and there is no interim analysis, a Data Monitoring Committee is not needed. The study may be subject to audit by the London School of Hygiene & Tropical Medicine under their remit as sponsor and other regulatory bodies to ensure adherence to GCP.

## ETHICS AND DISSEMINATION

### Research Ethics Approval

Ethical approval was obtained from London School of Hygiene and Tropical Medicine, Research Ethics Committee, London, UK (Ref: 31387), the University of Zambia Biomedical Research Ethics Committee (Ref: UNZABREC 5834-2024) and the Zambia National Health Research Authority (NHRA-1823/23/12/2024).

### Protocol amendments

Important protocol modifications will be submitted to relevant ethics and regulatory committees mentioned above and updated in the trial registries (clinicaltrials.gov). Participants already enrolled in the study will be re-consented on updated consent forms.

### Consent

There are two levels to consent: (1) Consent to participate in the study intervention or control and (2) consent for data collection. Procedures are outlined for each of these below.

All study information will be read in a language participants understand (ensuring a witness is present if the participant is illiterate). Information sheets and consent forms will be translated into the local language (Bemba and Nyanja) by the local research team. Questions on any aspect of the study will be invited from the participant. The study team will check for understanding of the information provided using a basic five question knowledge assessment questionnaire. If not understanding, the forms should be reviewed again with the participant.

If the participant understands the forms and is willing to participate, the team will ask they provide a written signature on the consent form. If the participant is illiterate, the participant should leave a thumbprint or X on their signature line, and the witness will sign in the appropriate part of the consent form.

### Consent to participate in the study

Informed consent for household study participation will be obtained at enrolment by the data collection team. The team will explain the purposes of the study in broad terms to households that meet eligibility requirements and invite them to learn more about the study. If they agree, the team will read the Study Participant Information Sheet (PIS) to available members of the household, or ask the participant to read themselves, and give the household time to ask questions. The Study PIS will explain randomization procedures, anticipated interaction / time with the households, participant rights, and data collection procedures. The Study PIS will explain that participating households have the right to end involvement in the study at any point without penalty.

The team will obtain written informed consent from the head of household or a nominated representative for household participation in the study, on behalf of all household members. If the head of household is not present, we will obtain verbal consent from the head household for a nominated representative to provide written consent for household study participation.

Additional signatures will be collected from all adults in the households present at the time of enrolment.

### Consent for data collection

A separate informed consent process will be completed for members of participating households that participate in specific data collection activities. Consent will be obtained at both baseline and endline, with the household member being observed. The data collection PIS will provide a brief overview of the specific data collection activities, explain the participants rights, and that they are free to terminate data collection at any moment. Participants will also be informed that they can choose to have their data destroyed or withdrawn. Written informed consent will be collected for the person who is participating in data collection.

### Confidentiality

Any participants’ identifiable data will be stored securely, and their confidentiality protected. All electronic data will be stored on a secure, password protected server. Only named individuals on the protocol will have access to the data. All mobile devices used for data collection will be encrypted and password protected. Any non-electronic documents will be stored in a locked filing cabinet, with access limited to field supervisors and study team members.

### Data Access

De-identified participant data, data dictionary, and analytical code used to process the data will be posted on a public repository for third party use. To completely anonymise the data prior to uploading it onto an open data site, any information that holds the potential to identify the respondents will be redacted. Explicit consent to put de-identified data on a public repository will be sought from trial participants. Only named investigators will have access to the full trial dataset.

### Ancillary and post-trial care

At the end of the study period, the most effective combination of interventions will be delivered to households in the control group. London School of Hygiene & Tropical Medicine holds Clinical Trial/Non-Negligent Harm Insurance and Medical Malpractice Insurance policies which apply to this trial. Due to the nature of the study, we anticipate the risk of harm to be very low to participants. Participants are informed to contact the local principal investigator, or the research ethics committee/research governance and integrity office, if they experience harm or injury because of taking part in the study.

### Dissemination policy

All results from the study will be submitted for publication in peer-reviewed journals. All code and fully anonymised data will be posted on a public repository for third party use. The final list and order of authors will reflect each researcher’s contribution and will comply with the Vancouver recommendations. Results may also be communicated through additional formats, such as a plain language summary.

## FOOTNOTES

### Contributors

RD is the grant holder. RD and JC were involved in study conception. RD, JC, AS, KD, KK and SB took part in weekly meetings in which the design of the study was decided and data collection planned. KD and KK planned data collection and intervention delivery activities. SB provided statistical expertise in clinical trial design, including developing the randomisation sequence. All authors contributed to refinement of the study protocol draft and approved the final manuscript.

### Trial Sponsor

LSHTM are acting as the research sponsor (ref: 2024-KEP-1164). The sponsor will have no direct involvement in the trial, including study design, conduct, data analysis and interpretation, manuscript writing or review and results dissemination.

## SUPPLEMENTAL MATERIAL

**S1 Text. Behavioural Promotion Intervention Details**

**S2 Table. SPIRIT 2025 checklist of items to address in a randomized trial protocol**

**S3 Table. TIDieR-WASH Checklist**

## Data Availability

N/A - protocol manuscript.

## Hygiene Promotion: Ulupwa (The Family Intervention)

### Intervention overview

*Ulupwa* employs interactive African storytelling and visual aids to promote handwashing with soap in peri-urban communities in Lusaka, Zambia. Participants associate themselves with character profiles – Nevers, Sami, or Always – based on their handwashing habits, and the stories of these characters highlight causes, barriers, consequences, and solutions for handwashing behaviour. Storytellers use cards to illustrate their story, making the intervention engaging while teaching essential hygiene practices.

The intervention progresses through visits focusing on different stages: identifying behaviours, understanding disease risks, encouraging practical solutions and promoting practice. The penultimate stage of the intervention focuses on reflection and commitment, encouraging participants to assess their behaviours and pledge to adopt improved hygiene practices. A final follow-up visit allows participants to re-cover any information and reflect further on the intervention. Through motivational messaging and nurturing themes, the intervention aims to not only educate but foster long-term behavioural change.

### Playing cards

The key prop for this intervention is a set of visual ‘playing’ cards that illustrate the characters, handwashing barriers and other related concepts. They help make the intervention accessible and engaging. Examples of the cards are provided at the end of this document.

The pack of cards illustrate the following:

- Nevers handwashing tick cards (x4 per pack)
- Sami handwashing tick cards (x4 per pack)
- Always handwashing tick cards (x4 per pack)
- Nevers character picture card (x1 per pack)
- Sami character picture card (x1 per pack)
- Always character picture card (x1 per pack)
- Community cards (x5 per pack)
- Handwashing barriers cards (set of x10 cards per pack)

`o` Nothing to wash hands with
`o` Don’t have money
`o` Broken basin
`o` Can’t reach of find the soap
`o` Some jobs are only for women
`o` There is no one to teach me
`o` There is no community support
`o` Too busy doing other things
`o` Is handwashing important?
`o` Any other reason?
- How we get sick cards (set of x2 cards per pack)

`o` Germs on hands
`o` Stomach pain
- Handwashing junctures cards (set of x7 cards per pack)

`o` Wash hands before cooking
`o` Wash hands before eating
`o` Wash hands before feeding children
`o` Wash hands after toilet use
`o` Wash hands after cleaning children
`o` Wash hands after handling animals
`o` Wash hands after coughing
- Handwashing resources cards (set of x6 cards per pack)

`o` Water container
`o` Soap
`o` Bucket
`o` Jug
`o` Covered bucket with tap
`o` Basin
- Handwashing reminder (Don’t forget!) cards (x6 per pack)
- Pledge cards (x9 per pack)
- Storyteller ‘business’ cards (x1 per pack)

### Other resources

a. Branded plastic water bottle (with brand sticker)
b. Storytellers
c. Story scripts

## Visit 1: Greeting

### Aim of the visit

To establish identities, including handwashing behaviours and perceived barriers.

### Intended outcomes

- Establishes a connection/builds trust with the family for future communication.
- Create an environment where resources can lead to transformative conversations.
- Understand the handwashing behaviours of each family member.
- Form an appreciation of perceived handwashing barriers.

### Supporting Mechanism

*ubuntu* (a cousin to *a*iliation*) – the tactics of ubuntu are about sharing knowledge/information about others, reducing threats and creating associations. It promotes an environment for transformative conversations that improve understanding and promote harmony.

### Resources

- Nevers handwashing tick cards (x4 per pack)
- Sami handwashing tick cards (x4 per pack)
- Always handwashing tick cards (x4 per pack)
- Handwashing barriers cards (set of x10 cards per pack)

### Section 1: Introductions

The purpose is to explain the visit, introduce the topic of handwashing and establish the context of the intervention. Storytellers introduce themselves and invite the members of the household to do the same. The conversation should flow using the following semi-structured guide:

1. My name is […] I am a Storyteller from Ulupwa, a project to help families. My home is in […]. I am pleased to meet you and would like to share some stories with you. May I please know your names?
2. Family introduces themselves and may give details about their home situation. If not, then prompt with the following:

a. Before I begin with my stories, I would like to understand your home set up. I have some questions e.g.:

i. Where is your toilet?
ii. Where do you get your water?
iii. Do you have any jugs or basins for handwashing?

### Section 2: Identifying barriers

The purpose of this session is to explore handwashing barriers by introducing the three main characters and discussing the barriers to handwashing they may face.

There are three main characters:

1. **Nevers** doesn’t understand the importance of handwashing and never washes hands, providing many excuses for their behaviour.
2. **Sami**, short for Sometimes, knows handwashing is important but often forgets to wash hands. Sami only occasionally washes hands.
3. **Always** washes hands properly with soap and water all the time.

***Storytellers*** present the family with (gender and age) neutral character tick cards. The cards have 12 unticked boxes to represent the times hands could be washed throughout the day such as before cooking, before eating, before feeding, and after toilet use, at different times of the day (morning, noon and evening).

In front of the family, and while describing the three main characters and their handwashing behaviours, the story-teller ticks all of the boxes for Always, some for Sami and none for Nevers.

***Storyteller*** discusses the barriers each character may face:

1. How does Nevers end up with no ticks?
2. How does Sami end up with some ticks?
3. What are the differences between the barriers?

As prompts, present the handwashing barriers cards.

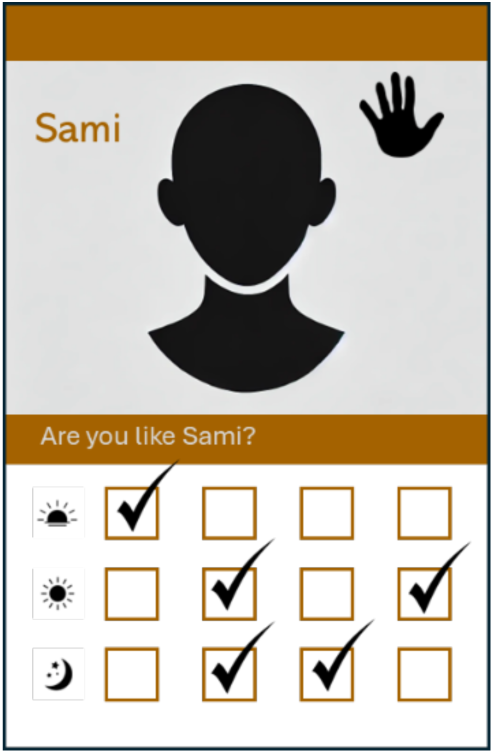

### Section 3: Selecting a character profile

The purpose is to identify the handwashing behaviours through asking the family to choose the character profile that is closest to their behaviour.

***Storyteller*** asks each family member is asked to pick a card that best represents their behaviours.

The facilitators and other participants may challenge these choices. The participants will keep this card as a reminder of their choices until the next visit.

## Visit 2: Understanding disease risks

### Aim of the visit

Build awareness around the disease risks associated with inadequate handwashing with soap and provide information on when to wash hands.

### Intended outcomes

Improved knowledge on disease risks and critical junctures for handwashing.

### Supporting Mechanism

*Curiosity* – the tactics of curiosity are to observe, try things out, seek to understand/ explain, manipulate (novel) objects and consume symbolic information.

### Resources

- Nevers character picture card (x1 per pack)
- How we get sick cards (set of x2 cards per pack)
- Handwashing junctures cards (set of x7 cards per pack)

### Section 1: Introductions

***Storyteller*** takes time to greet everyone then recaps the last visit using the following prompt:

- What did we learn from the last meeting and what have we done differently?

### Section 2: Nevers’ story

***Storyteller*** tells Nevers’ story. The story explains who Nevers is and how Nevers goes through a day without washing hands. The story mentions how this behaviour has now put a vulnerable family member, the grandmother, at risk of getting sick. Sami and Nevers father intervened. As part of this story, test hands for cleanliness using the Protect Hand: Protect Health app.

### Section 3: Disease risk + critical junctures

***Storyteller*** leads a discussion on:

- how germs cause illness (using how do we get sick cards as prompts)
- when we should wash hands (using handwashing juncture cards as prompts).

## Visit 3: Encouraging Practical Solutions

### Aim of the visit

To introduce nudges or other environmental cues designed to prompt handwashing with soap.

### Intended outcomes

To ensure each house has an appropriately located handwashing facility.

### Supporting Mechanism

*Play* – the tactics of play are practice skills, observe and imitate behaviours.

### Resources

- Sami character picture card (x1 per pack)
- Handwashing resources cards (set of x6 cards per pack)
- Handwashing reminder (Don’t forget!) cards (x6 per pack)
- Branded plastic water bottle (with brand sticker)

### Section 1: Introductions

*What did we learn from the last meeting and what have we done di*erently?*

### Section 2: Sami’s Story

***Storyteller*** tells Sami’s story. Sami knows that handwashing is important but sometimes forgets. Sami realises the need for a handwashing facility that is in an appropriate area for handwashing.

### Section 3: HWF Discussion

- ***Storyteller*** asks household to discuss which items they could use for handwashing in their home (prompt using handwashing resources cards).
- ***Storyteller*** asks household to discuss where a HWF could be placed in their home.
- ***Handwashing reminder cards*** *distributed, and household asked to stick them near places they need reminding to wash hands (e.g., where they eat or use the toilet)*.

### Section 4: Soapy water

- ***Storyteller*** shows household how to make soapy water – provide 500mL plastic bottle (with intervention brand sticker).

## Visit 4: Promoting practice

### Aim of the visit

To motivate households to take responsibility for handwashing, focused on protecting household members and the community from disease.

### Intended outcomes

Increased handwashing at the appropriate times.

### Supporting Mechanism

*Nurture* – the tactics of nurture are to give resources, protect from dangers, provide opportunities for play, transmit status and nepotism.

### Resources

- Always character picture card (x1 per pack)
- Community cards (x5 per pack)

### Section 1: Introductions

*What did we learn from the last meeting and what have we done di*erently?*

### Section 2: Always’ story

This visit tells the story of Always, Sami and Nevers’ cousin, who always washes hands with soap and water and wants to ensure all family members take responsibility for maintaining safe handwashing practices.

### Section 3: Discussing Responsibility

***Storyteller*** hands out community cards and asks:

- “How can you do the same as Always to take responsibility for yourselves and each other?”

## Visit 5: Reflection (Self-assessment and pledges)

### Aim of the visit

To reinforce key messages delivered across the intervention period and aims to encourage participants to re-assess their behaviours and pledge to adopt improved hygiene practices.

### Intended outcomes

A commitment to continued, appropriate handwashing behaviours.

### Supporting Mechanism

*Curiosity* – the tactics of curiosity are to display awareness of social error and confidence, and to seek recognition.

### Resources

- Pledge cards (x9 per pack)

### Section 1: Introductions

*What did we learn from the last meeting and what have we done di*erently?*

### Section 2: Reflection

***Storyteller*** asks household to reflect by asking several questions to everyone in the room:

1. Describe the whole storytelling experience.
2. What did you learn about handwashing?
3. Has your handwashing behaviour improved?
4. Do you now have a family roster?
5. Which character do you associated with now?

- Give a card with a handwashing behaviour card (if there is a difference).

### Section 3: Pledges

***Storyteller*** asks each member individually, if they are prepared to pledge. Guide them through the pledge and give them a pledge card to keep.

## Visit 6: Recap

### Aim of the visit

To enable households to reflect on the intervention as a whole and recap any information participants would like to re-cover.

### Section 1: Introductions

*What did we learn from the last meeting and what have we done di*erently?*

### Section 2: Recap

***Storyteller*** asks household to reflect on the below questions and re-covers information where households have forgotten.

1. Can you remember what diseases you can get from washing hands?
2. Can you remember when we should wash hands?
3. Have you set-up a handwashing facility?
4. Do you still have the handwashing reminder cards?
5. Can you remember how to make soapy water? [If no, remind how to make soapy water]

## Nevers’ Story (Visit 2)

In Lusaka, there lived a cheeky boy named Nevers. Their house was [*insert description of the house here*]. It had a [*describe toilet at the premises*] and [*describe water situation at the house*].

Nevers was known for two things: his love of being naughty and completely refusing to wash his hands. From morning to night, Nevers went about his day, purposefully ignoring soap and water which was [insert location of soap and water at the premises being visited].

It started early in the morning when Nevers woke up with a loud yawn. He dashed outside to the bush do to a poo, using leaves for toilet paper. He picks up a bucket to get some water to wash his hands, but instead of washing his hands, Nevers drops the bucket somewhere along the way back, and rubs his hands on his shorts, while saying to himself “Why waste water anyway when they will dry? Besides, the water well is too far. The guys are waiting for me.”

He runs back into the house and wakes up his sister, Sami by throwing a pillow at her. “Get up, lazy bones! It’s time to live,” he said. Sami who is much older and now working, unwillingly gets up. Nevers is her alarm clock, and when he is early, she is early. When he is late. She is late. This time, she was late because she stayed up talking to her father.

With a shock, Sami woke up and rushed out to get the bucket to wash her hands and face. She sees empty boxes for the produce her mother sells, but she could not find the bucket…

Meanwhile, as usual, Nevers is nowhere to be seen. He had gone to collect his cousin, Always to play with friends. Always lived not too far away and his family reared chickens. They would play with an old plastic football through the nearby dusty playground. Nevers was the goalkeeper. He’d pick up the ball with his hands, which was often being kicked into the manure near their playground. Every time the other team had a penalty, he would first spit in his hands and rub them for good luck, before saving some hot shots. Always often told him he had a “smell”, and along with their other friends, would wrinkle their noses. Whenever Always asked, “Nevers, what’s that smell?” He would always reply with a broad smile, “It’s the chickens, it’s your chickens. They’re everywhere!”

Lunchtime arrived, and Nevers rushed home, starving. He noticed his grandmother having some nshima and, without washing his hands, grabbed a juicy bone with meat from her plate and ate all the meat from it. He laughed as he swallowed it down, put the bone back in the plate and grabbed the rest of the nshima. He loved to irritate his grandmother, and she enjoyed his behaviours.

He then sat there pondering his next move, while licking his fingers clean. His sister, Sami, gagged. “Nevers! You didn’t wash your hands!” she exclaimed. “It’s flavouring,” Nevers joked, licking his lips. “Extra spice!”

As he had finished his grandmother’s food thought he would get her some more. He really liked her, after all. He reached over to the pot and dished some food into a plate that he would share with her. His father noticed, and with a loud voice shouted, “Nevers, you have not washed your hands!”

Nevers’ hands were not clean and full of germs. If he shared his plate with his grandmother she could get ill. But, before carrying on with the story, let’s see how clean your hands are.

### App Exercise

Address the youngest person in the family and ask to test their hands for cleanliness. With everyone watching, use the **Protect Hand: Protect Health app** on phones.

Place the phone screen up as if scanning the hands - this will reveal that they have germs on their hands as they are not 100% clean. Tell them the % cleanliness reported in the app.

### Story continued

Nevers’ father continued. “And there is big stink in the air,” he said. Everyone turned to look at Nevers, who looked like he did not know aswell. “It’s the firewood,” he offered, waving his oily hands.

“Go and wash your hands,” his father demanded strongly. “Your grandmother can get sick because of you.” Nevers seemed to get the message now. He still did not know how you get sick from not washing your hands. Let us help him understand.

## Sami’s Story (Visit 3)

Sami, Never’s sister, loved a good chat with her father, often staying up late exchanging stories. If it weren’t for Never waking her up every morning, she would probably be late to work every day. The previous night, Sami was up late listening to her father tell stories about how some families never seemed to get coughs and their children were always at school, and how some children were always absent from school due to illness. That was a late night for everyone, including Never.

The next morning, disaster struck - Never was late waking up, and so he was late waking up Sami. She felt a bang from a pillow hitting her face – that is how Nevers woke her up. With a shock, she woke up, glanced at the time, and let out a short scream. “Ah! I’m late! Nevers why did you not wake me up earlier!”

She threw on some clothes and rushed to the toilet. She finished and ran outside to grab the bucket of water to wash her face and hands. But it was missing from its usual place. “Ah! Who has taken this bucket again?” she complained, looking around the yard. With no time to search, she wiped her face with a dry towel and rushed oE to work.

Sami finished work by lunchtime, so she comes home, hungry. She goes into the kitchen and cuts herself a thick slice of bread. She takes a bite, then remembers she has not washed her hands. “Aaargh, it’s too late!” she says. This is the second time she has not washed her hands that day.

Sami realises she needs to sort out the bucket issue and get something to remind her and her family, especially Nevers, to wash their hands. She plans on buying a basin and placing it just outside the toilet so that she can find it and is reminded to wash her hands when she uses the toilet. She will put her family name on it, to make sure others know it belongs to them.

But, besides the basin, Sami will need other things too.

## Always’ Story (Visit 4)

Always, Nevers’ cousin and friend, had heard all about the activity at Never’s house - how Never puts his family at risk and how Sami had set up a new handwashing station with reminder cards on their walls, because she sometimes forgets to wash her hands. Always wasn’t one to take hygiene lightly. His family kept lots of chickens, and he knew firsthand how dirty hands could get after handling animals. The problem? His family only had one handwashing facility, and with so many people and animals living in the same place, the water bucket by the toilet was empty when he needed it most.

Determined to make things better, Always moved into action. He convinced his mother that they needed a timetable for ensuring the bucket always has water and there is soap nearby. His mother agreed, giving her nod of approval to the list Always had developed. “Now, no excuses!” he declared, pointing at it proudly. He stuck the piece of paper on the toilet door.

The benefits were almost immediate. His mother, usually worried about stomach upsets in the family, noticed fewer complaints and more handwashing before meals. His little brother, who usually had a suspicious layer of dirt on his face, started looking…well, slightly cleaner. Even his father, who had complained about “too much water being wasted,” admitted he liked having clean hands after working and before eating. He was proud of his clean hands and the cleanliness of the family.

One evening, as they gathered around for supper, Always raised an important question: “You know what happened at Nevers’ house. That was serious. How can we keep protecting our family’s health?” His father, rubbing his freshly washed hands together, nodded. “It’s all our responsibility,” he said. “If we want to stay healthy, we must keep up these duties. If you want Nevers to stay healthy, you must both wash your hands after playing football. You reduce the chances of getting diarrhoea and missing school and having fun.” His mother added, “We should also remind visitors to wash their hands when they come over,” she said. “Amake Nevers came visited earlier today I was very happy when she complimented us on having a working Kalingalinga bucket with water and soap.”

And so, it became a family effort. No one had to be forced; they all just did it. The chickens? Well, they were still chickens – as messy and as noisy as ever. But at least now, their human caretakers were keeping things clean. Always took great pride in knowing that he was not just avoiding sickness, he was making life better for everyone. And Nevers? Always made sure Nevers did not go home without washing his hands ever again.

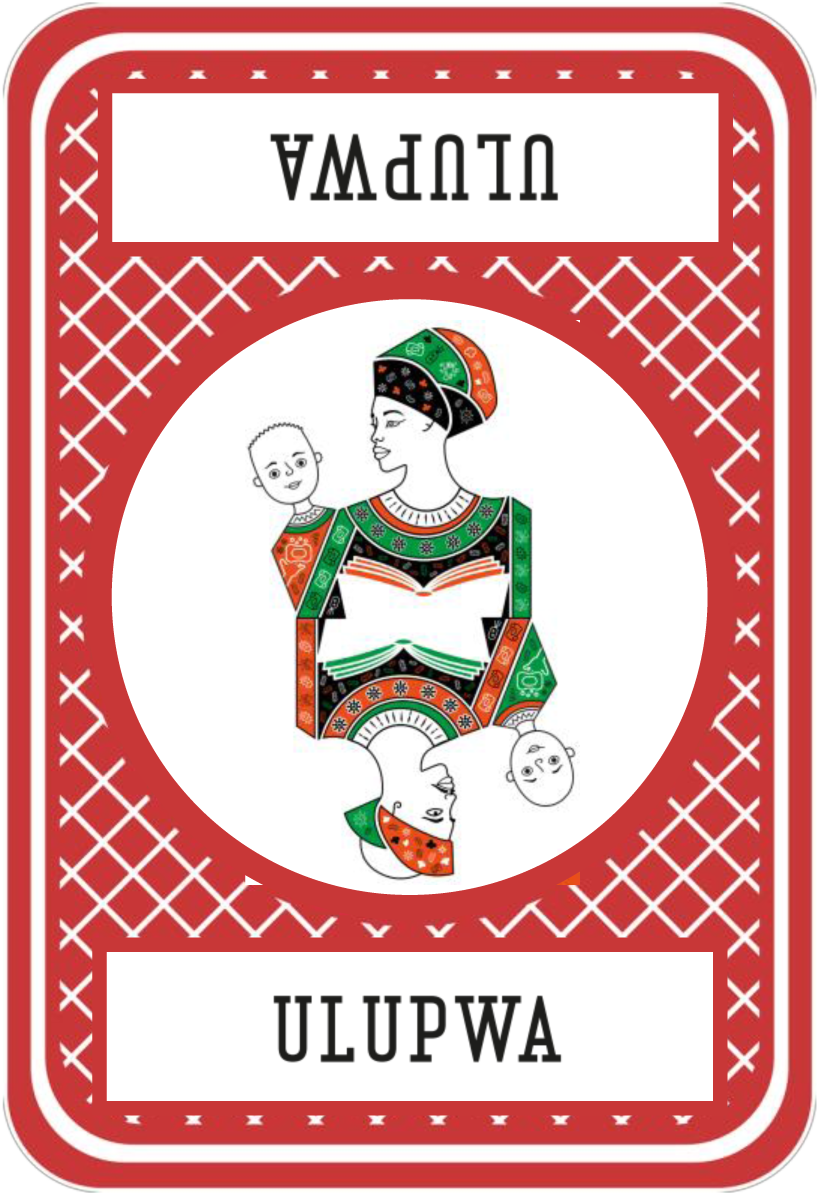

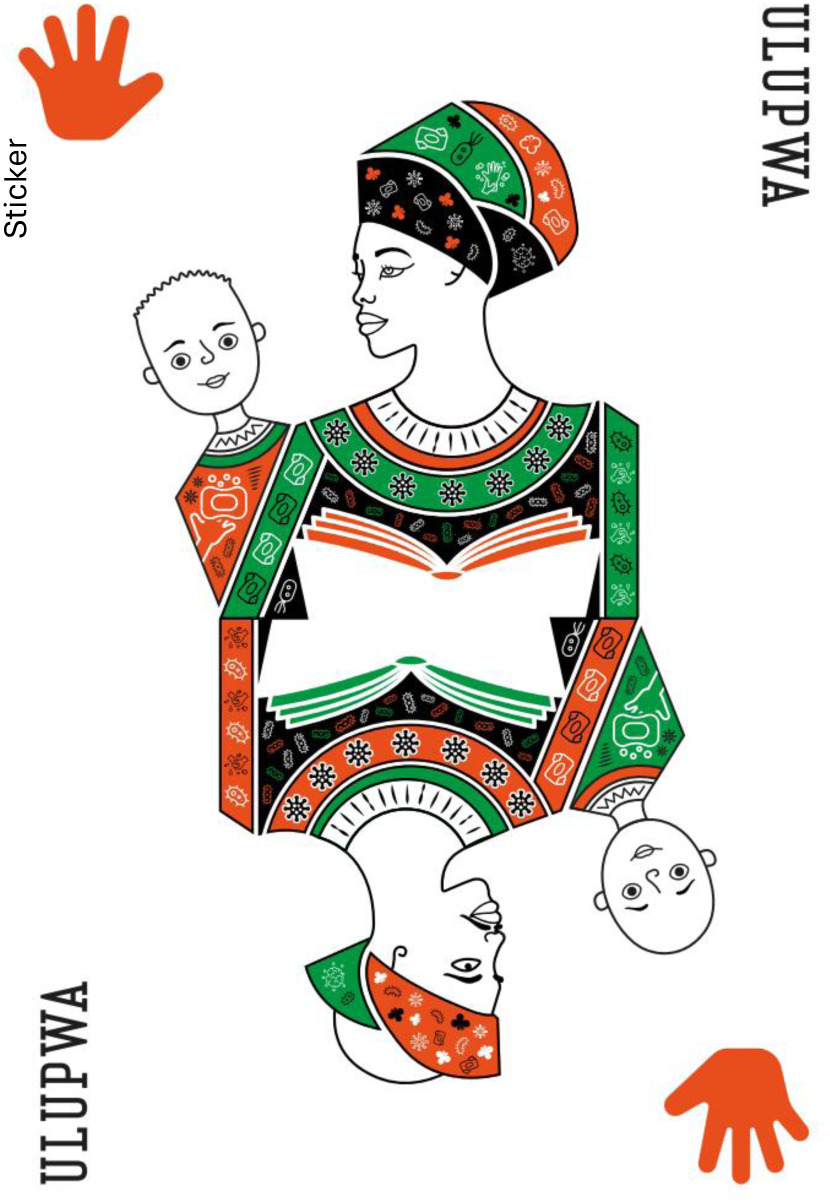

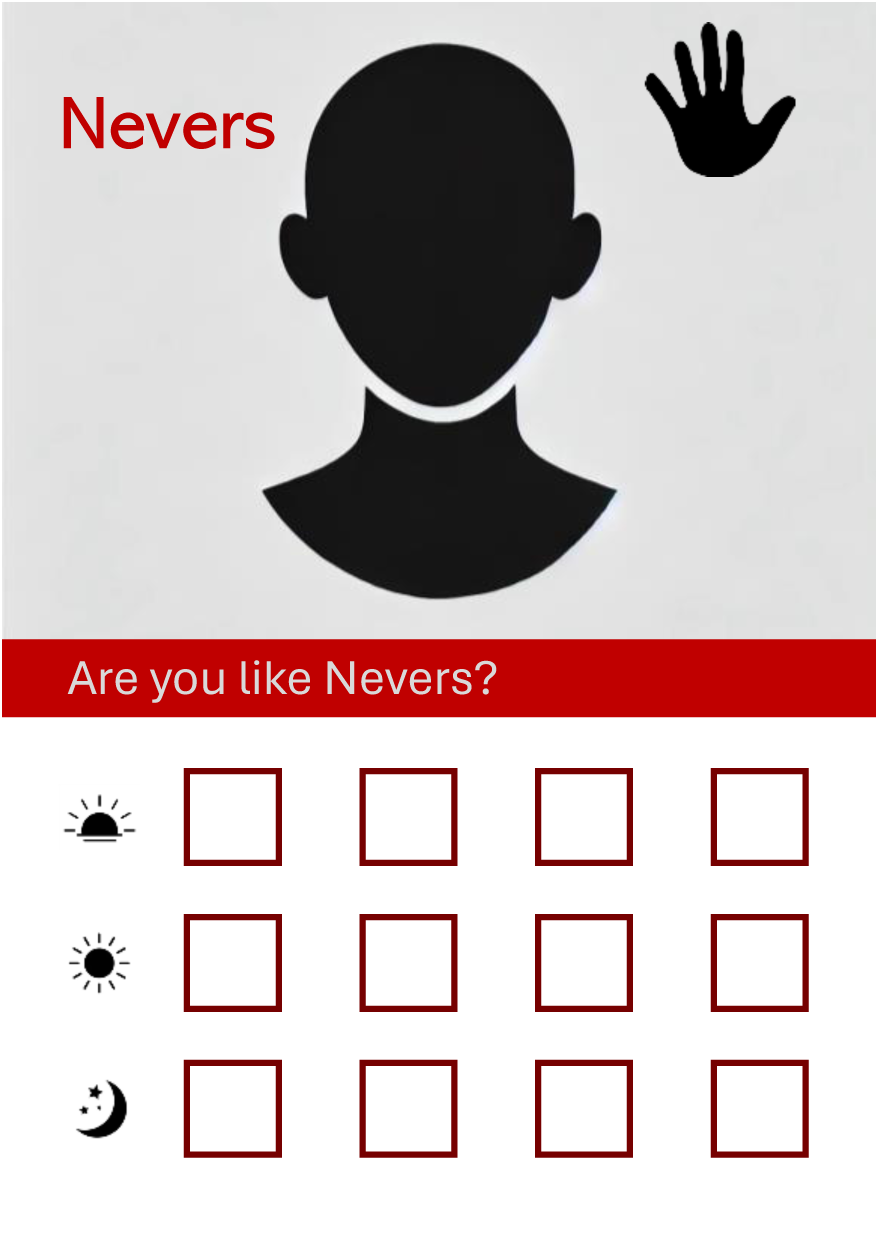

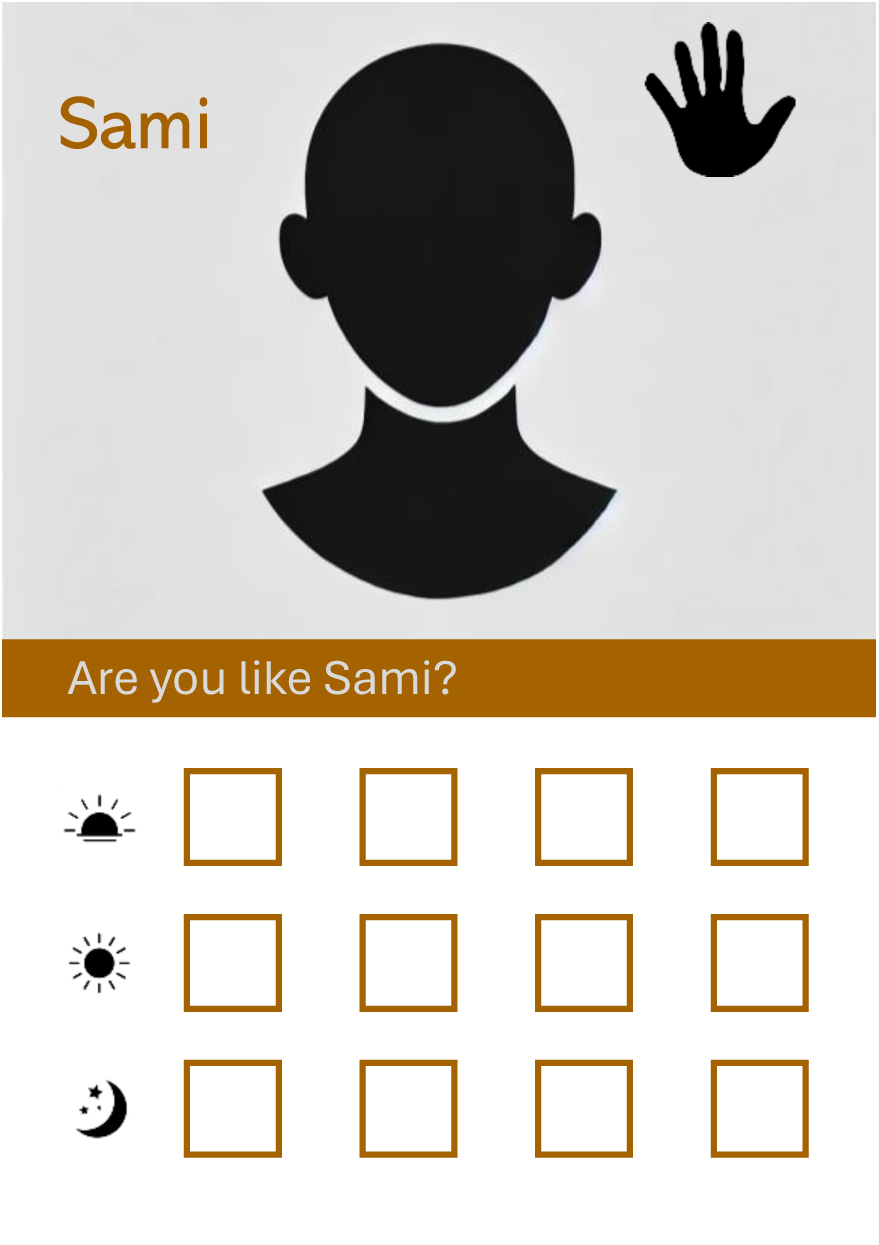

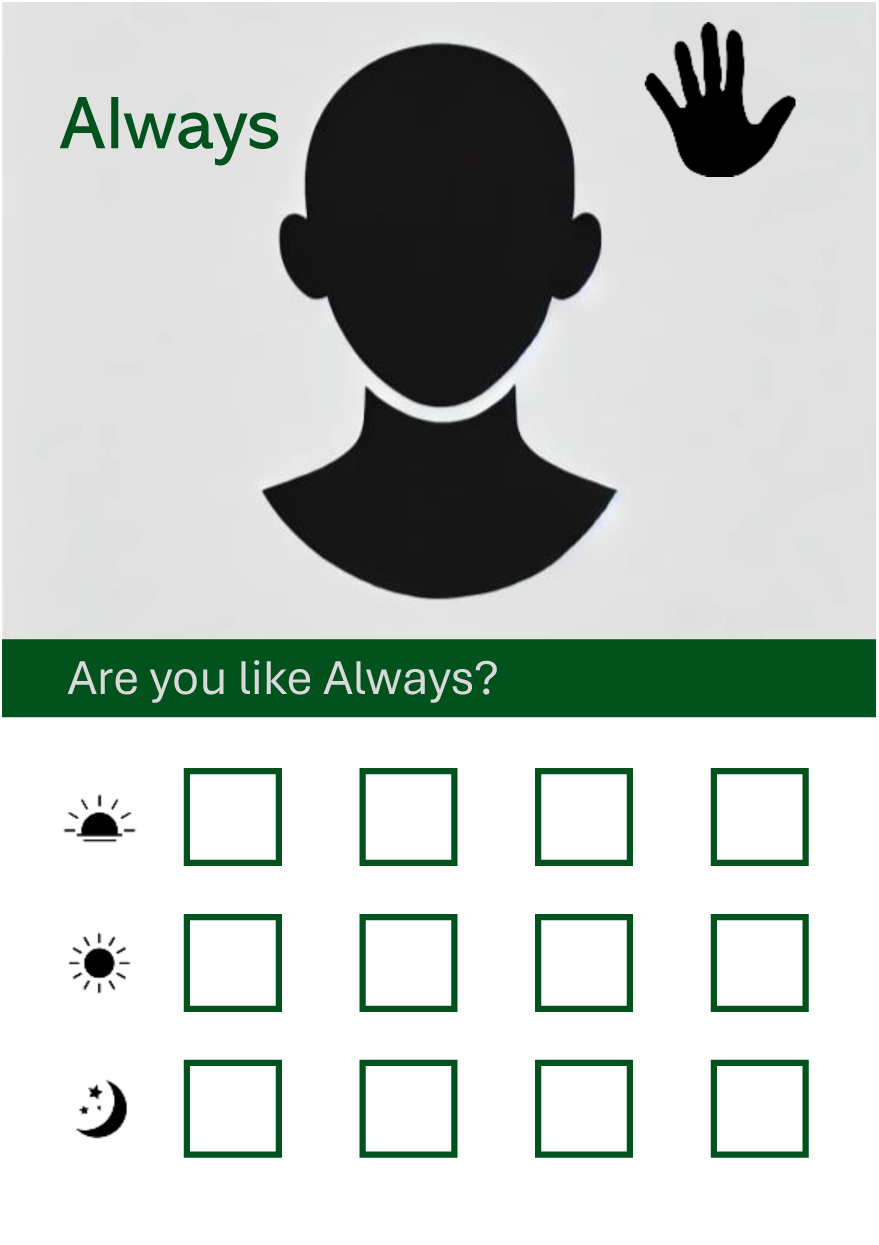

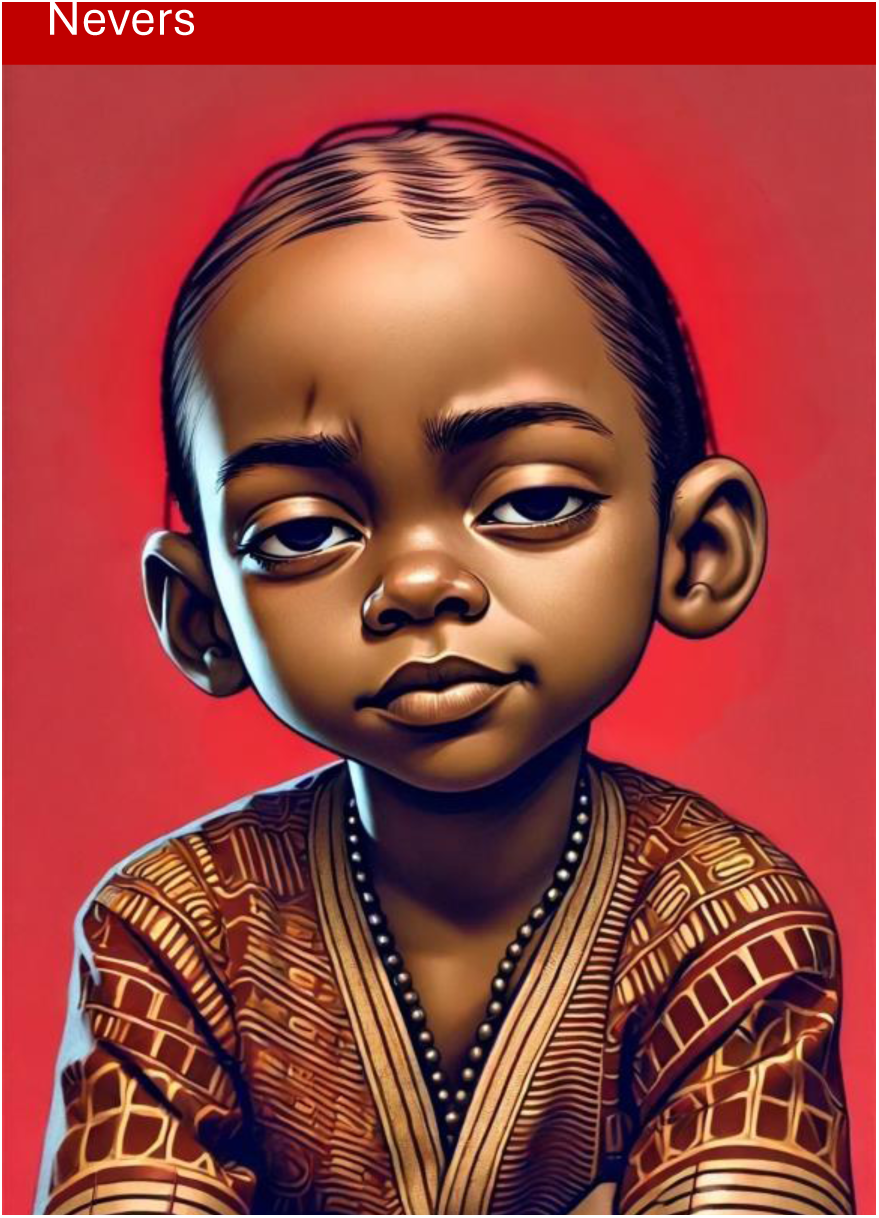

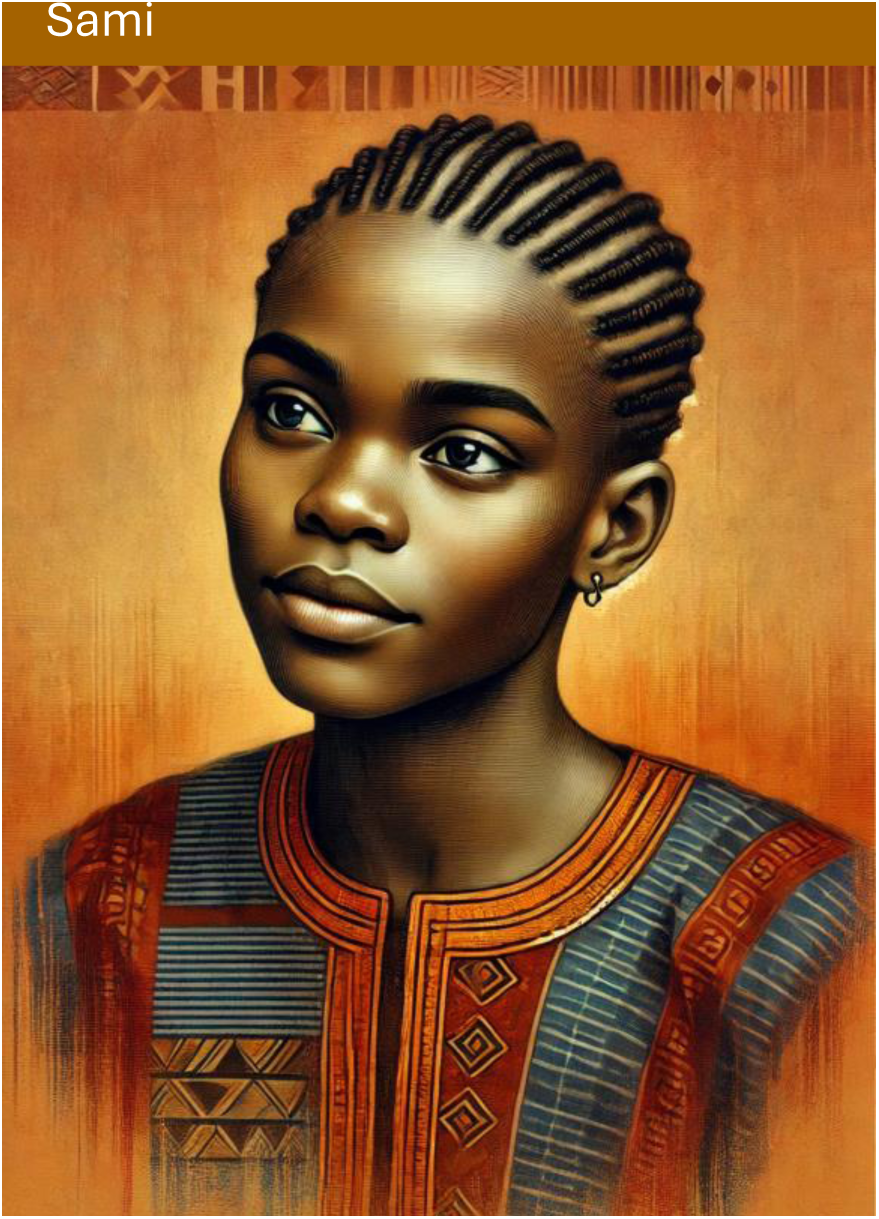

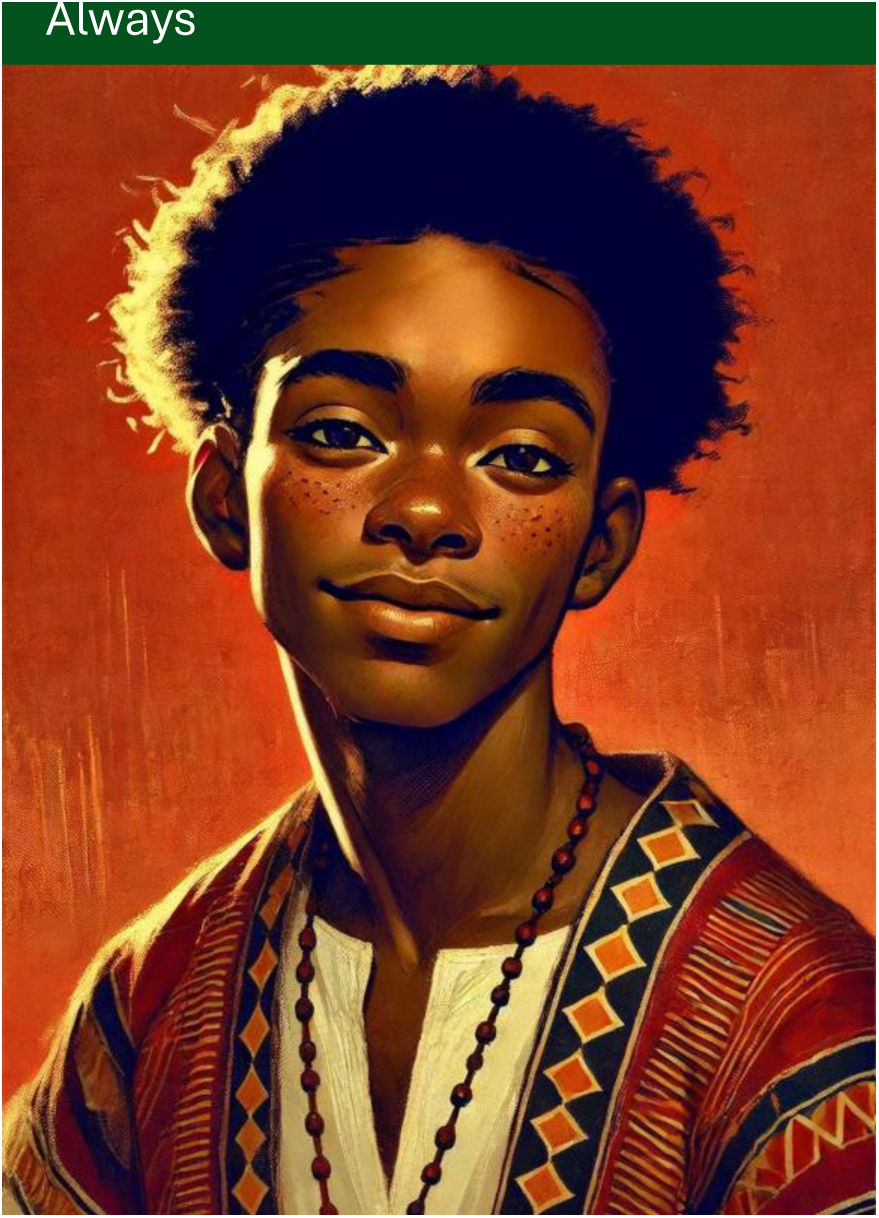

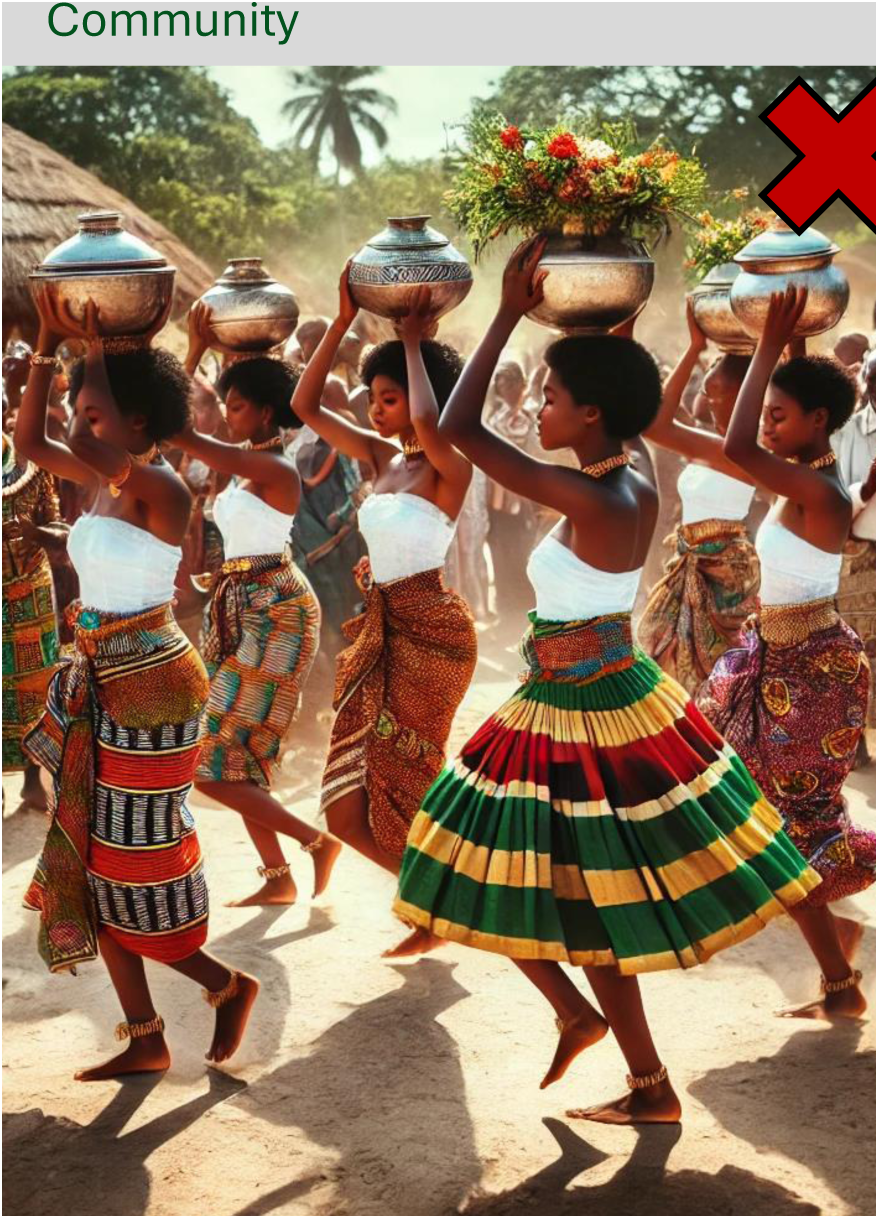

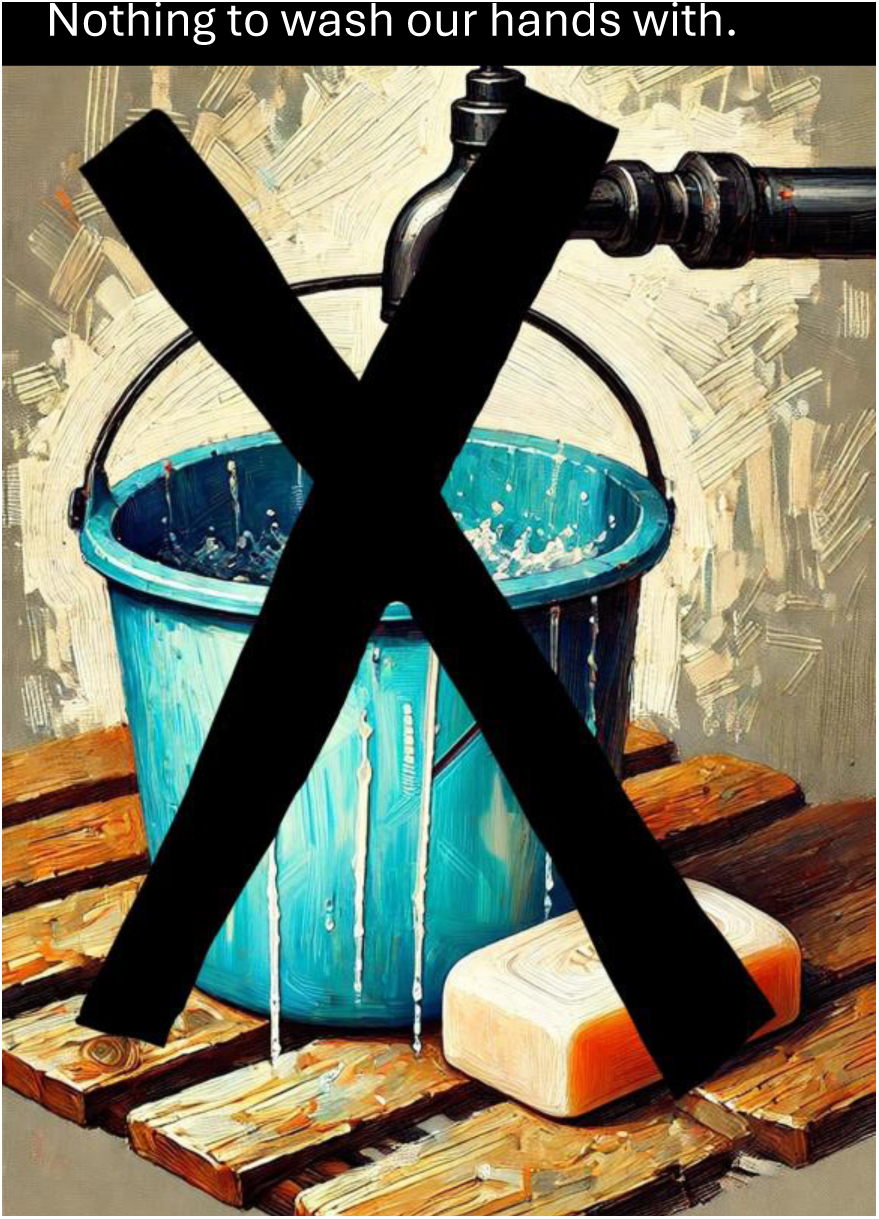

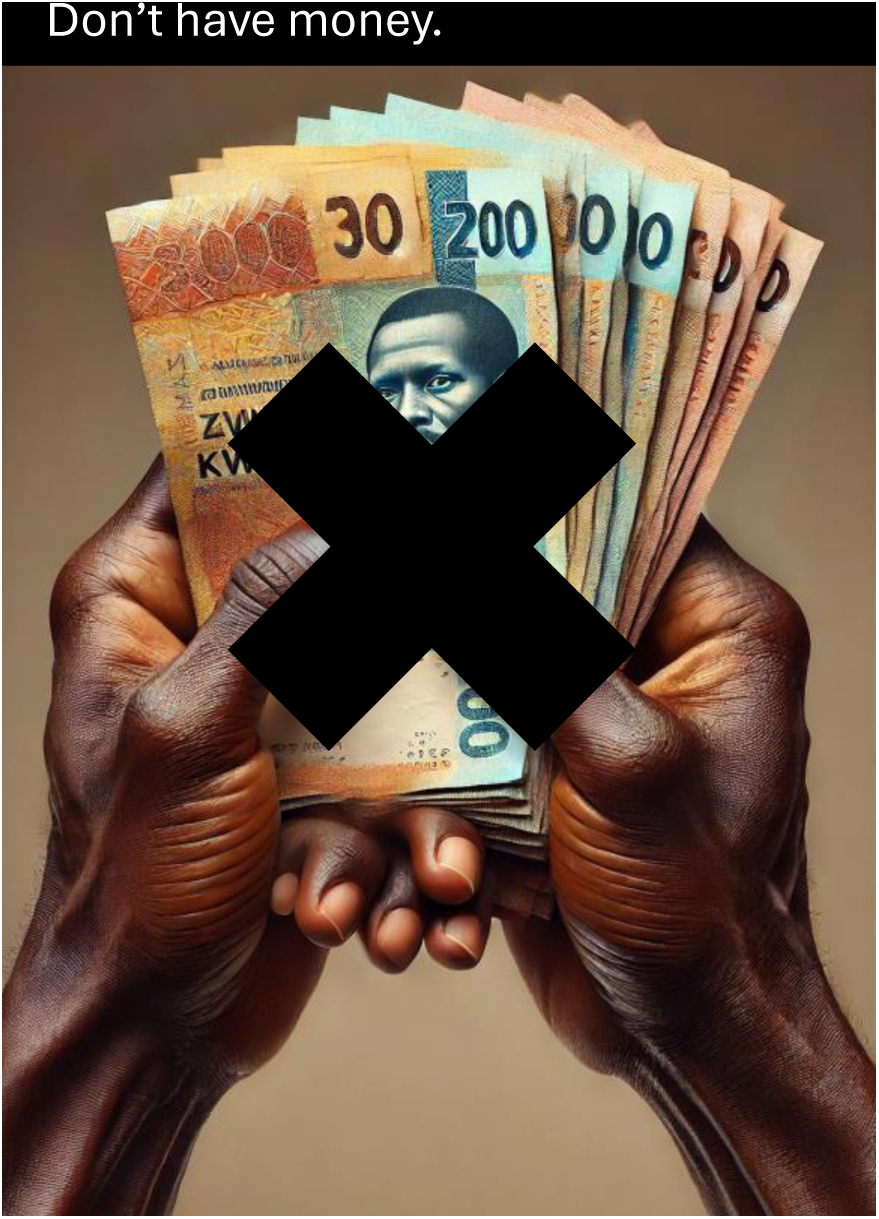

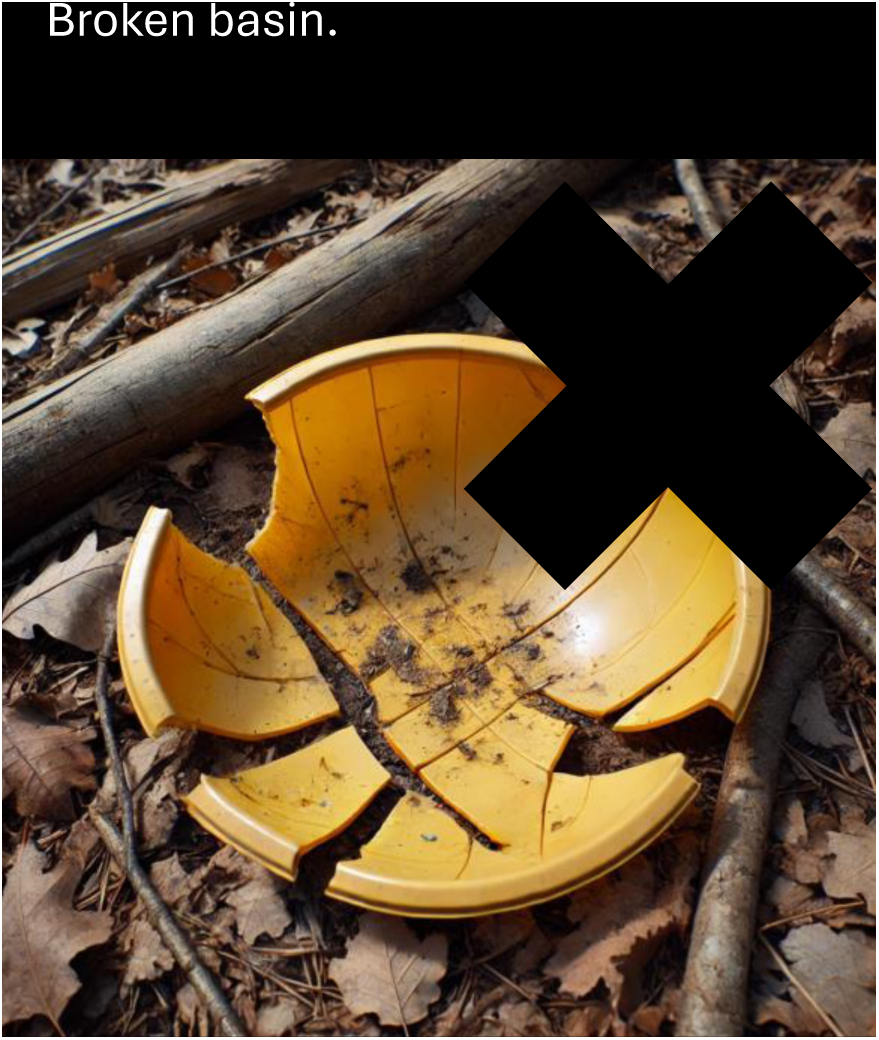

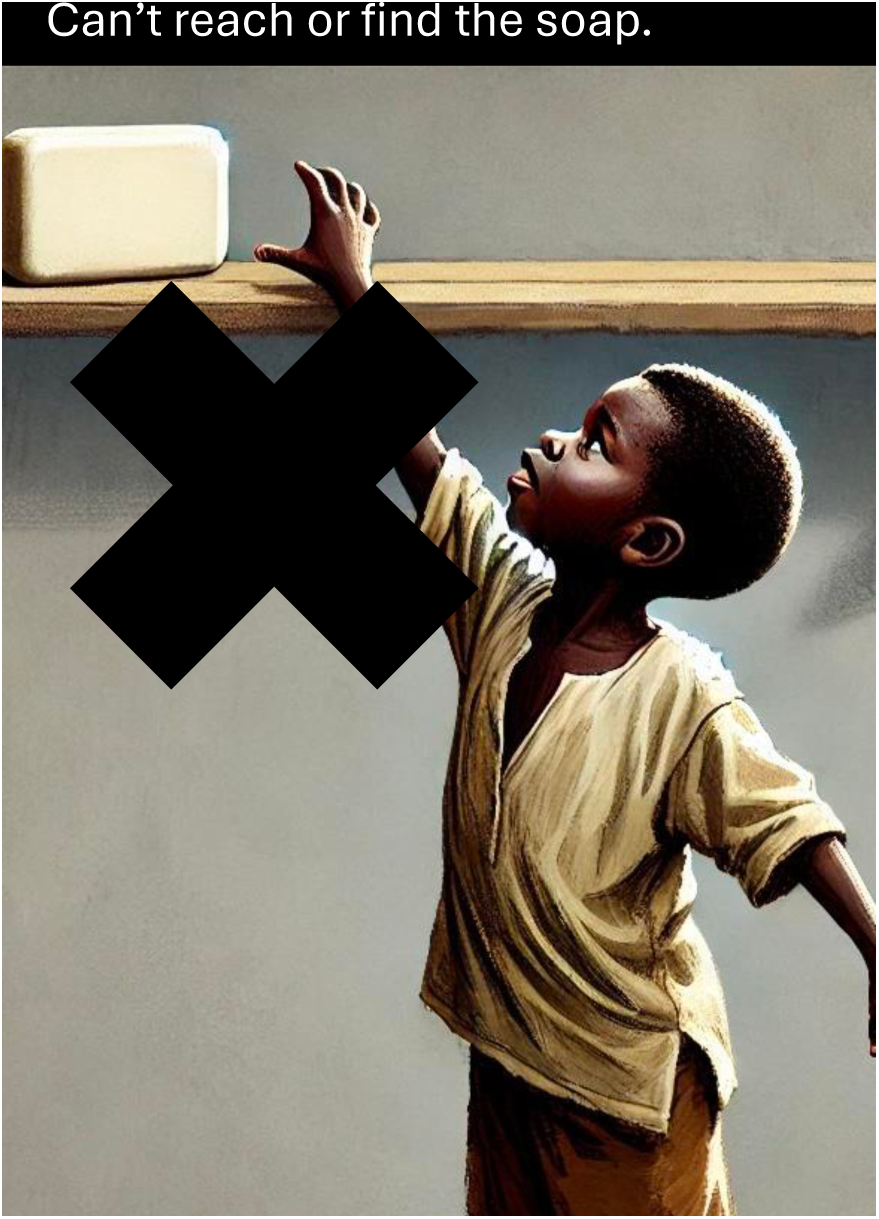

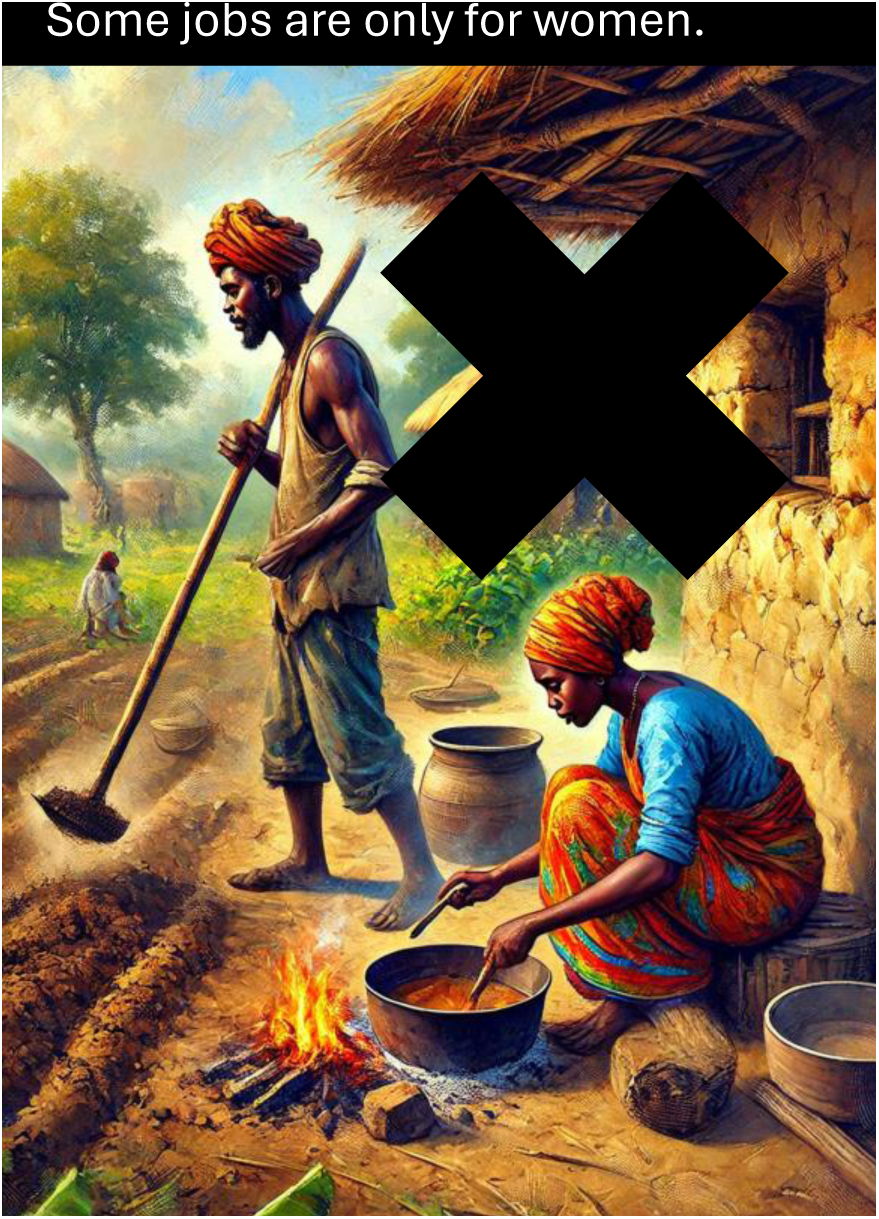

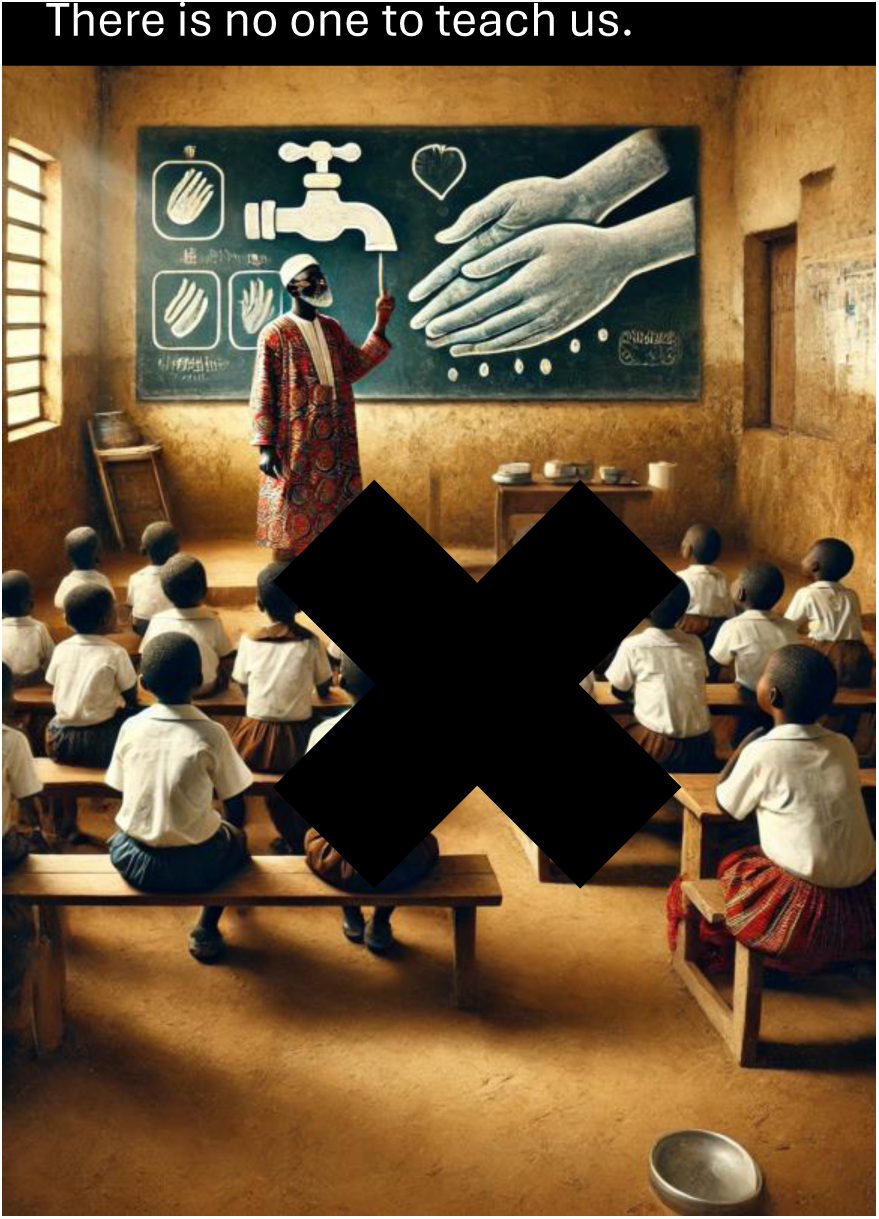

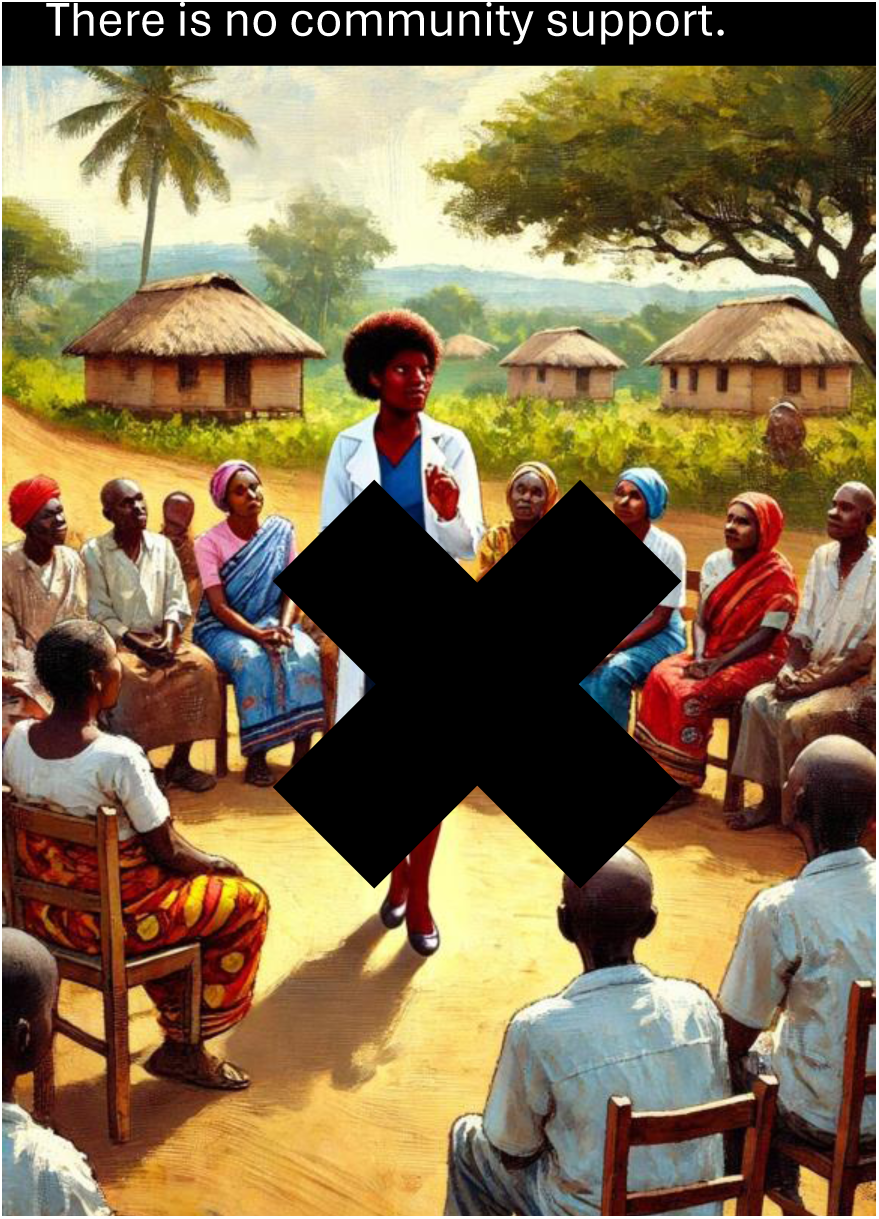

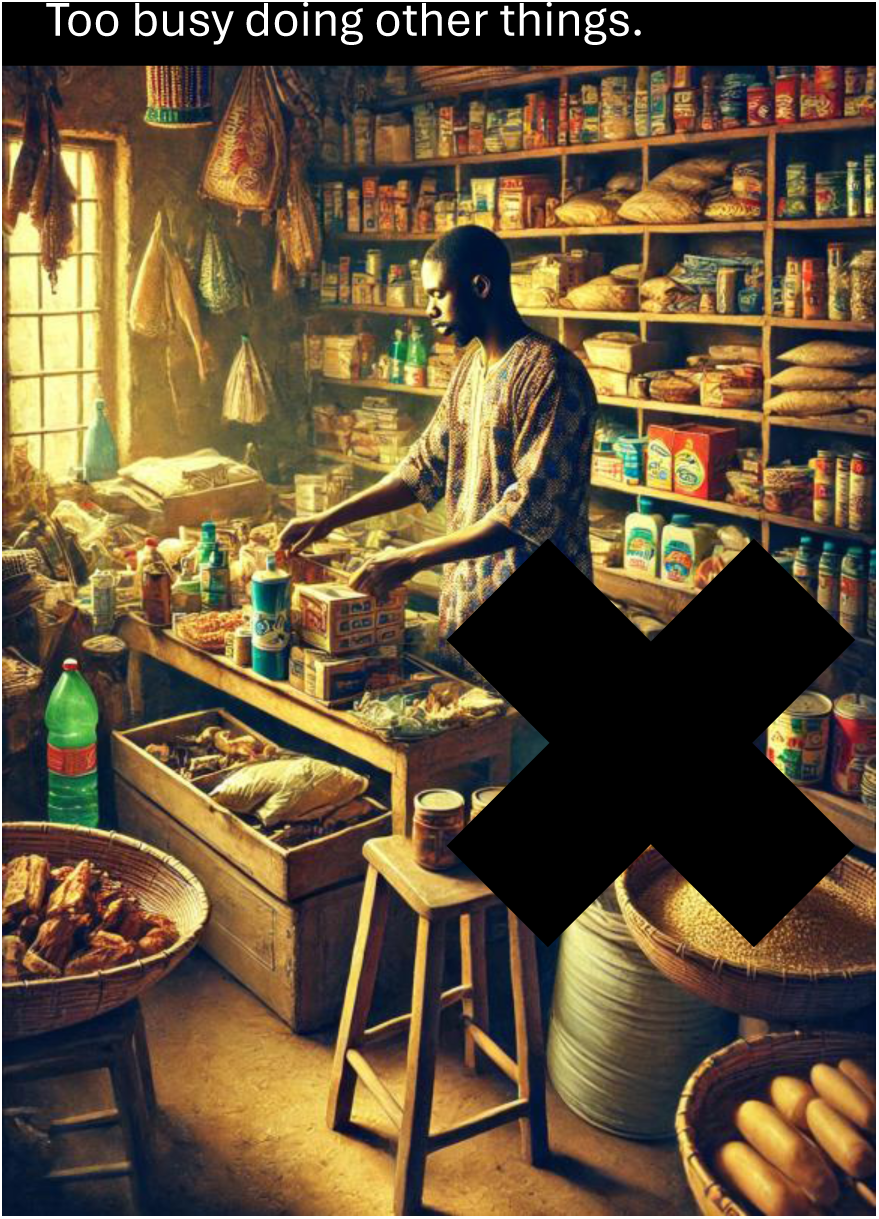

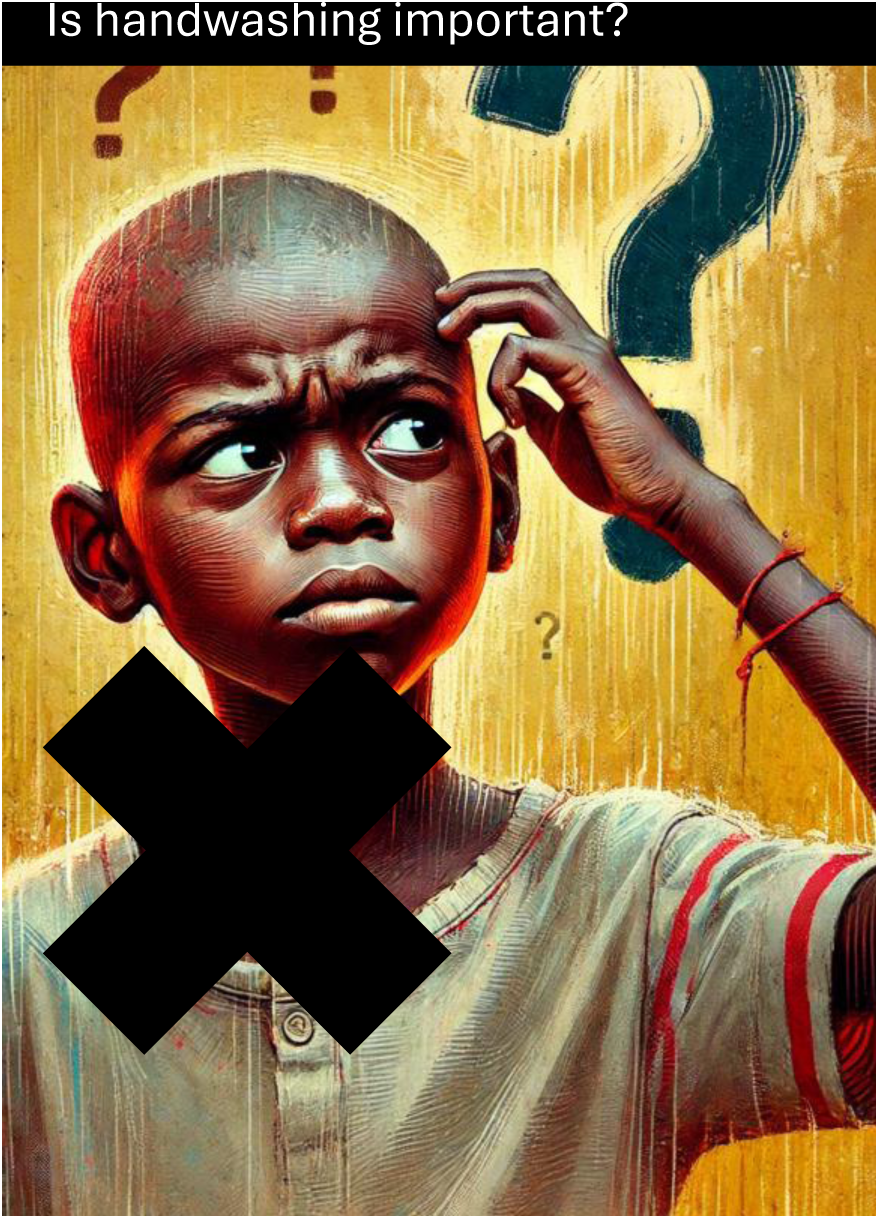

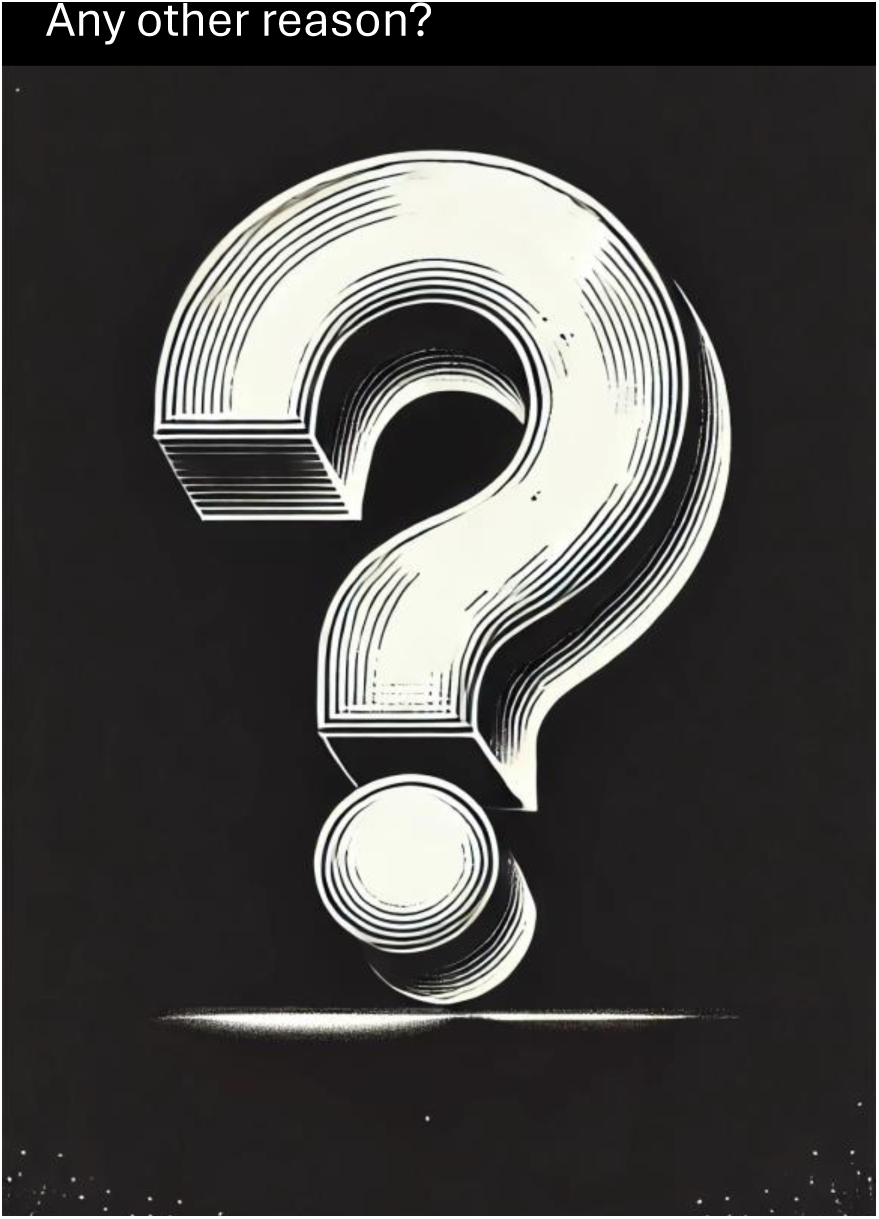

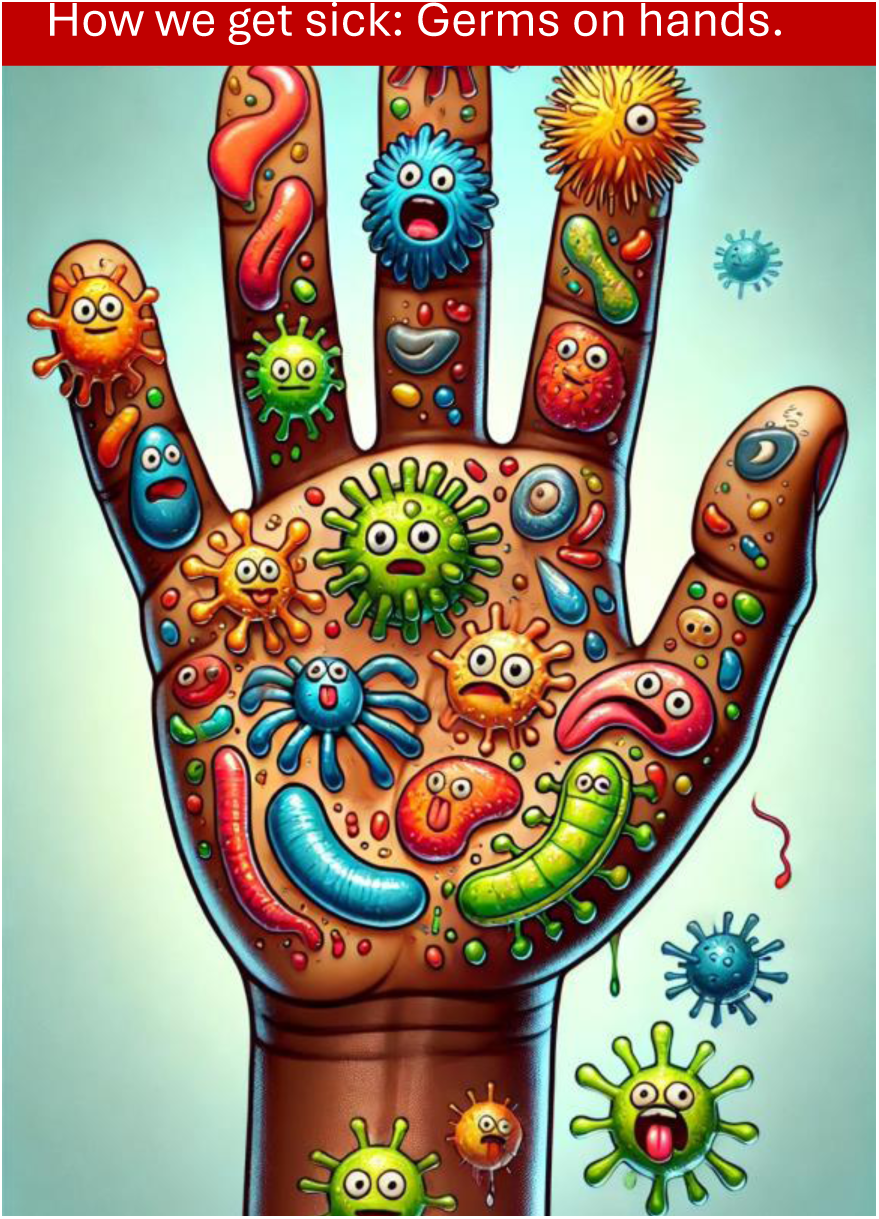

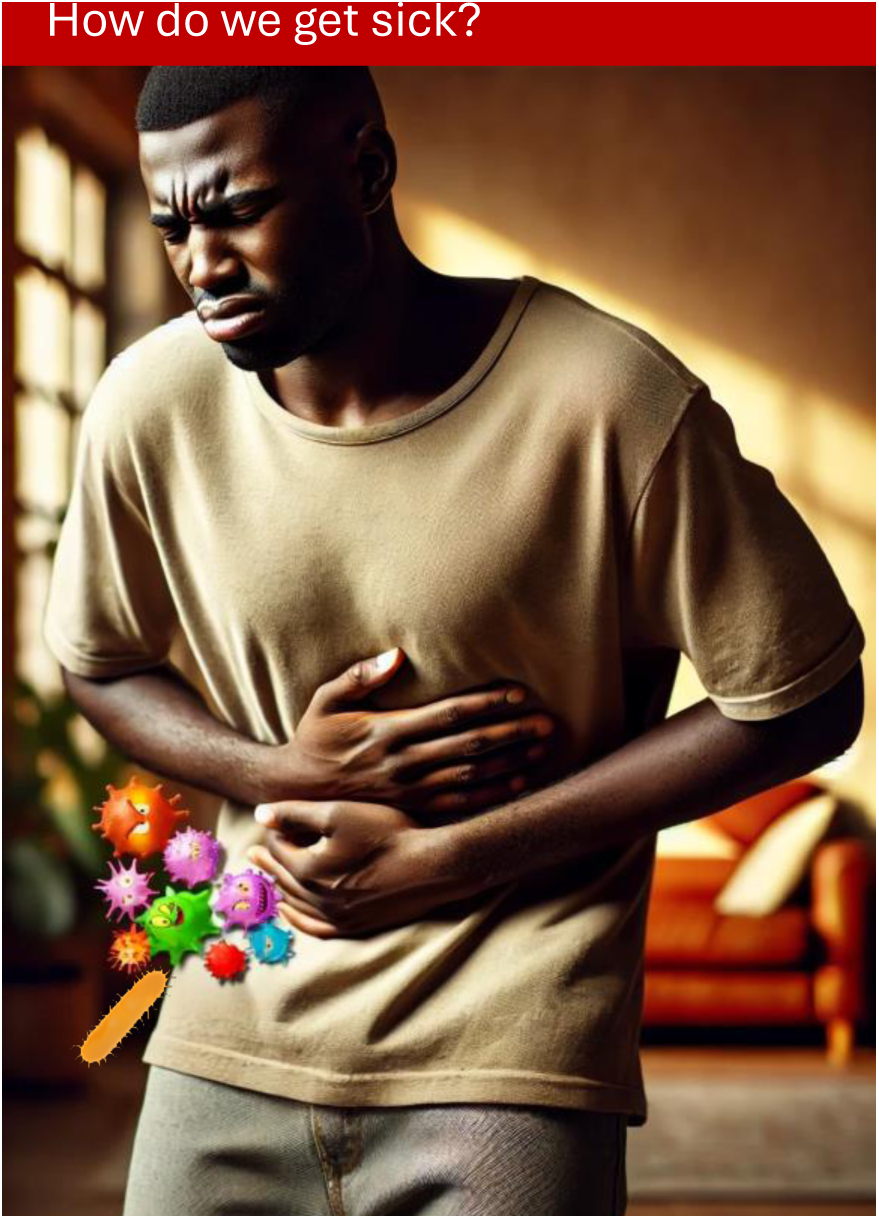

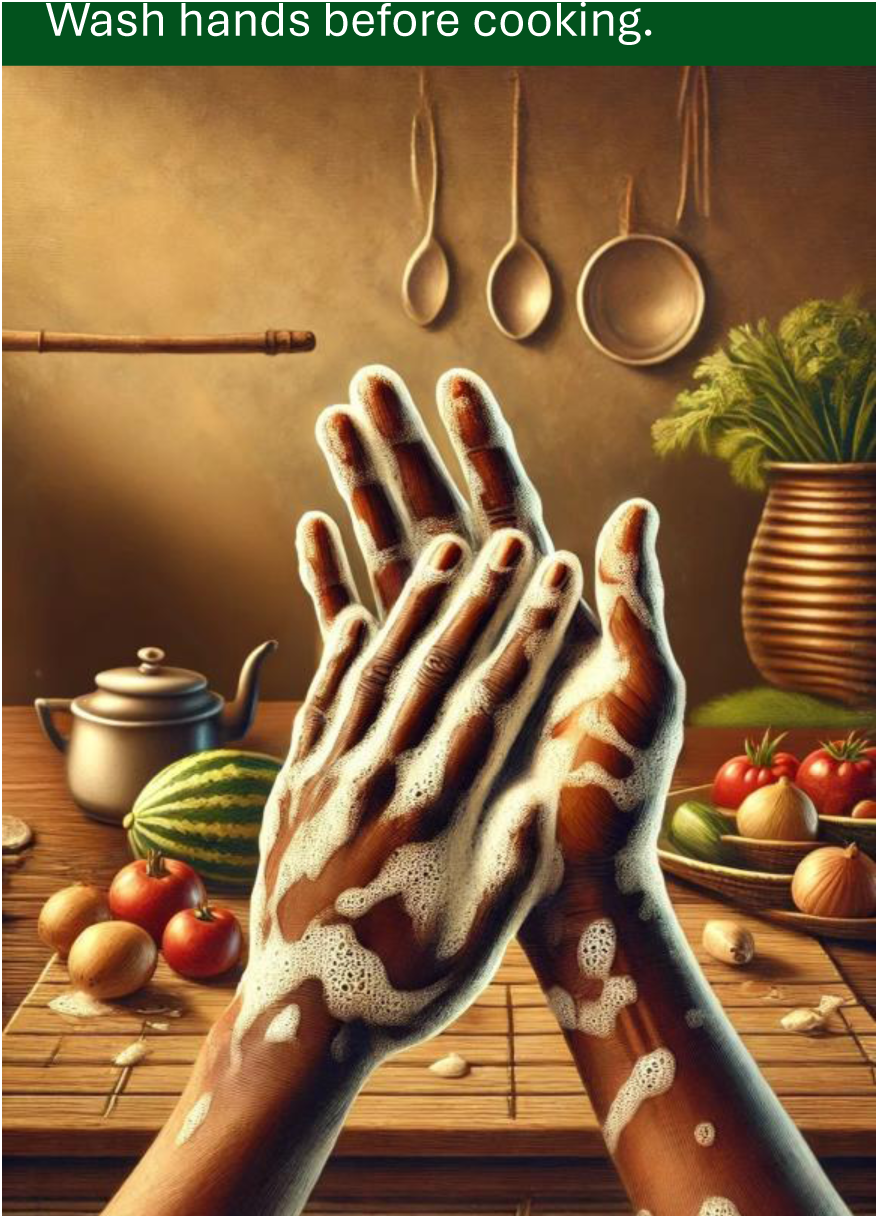

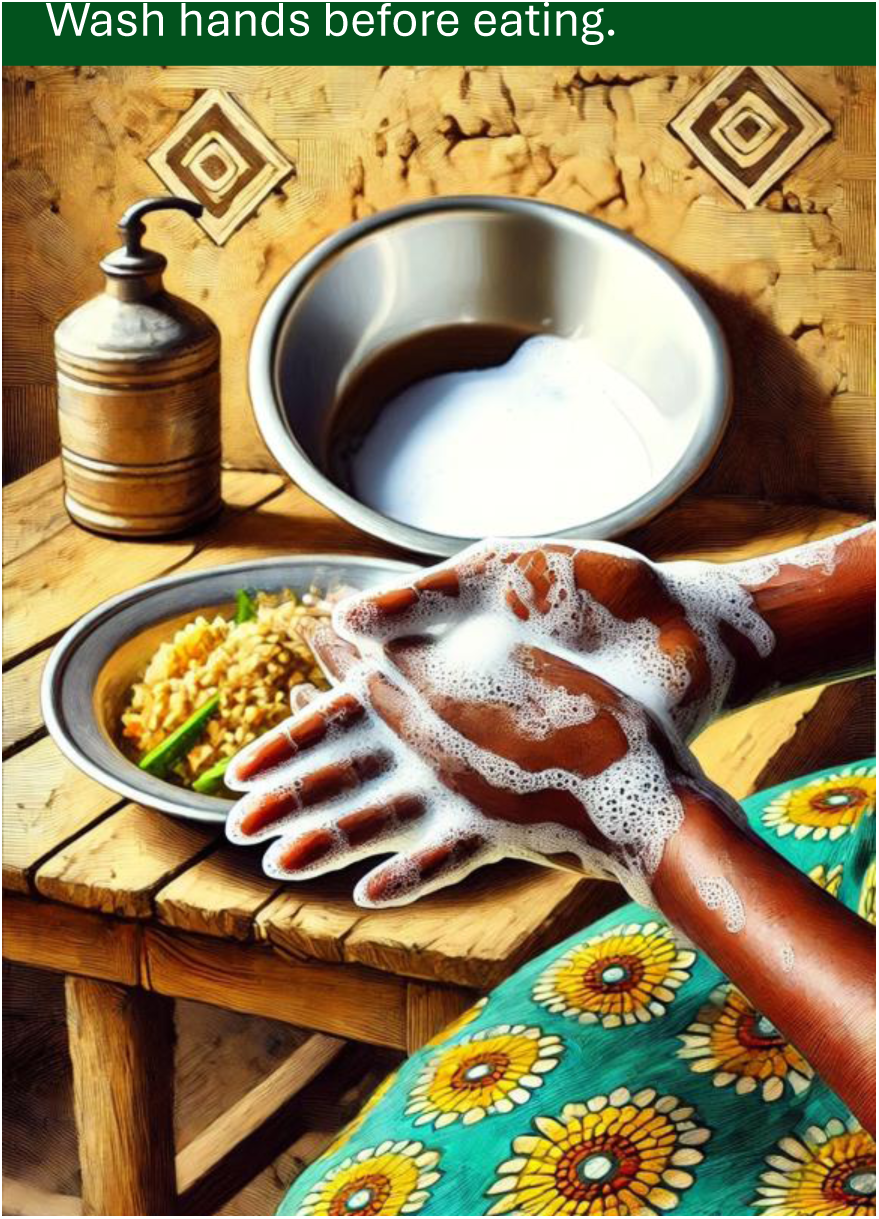

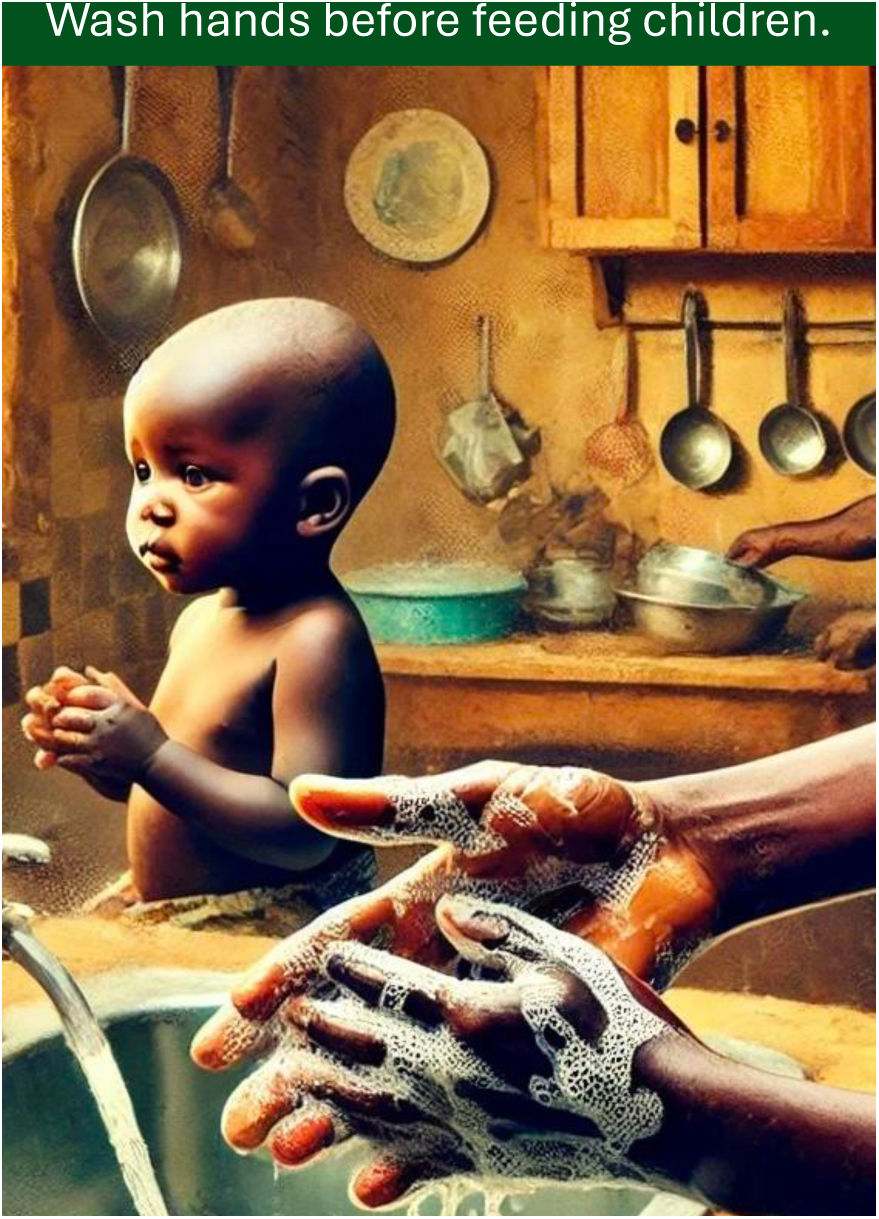

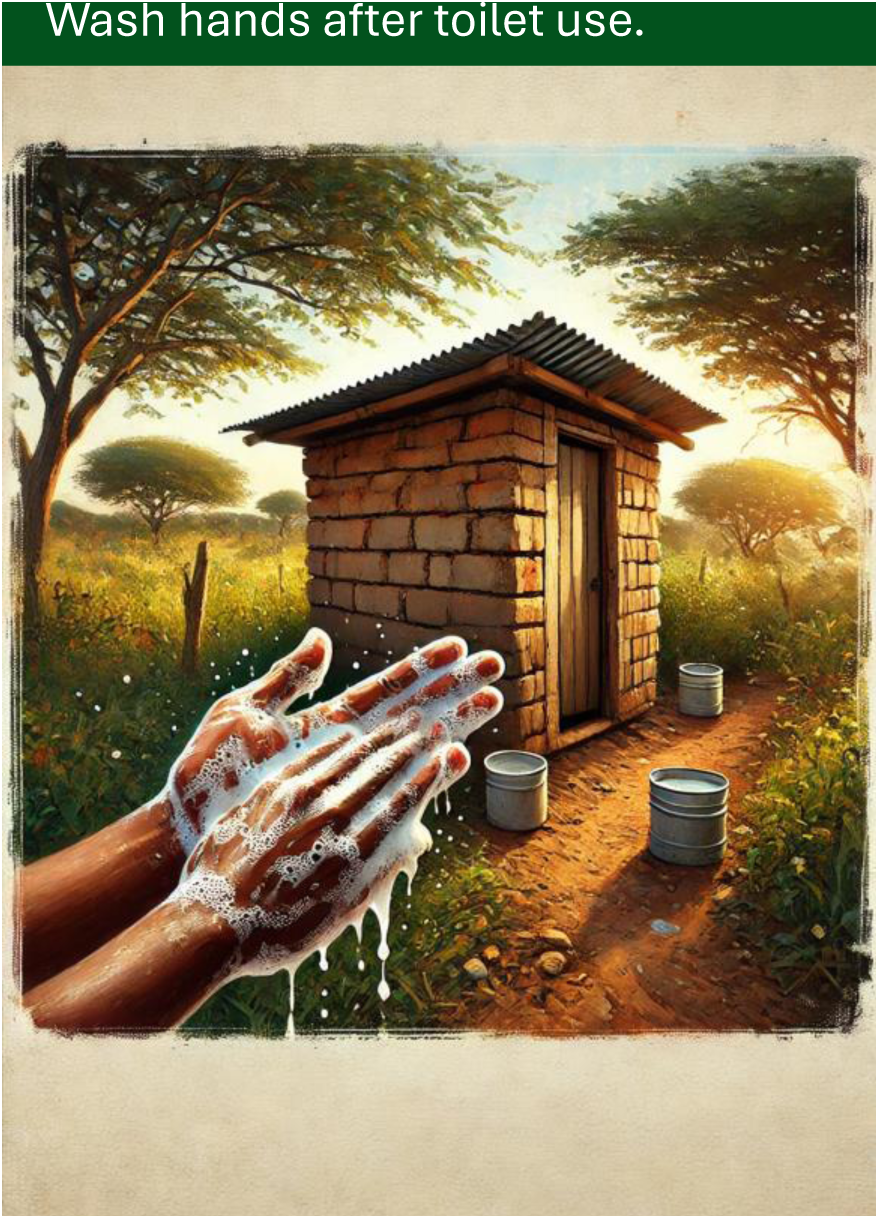

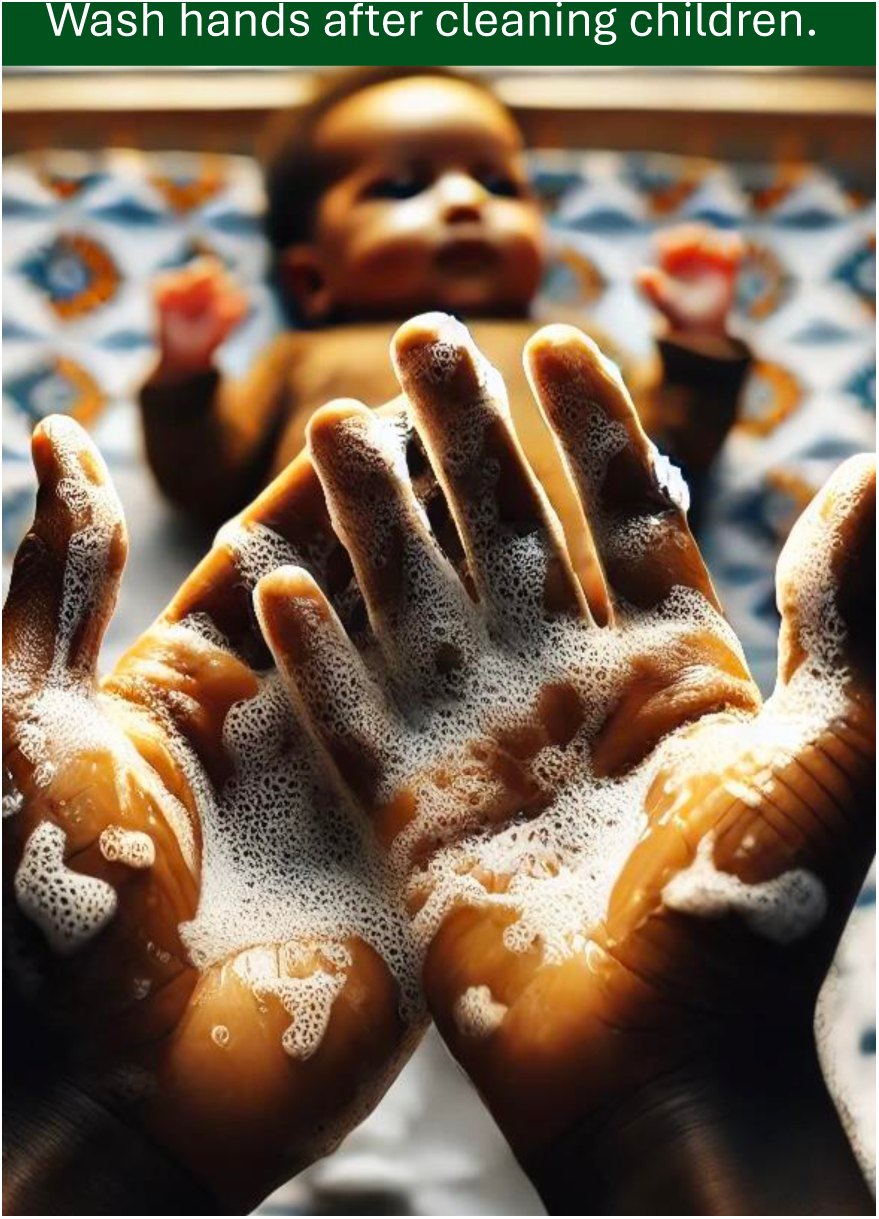

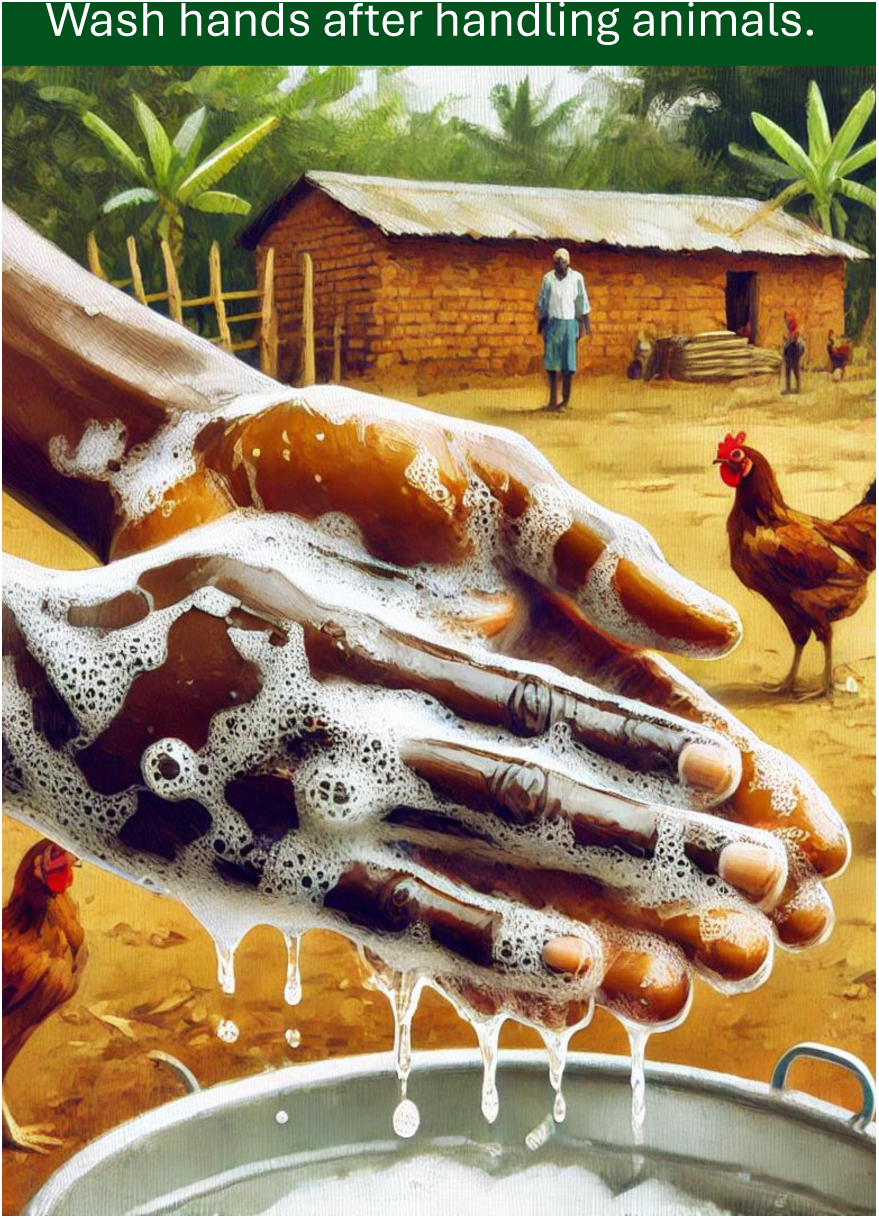

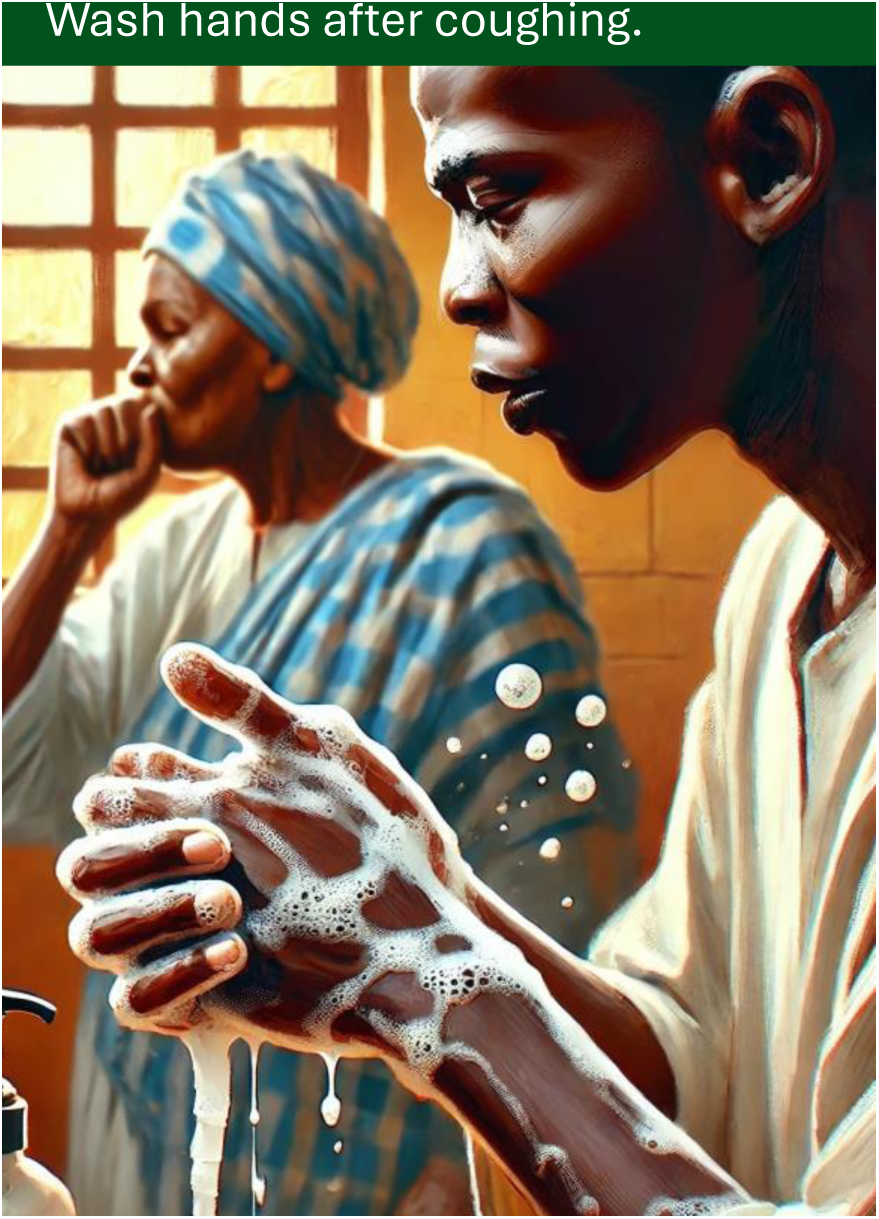

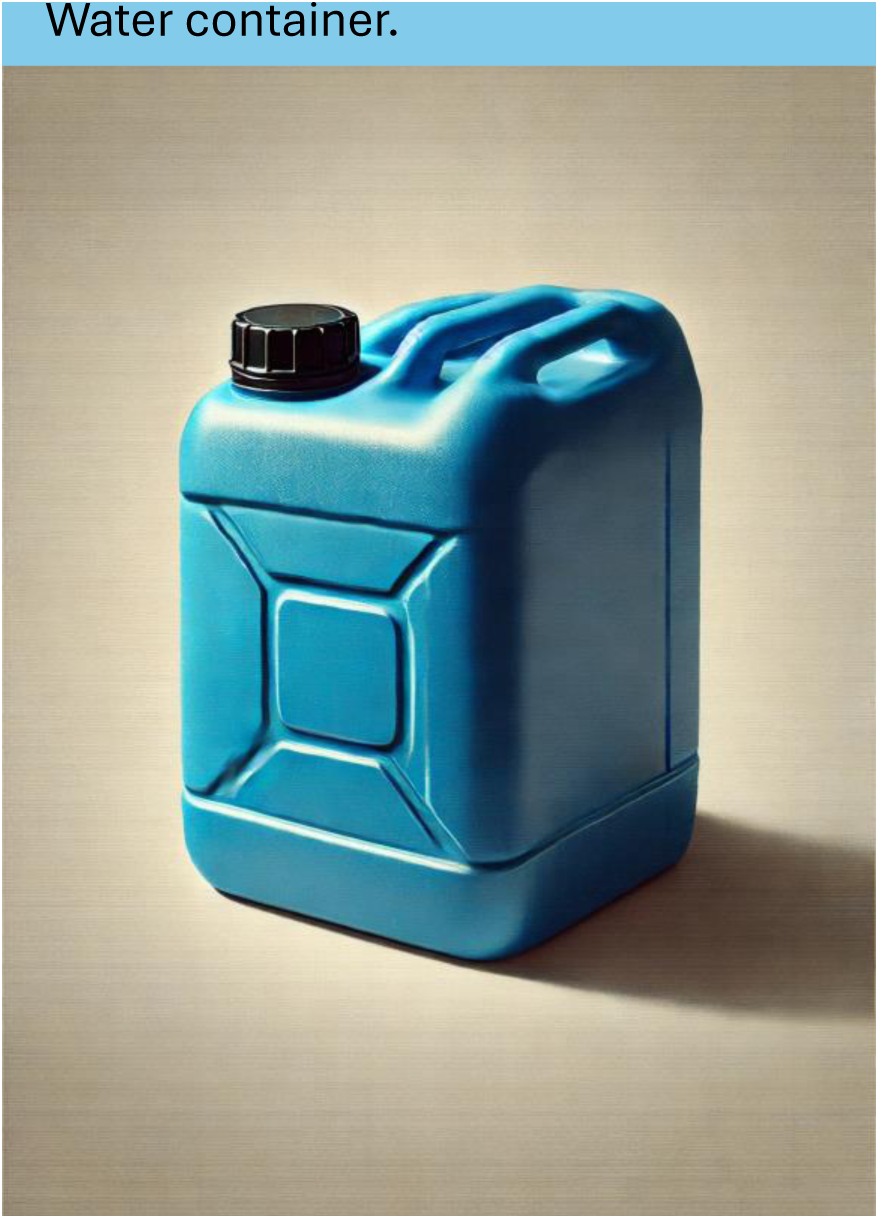

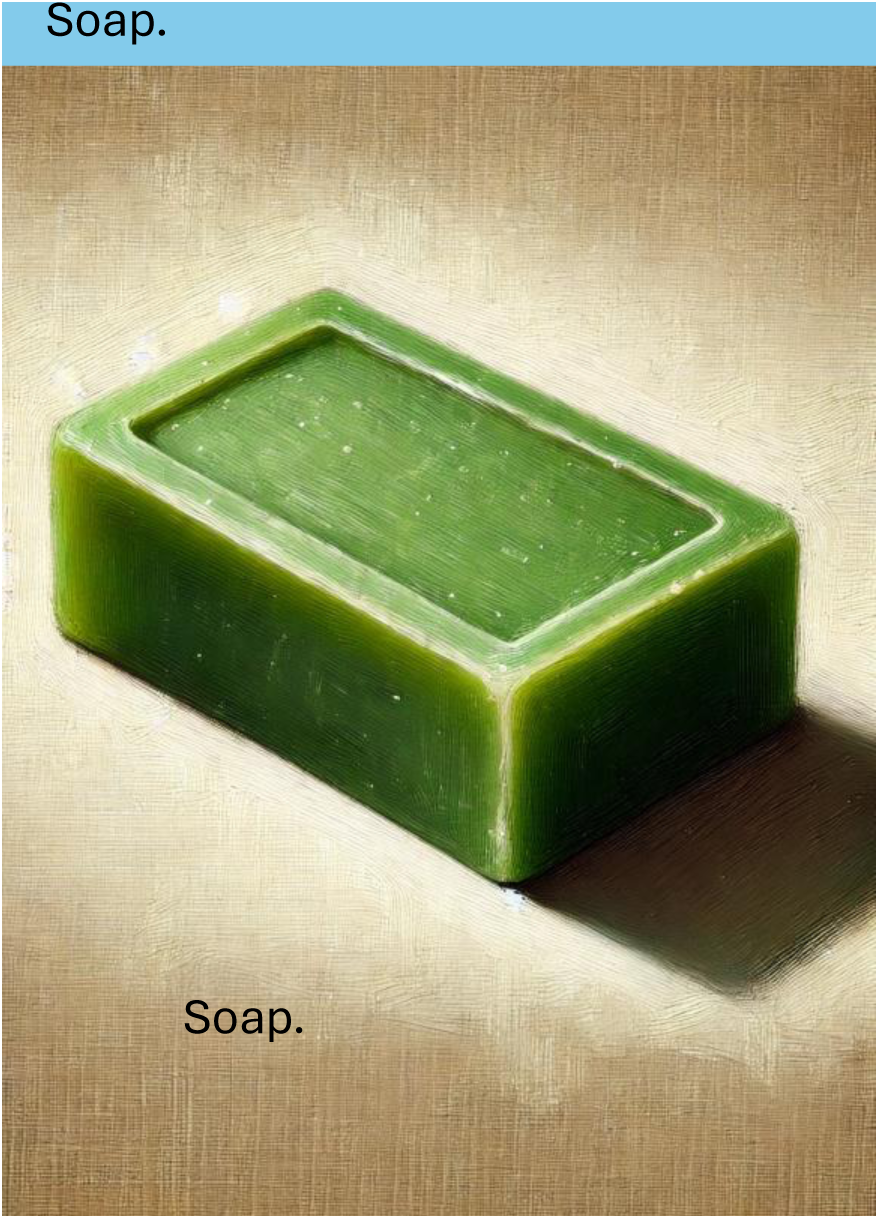

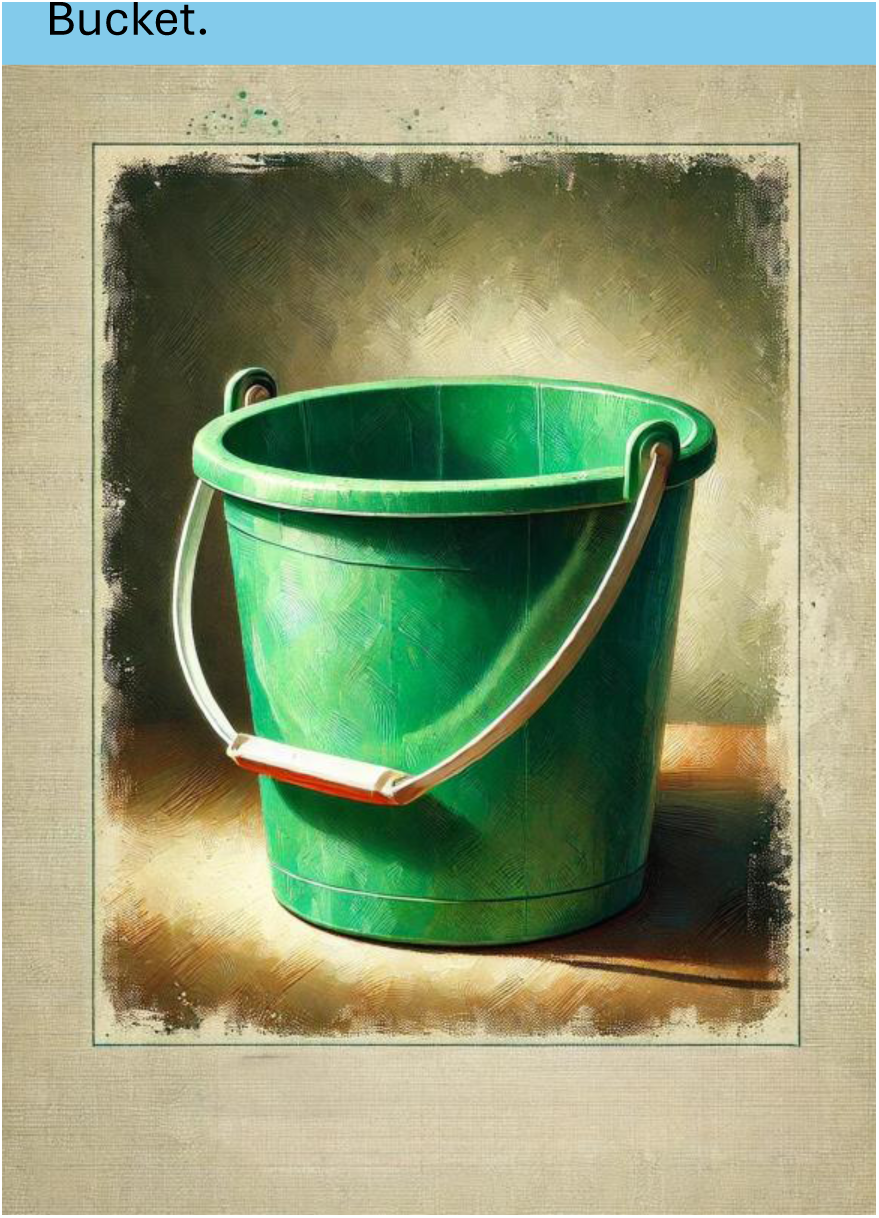

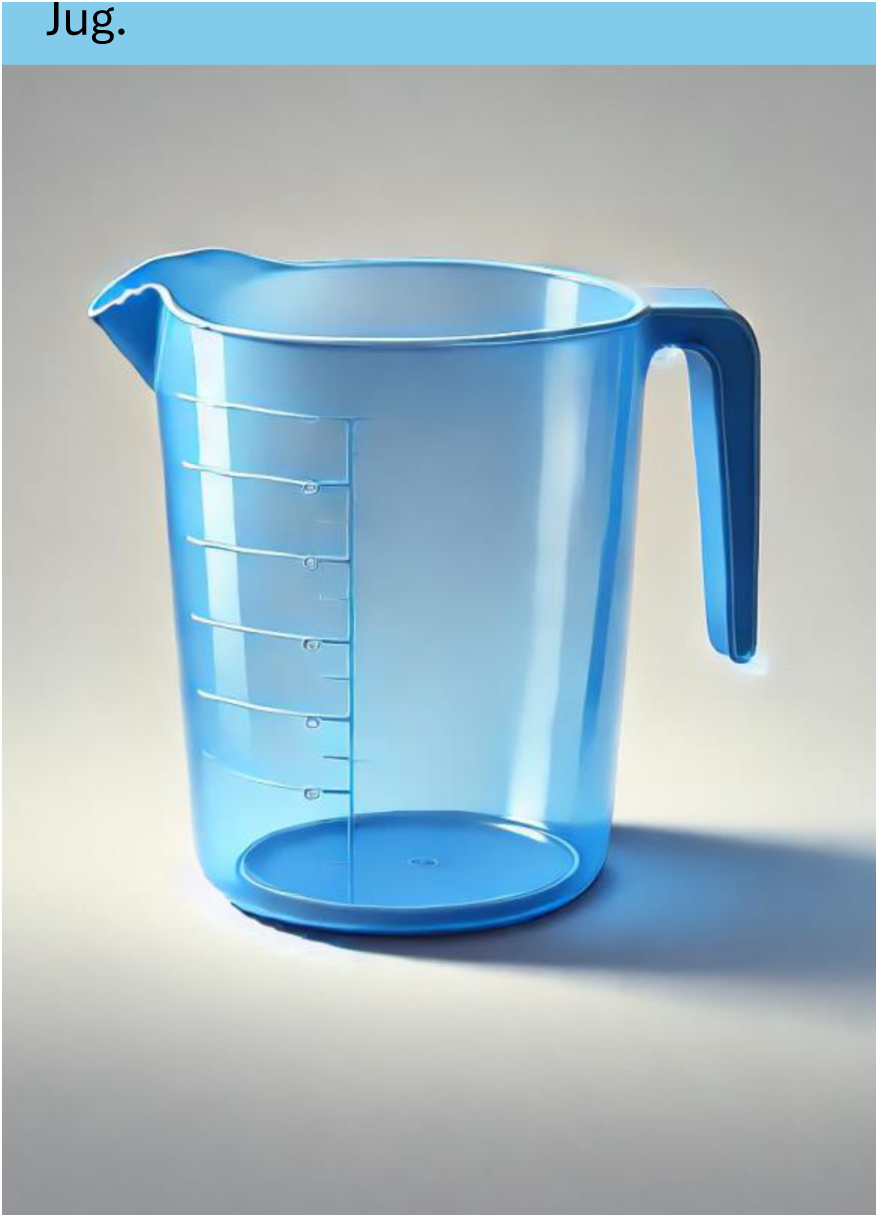

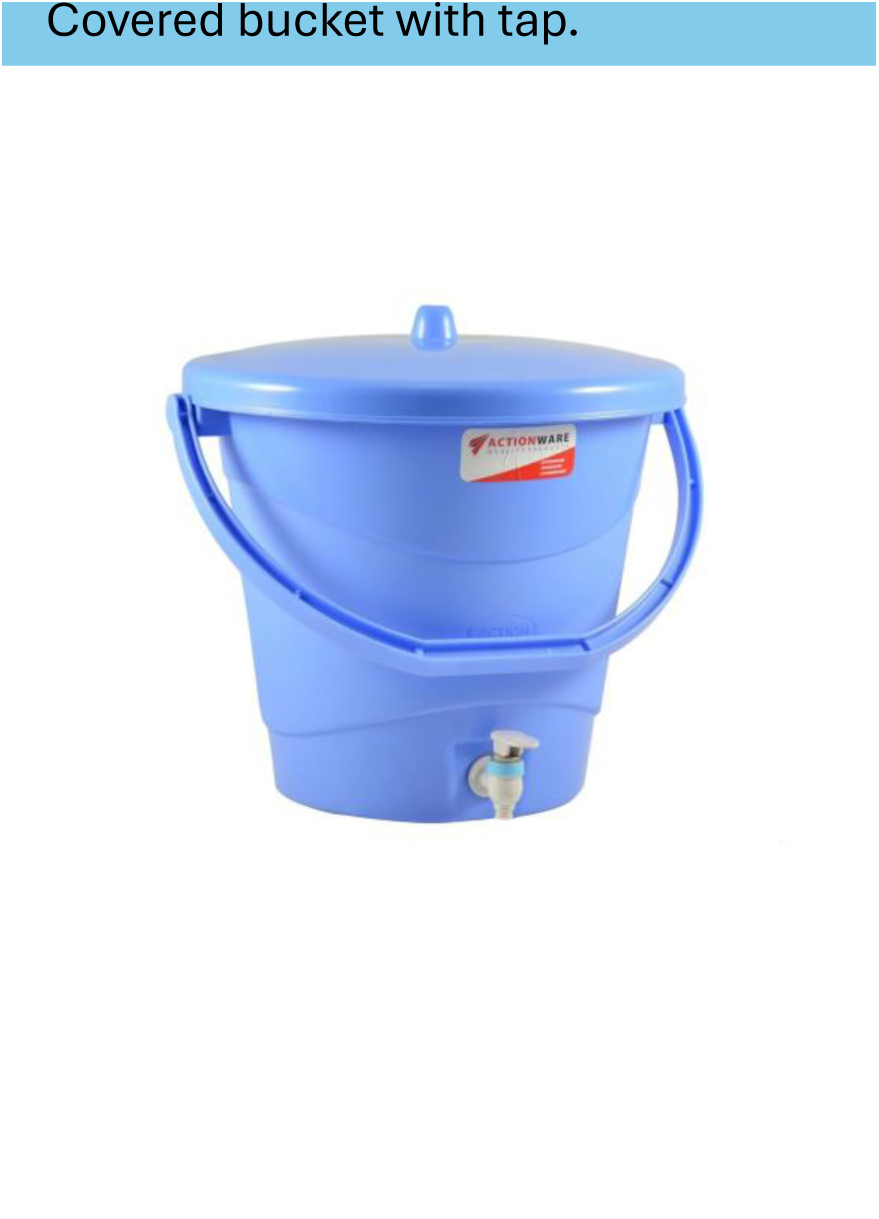

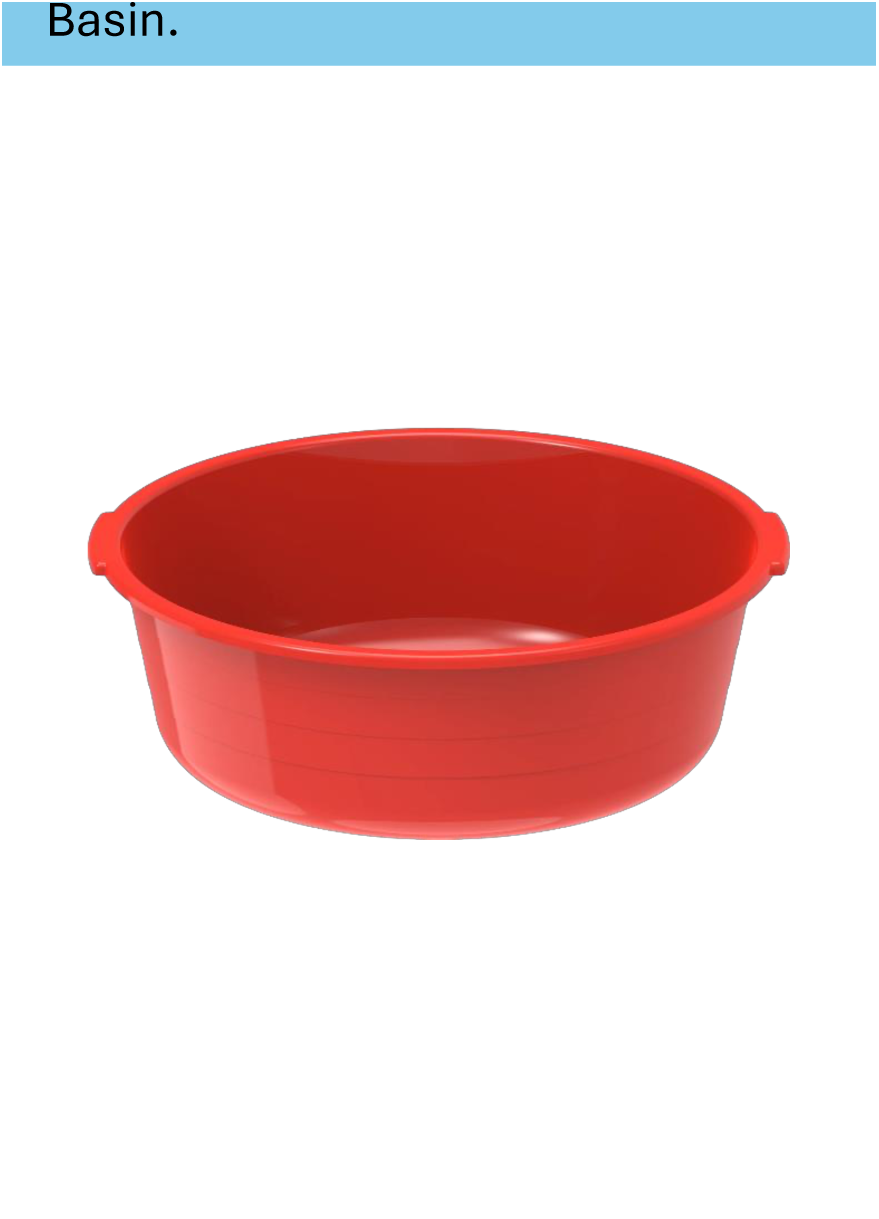

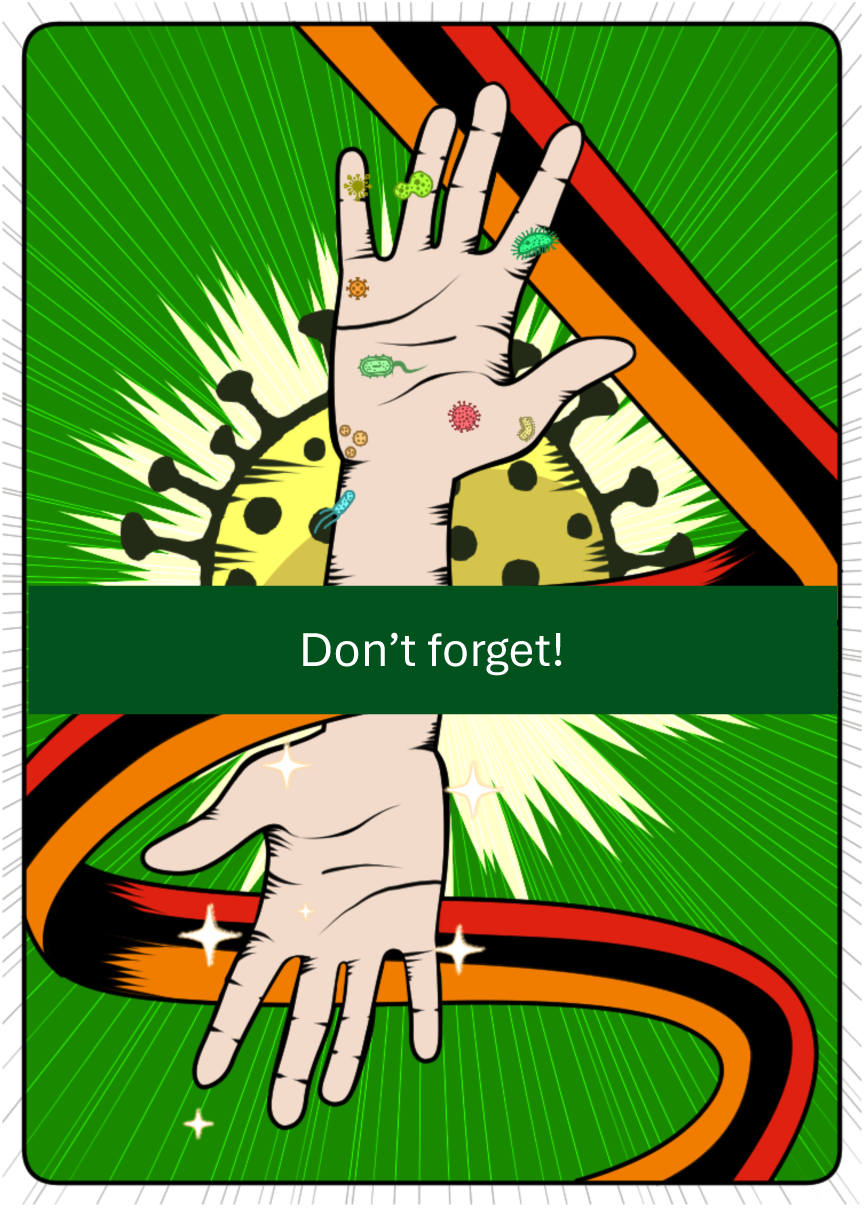

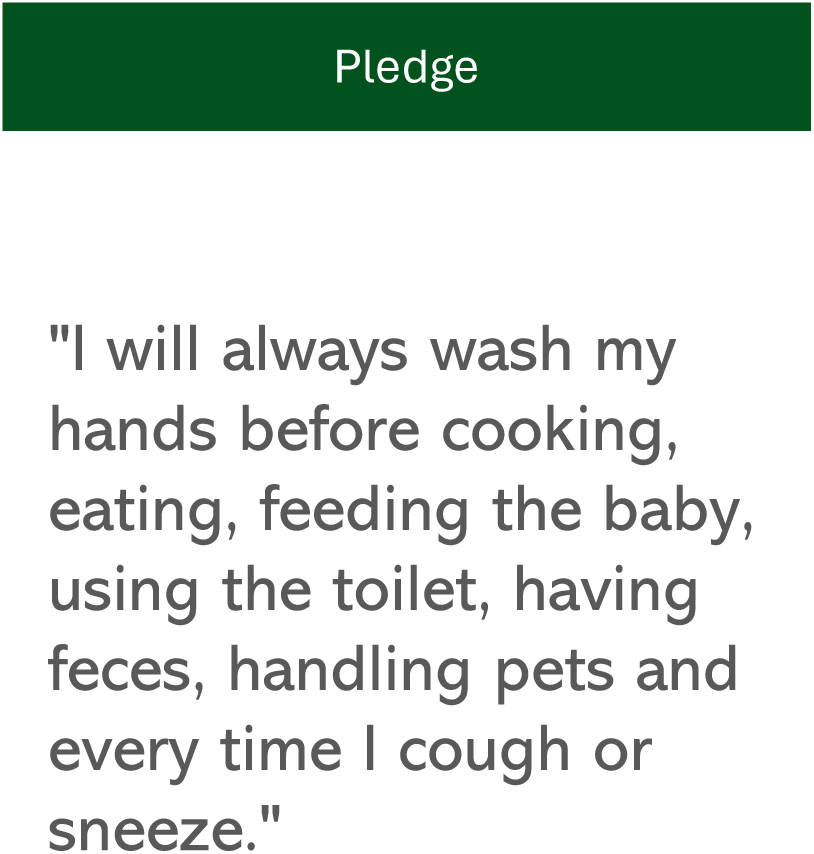

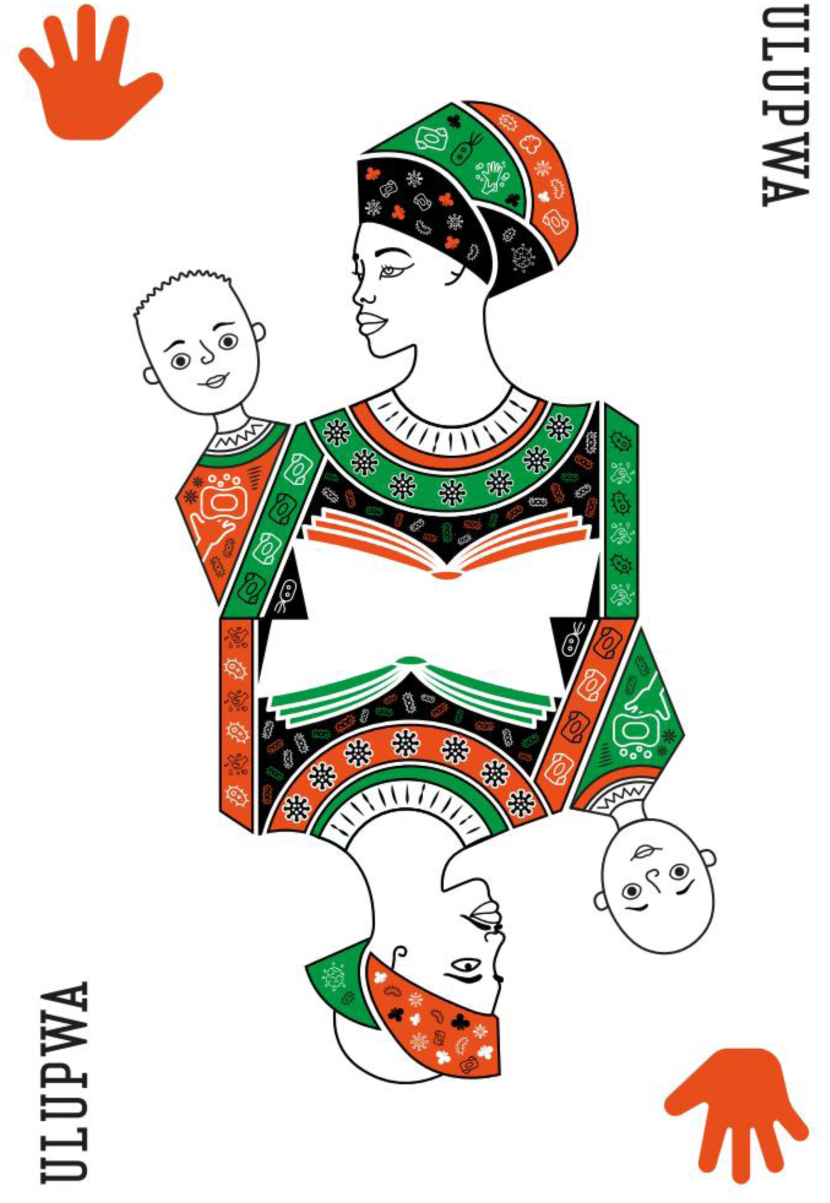

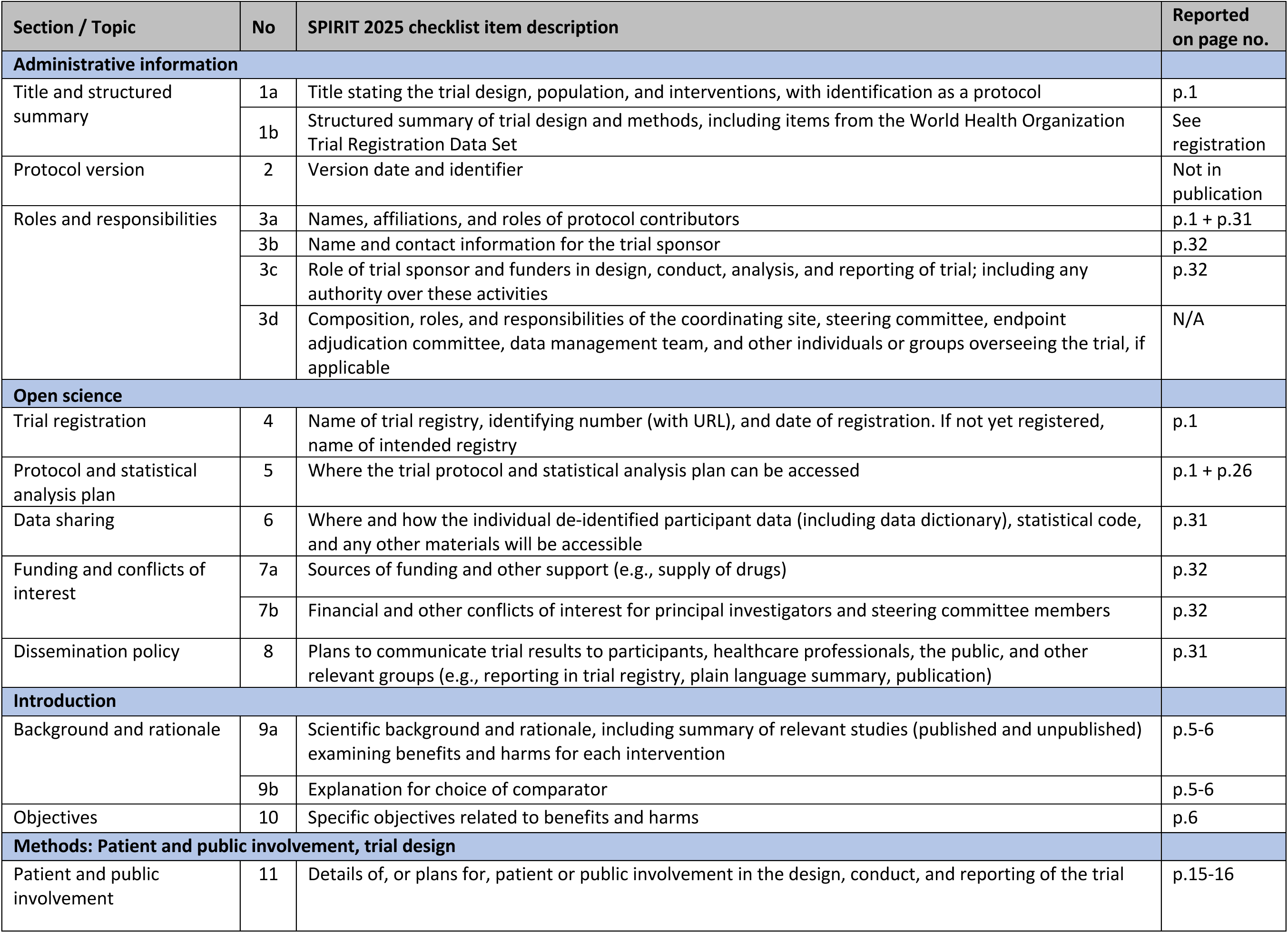
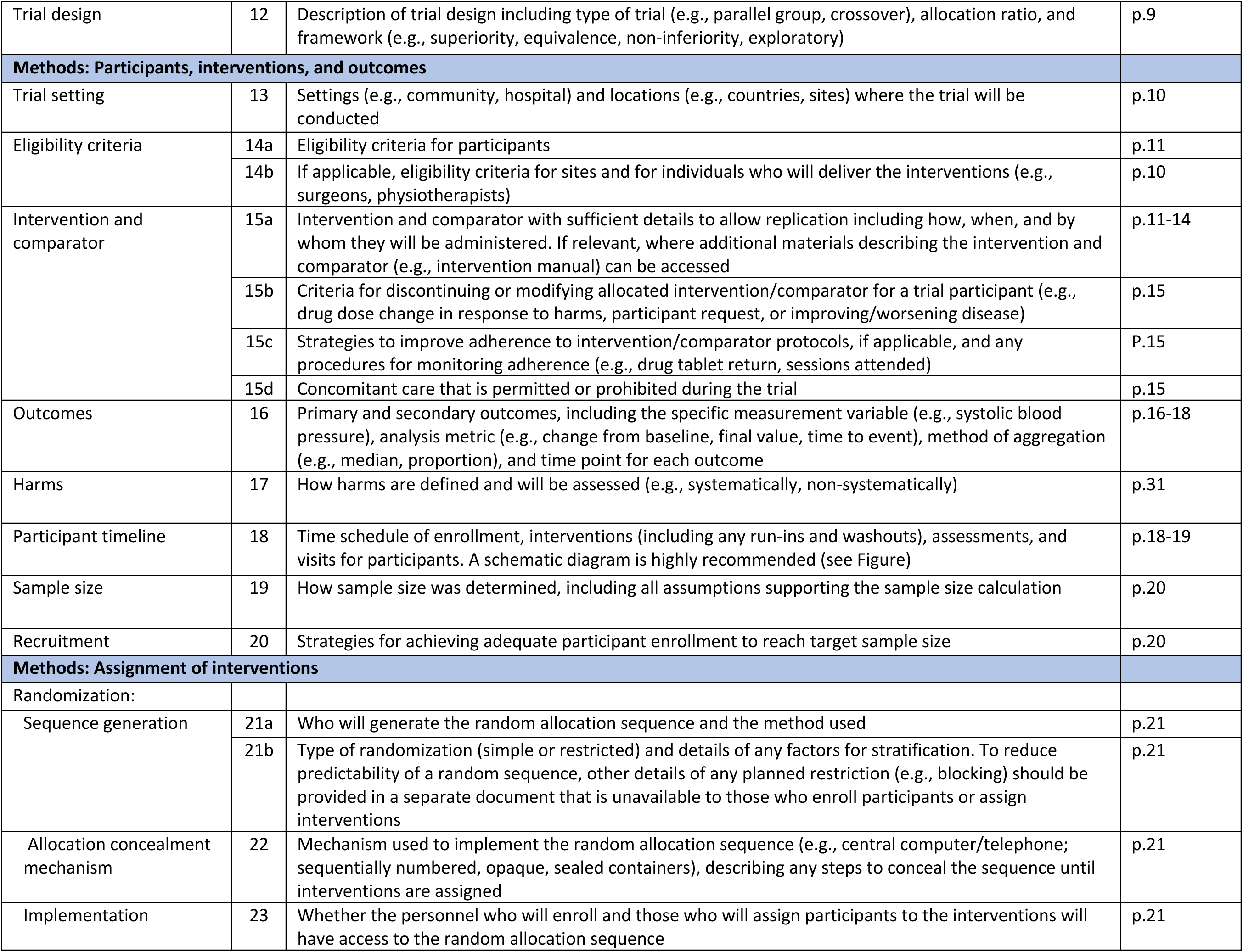
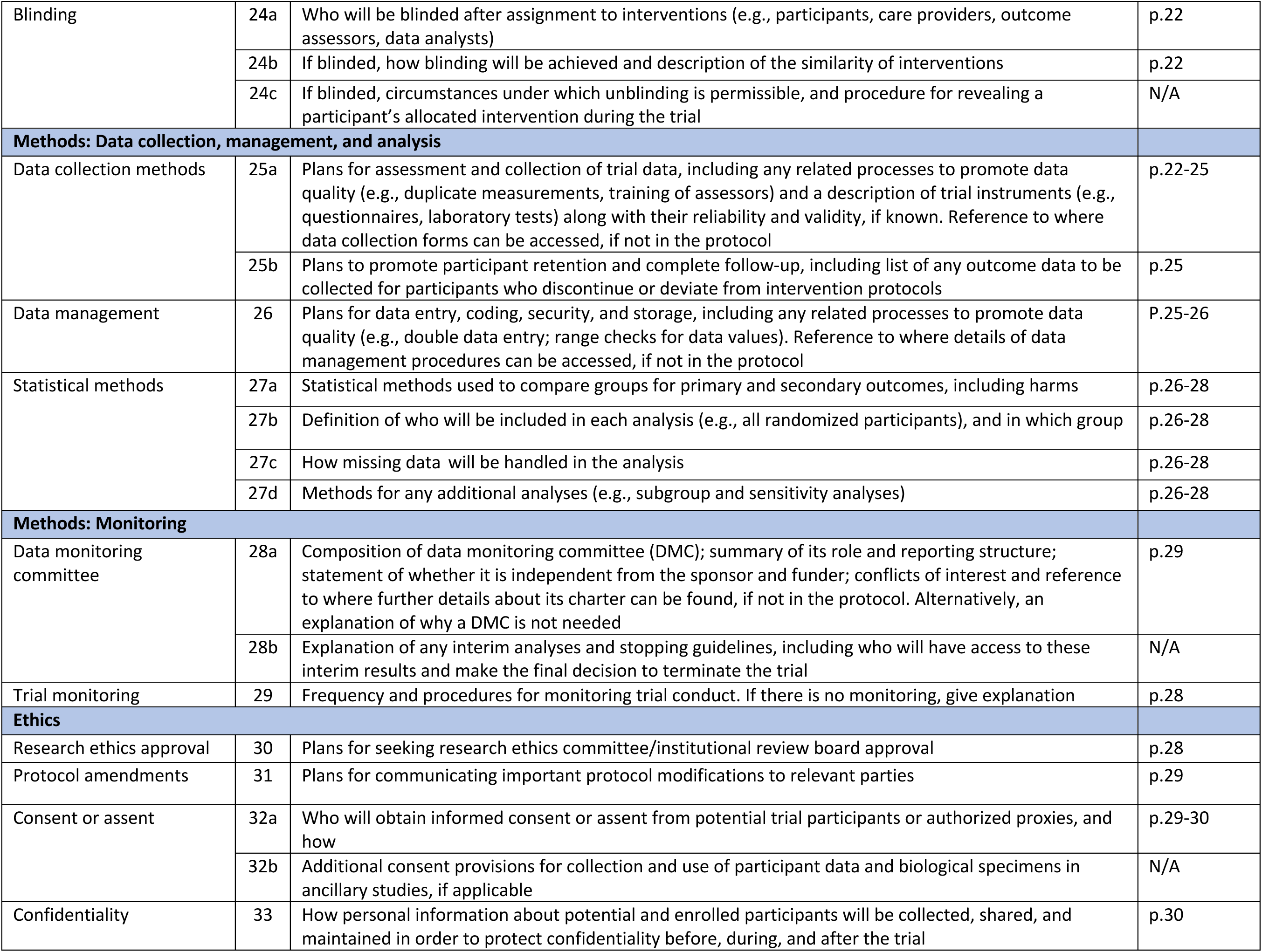
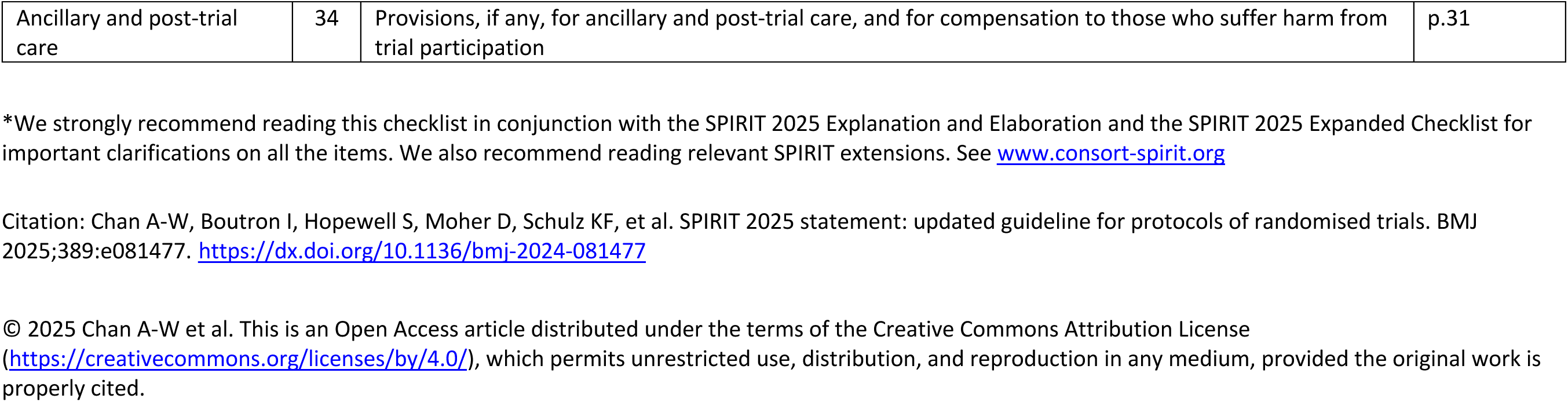
SPIRIT 2025 checklist of items to address in a randomized trial protocol*.

TiDieR-WASH Checklist.

TiDieR-WASH Checklist
| Item | Where is this information reported? |
| --- | --- |
| 1. Name | Intervention Section, page 11 |
| 2. Theory of Change | Intervention Section, pages 11-12 |
| 3. Prior evidence | Prior evidence supporting the use of multi-component interventions with hardware and behavioural components is detailed in the introduction (pages 5-6) |
| 4. Location and setting | Study setting, page 10 |
| 5. Context | Study setting, page 10 |
| 6. Suitability | Study setting, page 10 and Patient and Public Involvement Statement, page 15 |
| 7. Implementers | Intervention Section, page 12 |
| 8. Recipients | Study design, page 9 (characteristics not yet available) |
| 9. Targeting | Intervention Section, pages 11-12 |
| 10. Activities | Intervention Section, page 12-14 and Supplementary Material S1 Text |
| 11. Intervention dose | Not available |
| 12. Fidelity | Not available |
| 13. Cost | Not available |
| 14. Materials | Intervention Section, page 12-14 and Supplementary Material S1 Text |

## REFERENCES

1. Perin J, Mulick A, Yeung D, et al. Global, regional, and national causes of under-5 mortality in 2000-19: an updated systematic analysis with implications for the Sustainable Development Goals. Lancet Child Adolesc Health. 2022;6(2):106–15

2. Ross I, Bick S, Ayieko P, et al. Effectiveness of handwashing with soap for preventing acute respiratory infections in low-income and middle-income countries: a systematic review and meta-analysis. Lancet. 2023;401(10389):1681–90

3. Wolf J, Hubbard S, Brauer M, et al. Effectiveness of interventions to improve drinking water, sanitation, and handwashing with soap on risk of diarrhoeal disease in children in low-income and middle-income settings: a systematic review and meta-analysis. The Lancet. 2022;400(10345):48–59

4. Freeman MC, Stocks ME, Cumming O, et al. Systematic review: Hygiene and health: systematic review of handwashing practices worldwide and update of health effects. Tropical Medicine & International Health. 2014;19(8):906–16

5. Wolf J, Johnston R, Freeman MC, et al. Handwashing with soap after potential faecal contact: global, regional and country estimates. Int J Epidemiol. 2019;48(4):1204–18

6. Luby SP, Halder AK, Tronchet C, et al. Household characteristics associated with handwashing with soap in rural Bangladesh. Am J Trop Med Hyg. 2009;81(5):882–7

7. WHO/UNICEF Joint Monitoring Programme (JMP). Household WASH Data – Sub-national Regions Table. JMP Data Portal [dataset]. 2024 [accessed 04 September 2025]. Available from: https://washdata.org/data/household#!/table?geo0=region&geo1=sdg

8. Ezezika O, Heng J, Fatima K, et al. What are the barriers and facilitators to community handwashing with water and soap? A systematic review. PLOS Global Public Health. 2023;3(4):e0001720. doi: 10.1371/journal.pgph.0001720

9. White S, Thorseth AH, Dreibelbis R, et al. The determinants of handwashing behaviour in domestic settings: An integrative systematic review. International Journal of Hygiene and Environmental Health. 2020;227:113512

10. Caruso BA, Snyder JS, O’Brien LA, et al. Behavioural factors influencing hand hygiene practices across domestic, institutional, and public community settings: A systematic review. medRxiv. 2025:2025.03.11.25323561 [Preprint]. March 13, 2025. 10.1101/2025.03.11.25323561

11. De Buck E, Van Remoortel H, Hannes K, et al. Approaches to promote handwashing and sanitation behaviour change in low- and middle-income countries: a mixed method systematic review. Campbell Systematic Reviews. 2017;13(1):1–447

12. Staniford LJ, Schmidtke KA. A systematic review of hand-hygiene and environmental-disinfection interventions in settings with children. BMC Public Health. 2020;20(1):195.

13. MacLeod C, Braun L, Caruso BA, et al. Recommendations for hand hygiene in community settings: a scoping review of current international guidelines. BMJ Open. 2023;13(6):e068887. doi: 10.1136/bmjopen-2022-068887

14. Contzen N, Inauen J. Social-cognitive factors mediating intervention effects on handwashing: a longitudinal study. J Behav Med. 2015;38(6):956–69.

15. Amon-Tanoh MA, McCambridge J, Blon PK, et al. Effects of a social norm-based handwashing intervention including handwashing stations, and a handwashing station-only intervention on handwashing with soap in urban Cte d’Ivoire: a cluster randomised controlled trial. The Lancet Global Health. 2021;9(12):e1707–e18. doi: 10.1016/S2214-109X(21)00387-9

16. Prasad SK, Snyder JS, LaFon E, et al. Interventions to improve hand hygiene in community settings: A systematic review of theories, barriers and enablers, behavior change techniques, and hand hygiene station design features. medRxiv. 2025:2025.03.11.25323730 [Preprint]. March 12, 2025. 10.1101/2025.03.11.25323730

17. Richardson WS, Wilson MC, Nishikawa J, et al. The well-built clinical question: a key to evidence-based decisions. ACP journal club. 1995;123(3):A12–3.

18. Kahan BC, Hall SS, Beller EM, et al. Reporting of Factorial Randomized Trials: Extension of the CONSORT 2010 Statement. JAMA. 2023;330(21):2106–14.

19. Kahan BC, Tsui M, Jairath V, et al. Reporting of randomized factorial trials was frequently inadequate. Journal of Clinical Epidemiology. 2020;117:52–9.

20. Chan A-W, Boutron I, Hopewell S, et al. SPIRIT 2025 statement: updated guideline for protocols of randomised trials. BMJ. 2025;389:e081477.

21. Chiwele D, Lamson-Hall P, Wani S. Informal Settlements in Lusaka [online]. 2022. https://www.theigc.org/sites/default/files/2022/02/Informal-settlements-in-Lusaka-web.pdf (accessed 4 Sep 2025).

22. Hubbard SC, Meltzer MI, Kim S, et al. Household illness and associated water and sanitation factors in peri-urban Lusaka, Zambia, 2016–2017. npj Clean Water. 2020;3(1):26

23. WHO/UNICEF Joint Monitoring Programme (JMP). Household WASH Data – Zambia Table. JMP Data Portal [dataset]. 2024 [accessed 04 September 2025]. Available from: https://washdata.org/data/household#!/table?geo0=country&geo1=ZMB

24. Eubank N, Zambian 2006 to 2010 Constituency and Ward Boundaries. Standford Digital Respository [dataset]. 2014. Available from: http://purl.stanford.edu/yc436vm9005

25. Crocker J, Ogutu E, Snyder JS, et al. TIDieR-WASH: A Guideline for Reporting Implementation of Water, Sanitation, and Hygiene Interventions. Environ Health Perspect. 2024;132(11):117006.

26. Mwila-Kazimbaya K, Davies K, Mwenge MM, et al. Understanding end-user preferences for hand hygiene enabling technologies: A mixed-methods study in peri-urban Lusaka, Zambia. PLOS Water. 2025;4(8):e0000410. doi: 10.1371/journal.pwat.0000410

27. Ross I, Esteves Mills J, Slaymaker T, et al. Costs of hand hygiene for all in household settings: estimating the price tag for the 46 least developed countries. BMJ Global Health. 2021;6(12):e007361. doi: 10.1136/bmjgh-2021-007361

28. Mosler H-J. A systematic approach to behavior change interventions for the water and sanitation sector in developing countries: a conceptual model, a review, and a guideline. International Journal of Environmental Health Research. 2012;22(5):431–49.

29. Dreibelbis R, Winch PJ, Leontsini E, et al. The Integrated Behavioural Model for Water, Sanitation, and Hygiene: a systematic review of behavioural models and a framework for designing and evaluating behaviour change interventions in infrastructure-restricted settings. BMC Public Health. 2013;13(1):1015.

30. Aunger R, Curtis V. Behaviour Centred Design: towards an applied science of behaviour change. Health psychology review. 2016;10(4):425–46.

31. Rizk O, Bick S, White B, et al. Assessing the reliability and validity of pictorial-assisted 24-h recall for measuring hand hygiene and child faeces disposal: A cross-sectional study in Malawi. International Journal of Hygiene and Environmental Health. 2025;264:114516.

32. Freidlin B, Korn EL. Two-by-Two Factorial Cancer Treatment Trials: Is Sufficient Attention Being Paid to Possible Interactions? JNCI: Journal of the National Cancer Institute. 2017;109(9).

33. Zambia Statistics Agency, Ministry of Health (MOH) Zambia, and ICF. Zambia Demographic and Health Survey 2018 [online]. 2019. https://dhsprogram.com/pubs/pdf/FR361/FR361.pdf (accessed 4 Sep 2025).

34. Tidwell JB, Chipungu J, Chilengi R, et al. Theory-driven formative research on on-site, shared sanitation quality improvement among landlords and tenants in peri-urban Lusaka, Zambia. International Journal of Environmental Health Research. 2019;29(3):312–25.

35. Biran A, Rabie T, Schmidt W, et al. Comparing the performance of indicators of hand-washing practices in rural Indian households. Tropical Medicine & International Health. 2008;13(2):278–85.

36. Ram PK, Halder AK, Granger SP, et al. Is structured observation a valid technique to measure handwashing behavior? Use of acceleration sensors embedded in soap to assess reactivity to structured observation. Am J Trop Med Hyg. 2010;83(5):1070–6

37. Schmidt WP, Lewis HE, Greenland K, et al. Comparison of structured observation and pictorial 24 h recall of household activities to measure the prevalence of handwashing with soap in the community. International Journal of Environmental Health Research. 2019;29(1):71–81

38. Ram PK, Sahli MW, Arnold BF, et al. Validity of Rapid Measures of Handwashing Behavior: An Analysis of Data from Multiple Impact Evaluations in the Global Scaling Up Handwashing Project [online]. Washington, D.C.: World Bank Group Water and Sanitation Program; 2014. https://globalhandwashing.org/wp-content/uploads/2015/03/Validity-of-Rapid-Measures-Handwashing-Behavior.pdf (accessed 4 Sep 2025).

39. Hopewell S, Chan A-W, Collins GS, et al. CONSORT 2025 statement: updated guideline for reporting randomised trials. BMJ. 2025;389:e081123. doi: 10.1136/bmj-2024-081123

40. Juszczak E, Altman DG, Hopewell S, et al. Reporting of Multi-Arm Parallel-Group Randomized Trials: Extension of the CONSORT 2010 Statement. Jama. 2019;321(16):1610–20.

41. Korn EL, Allegra CJ, Freidlin B. Phase III Evaluation of Treatment Combinations in Three-Arm Trials. Journal of Clinical Oncology. 2025;43(2):226–33

42. Young SL, Boateng GO, Jamaluddine Z, et al. The Household Water InSecurity Experiences (HWISE) Scale: development and validation of a household water insecurity measure for low-income and middle-income countries. BMJ Glob Health. 2019;4(5):e001750. doi: 10.1136/bmjgh-2019-001750

